# Characterising patterns of COVID-19 and long COVID symptoms: Evidence from nine UK longitudinal studies

**DOI:** 10.1101/2022.06.20.22275994

**Authors:** Ruth C. E. Bowyer, Charlotte Huggins, Renin Toms, Richard J. Shaw, Bo Hou, Ellen J. Thompson, Alex Kwong, Dylan Williams, Milla Kibble, George B. Ploubidis, Nic Timpson, Jonathan Sterne, Nish Chaturvedi, Claire J. Steves, Kate Tilling, Richard J. Silverwood, the CONVALESCENCE Study

## Abstract

Multiple studies across global populations have established the primary symptoms characterising COVID-19 (Coronavirus Disease 2019) and long COVID. However, as symptoms may also occur in the absence of COVID-19, a lack of appropriate controls has often meant that specificity of symptoms to acute COVID-19 or long COVID could not be examined. We aimed to characterise patterns of COVID-19 and long COVID symptoms across nine UK longitudinal studies, totalling over 42,000 participants. Conducting latent class analyses separately in three groups (‘no COVID-19’, ‘COVID-19 in last 12 weeks’, ‘COVID-19 > 12 weeks ago’), the data did not support the presence of more than two distinct symptom patterns, representing high and low symptom burden, in each group. Comparing the high symptom burden classes between the ‘COVID-19 in last 12 weeks’ and ‘no COVID-19’ groups we identified symptoms characteristic of acute COVID-19, including loss of taste and smell, fatigue, cough, shortness of breath and muscle pains or aches. Comparing the high symptom burden classes between the ‘COVID-19 > 12 weeks ago’ and ‘no COVID-19’ groups we identified symptoms characteristic of long COVID, including fatigue, shortness of breath, muscle pain or aches, difficulty concentrating and chest tightness. The identified symptom patterns among individuals with COVID-19 > 12 weeks ago were strongly associated with self-reported length of time unable to function as normal due to COVID-19 symptoms, suggesting that the symptom pattern identified corresponds to long COVID. Building the evidence base regarding typical long COVID symptoms will improve diagnosis of this condition and the ability to elicit underlying biological mechanisms, leading to better patient access to treatment and services.

## INTRODUCTION

Hundreds of millions of people worldwide have now been infected with SARS-CoV-2, the coronavirus responsible for the COVID-19 (coronavirus disease 2019) pandemic (1, 2). SARS-CoV-2 enters the body via the Angiotensin-converting enzyme 2 (ACE-2) receptor; as ACE2 is located on cells across multiple body sites the virus has the capacity to infect and damage cells within multiple organs (3, 4). This is reflected in the variety of symptoms associated with acute COVID-19 (signs and symptoms of COVID-19 for up to 4 weeks), ongoing symptomatic COVID-19 (signs and symptoms of COVID-19 from 4-12 weeks) and in post-COVID-19 syndrome (where signs and symptoms developing during or after COVID-19 infection continue for more than 12 weeks and are not explained by an alternative diagnosis) (5). Both ongoing symptomatic COVID-19 and post-COVID-19 syndrome are regarded under the umbrella of long COVID, as patient advocates prefer the condition to be termed (6). However, for the purpose of understanding difference in symptomology at various stages of illness, we consider only symptoms greater than 12 weeks as long COVID as long term symptoms are likely to have a more detrimental impact on quality of life.

Whilst multiple studies across global populations have established the primary symptoms characterising long COVID to include fatigue, shortness of breath, cough, cognitive impairment and anosmia, a plethora of persistent symptoms have been reported by patients (7, 8). Better understanding of symptoms which characterise long COVID – or subvariants thereof and thus whether it is meaningful to describe long COVID as one syndrome (1) – may improve diagnostic precision and help elicit underlying mechanisms to better target therapy via patient-centred strategies (3, 9-14). Further, as these symptoms often also occur in the absence of COVID-19, the lack of appropriate controls has often meant that specificity of symptoms to acute COVID-19 or long COVID could not be examined. Inadequate consideration of cohort selection biases, particularly where participants have been recruited via support groups, may undermine generalisability of findings and therefore their utility in guiding clinical practice (15).

We aimed to characterise patterns of symptoms in individuals who had experienced COVID-19, before and after twelve weeks of illness onset, as well as those who had not, across nine UK longitudinal studies, to shed light on specific symptom patterns of COVID-19 and long COVID. We then examined how patterns differed by key factors such as sex, age and (for long COVID) self-reported functional limitation following COVID-19.

## METHODS

### Data

The UK National Core Studies – Longitudinal Health and Wellbeing programme (https://www.ucl.ac.uk/covid-19-longitudinal-health-wellbeing/) combines data from multiple UK population-based longitudinal studies and electronic health records to conduct analyses that allow researchers to investigate pandemic-related changes in population health. As symptom persistence is poorly captured in electronic health records, we performed co-ordinated standardised analyses across multiple longitudinal studies. This approach minimises methodological heterogeneity and maximises comparability, while appropriately accounting for study designs and characteristics of individual datasets. Meta-analyses of key study-specific estimates were performed, maximising statistical power and representativeness.

We analysed data from nine UK longitudinal studies. Four of the studies are birth cohorts, containing participants of a similar age (age-homogeneous): National Child Development Study (NCDS; born 1958) (16, 17), British Cohort Study (BCS70; born 1970) (17, 18), Next Steps (NS; born 1989-90) (17, 19) and Millennium Cohort Study (MCS; born 2000-02) (17, 20). The remaining five studies covered a wider range of ages (age-heterogeneous): Avon Longitudinal Study of Parents and Children (ALSPAC)^1^ (21, 22), TwinsUK (23, 24), Born in Bradford (BiB) (25, 26), Understanding Society (USoc) (27, 28) and Generation Scotland (GS) (29, 30). Full details of the studies are provided in Table S1 (Supplementary Material), with ethics and data access statements in Table S2 (Supplementary Material).

Information relating to COVID-19 and symptoms was obtained from questionnaires completed by study participants between July 2020 and September 2021 (periods differed by study).

### Variables

Here we provide an overview of the variables used in the analysis. Further details of how information was captured and variables derived in each study are provided in Methods S1 (Supplementary Material).

Symptoms: In each study, respondents reported the presence of different individual symptoms, such as fever, cough and sore throat, regardless of whether they attributed these symptoms to any specific cause. Primary clustering analyses were undertaken using a “core” set of symptoms which were (almost) all available in all studies to aid between-study comparability; secondary clustering analyses were undertaken in a subset of studies using a “maximal” set of symptoms to allow a broader exploration of symptom patterns. The symptoms included in each set are shown in Table 1. The period over which the presence of symptoms was reported also differed by study, between two weeks and two months. In two studies (TwinsUK, USoc), symptoms were observed at multiple timepoints for each individual with the presence or absence of each symptom derived for each symptom timepoint.

**Table 1.**
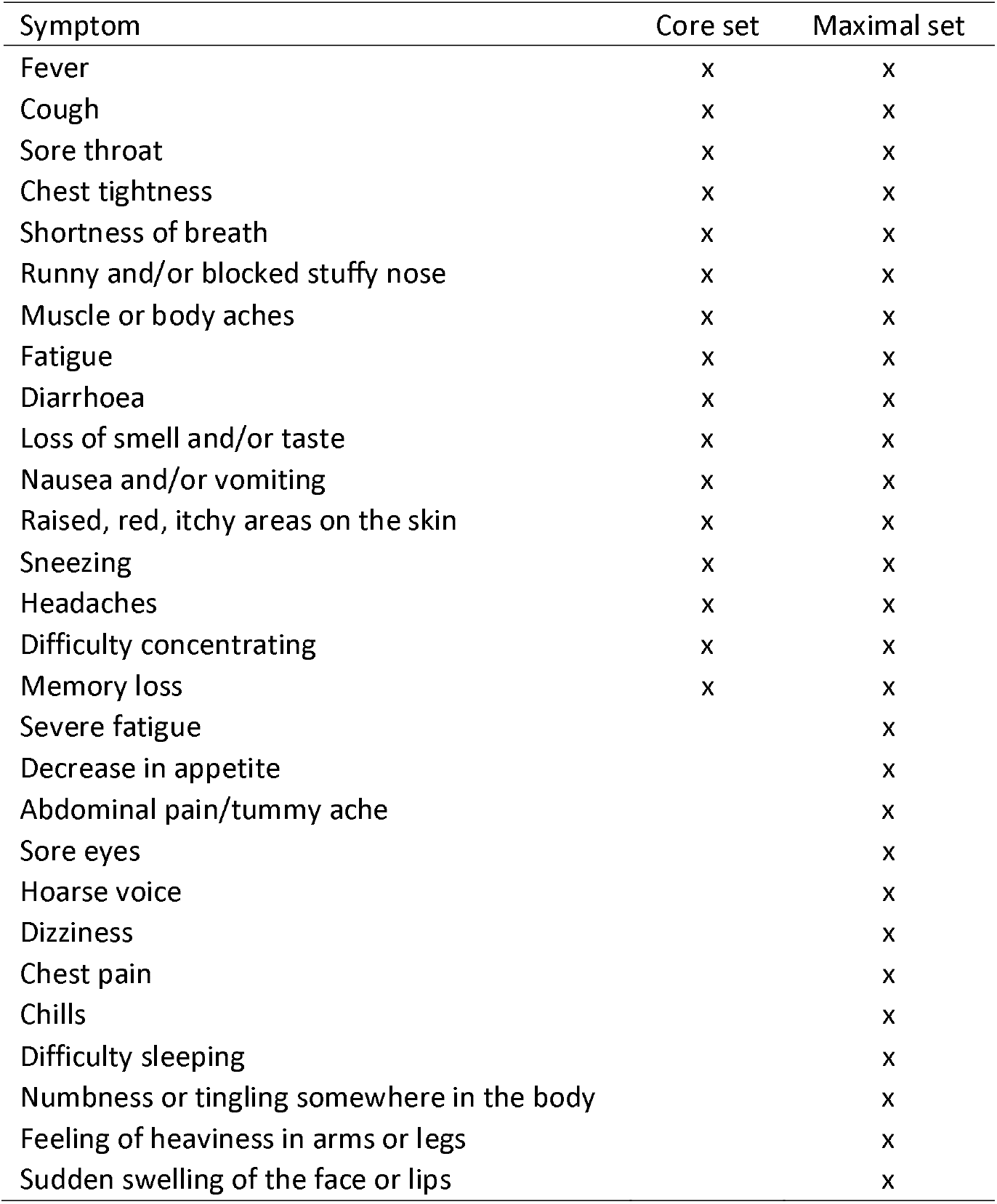
Core and maximal symptom sets.

Functional limitation following COVID-19: This was asked about in NCDS, BCS70, NS, MCS and TwinsUK only using the question *“For how long were you unable to function as normal due to Coronavirus symptoms?”* (or a subtle variation thereof).

COVID-19: Prior or current COVID-19 was self-reported in all studies. Among individuals reporting prior or current COVID-19, time since COVID-19 onset at the point of symptom reporting was derived using the date of the symptom timepoint and the reported date of COVID-19 onset (complete date or month and year only depending on study). For cohorts unable to derive time since COVID-19 onset (USoc, GS), self-reported symptom length was instead used. We derived a COVID-19 status indicator (time-varying for studies with multiple symptom timepoints (TwinsUK, USoc)) using information on prior COVID-19, time since COVID-19, and functional limitation at 12 weeks post-COVID-19, with categories:

1. No COVID-19
2. COVID-19 in last 12 weeks
3. COVID-19 > 12 weeks ago + no functional limitation at 12 weeks
4. COVID-19 > 12 weeks ago + functional limitation at 12 weeks

In studies where data on functional limitation were not collected (ALSPAC, BiB, USoc, GS), categories 3 and 4 could not be differentiated so were pooled. In studies which only collected symptom data for participants who reported prior COVID-19 (USoc, GS), category 1 was not present and only individuals with prior COVID-19 were analysed. We emphasise that these categories capture only time since reported COVID-19 at the point of symptom reporting and self-reported functional limitation at 12 weeks; they do not necessarily suggest that reported symptoms were due to COVID-19, which is why we make use of the symptoms reported in the ‘No COVID-19’ group.

Sex: Sex (male/female) was obtained from responses to the same or earlier questionnaires.

Age: Age at each symptom timepoint was derived from the date of the symptom timepoint and the date of birth reported at the same or earlier questionnaires (age-heterogeneous studies) or the known common date of birth (age-homogeneous studies).

### Statistical analysis

Individual symptom analyses: For each available symptom within each study, the number and percentage of participants reporting the symptom within each COVID-19 group were tabulated. Logistic regression (for studies with a single symptom timepoint), logistic generalised estimation equations (GEE) with clustering by participant identifier and an unstructured correlation matrix (USoc) or fixed effects logistic regression (TwinsUK; due to non-convergence of GEE approach) were used to estimate odds ratios (ORs) comparing symptom presence in the COVID-19 groups. The ‘no COVID-19’ category was considered as the reference group, except in studies where symptom data were only available for participants who had reported prior COVID-19 (GS, USoc). In such cases the ‘COVID in last 12 weeks’ category was considered the baseline group. Models were adjusted for sex (male/female), age (age-heterogeneous studies only; continuous) and calendar time (for most studies, month). Survey design weights (where necessary) and non-response weights (where available) were used. To be included in these analyses, study participants needed to have observed data on a given symptom (plus calendar time and age, though these were fully observed). ORs for symptoms within the core symptom set were subsequently combined across studies using random-effects meta-analysis. These analyses were intended to be descriptive, providing an exploration of the symptom data prior to undertaking the clustering analyses.

Symptom clustering analyses: We conducted, within each study, latent class analyses (LCAs) of reported symptoms separately within each category of the previously derived COVID-19 status indicator. This was undertaken separately in primary (core symptom set) and secondary (maximal symptom set) analyses following an identical procedure. Due to small numbers in the ‘COVID-19 > 12 weeks ago + functional limitation at 12 weeks’ group, this category was combined with the ‘COVID-19 > 12 weeks ago + no functional limitation at 12 weeks’ category to form a ‘COVID-19 > 12 weeks ago’ category. All individuals within each study with observed data on symptoms from at least one wave of data collection were included in the LCA.

We fitted LCAs of symptoms with increasing numbers of classes, from 1 to 5, unless non-convergence occurred first. Where available, calendar time (in months for most studies) of symptom observation or wave of data collection was allowed to affect latent class membership. Study design weights (if applicable) and non-response weights (if available) were utilised. Sufficient different starting values were used to ensure that the obtained maximum likelihood solution was replicated. Full information maximum likelihood was used to handle a small amount of missing symptom data in some studies. For each obtained LCA solution, we noted model fit statistics (Akaike information criterion (AIC), Bayesian information criterion (BIC), adjusted-BIC), entropy (a summary measure of the certainty with which individuals can be allocated to classes) and the percentage of individuals in the smallest class. The optimal number of latent classes in each LCA was determined through consideration of the model fit statistics. The information criteria were plotted against the number of classes and the optimal number chosen through identification of a point of inflection in the BIC curve (31), with the additional criterion that the smallest class must be >5% of the total sample. The LCA outputs of primary interest were the number of classes supported by the data in each COVID-19 group and the probability of each symptom within each latent class (i.e. how the symptom pattern could be characterised). Formal quantitative cross-study synthesis (e.g. meta-analysis) of the symptom patterns was not undertaken, with a more qualitative approach utilised.

Associations with symptom patterns: For the core symptom set findings, subsequent analyses used logistic regression to examine how patterns differed by sex, age group (age-heterogeneous studies only) and self-reported functional limitation post-COVID onset (individuals with COVID-19 onset > 12 weeks ago only). Given the high entropy values observed, participants were allocated to their most likely latent class (symptom pattern) according to their posterior probabilities of class membership. Inclusion in each of these analyses had the additional requirement of complete data on the relevant variable. ORs were subsequently combined across studies using random-effects meta-analysis.

## RESULTS

Across the nine studies we analysed a total of over 42,000 individuals, considering between 16 and 28 symptoms reported across 15 months of the pandemic (July 2020 to September 2021).

### Individual symptom analyses

For each study, the number and percentage of participants reporting each symptom within each COVID-19 group are reported in Table S4 (Supplementary Material). An important observation is that individuals within the ‘no COVID-19’ group reported moderate levels of many symptoms, including some of those commonly associated with COVID-19. For example, headaches were reported by between 16.9% and 28.3% of those with no reported COVID-19 in each study, fatigue by between 15.2% and 28.0%, and muscle or body aches or pains by between 7.1% and 21.0%.

Estimated ORs comparing symptom presence in the COVID-19 groups within each study are also reported in Table S4 (Supplementary Material); meta-analysed ORs are presented in Fig. 1 with the underlying data in Table S5 (Supplementary Material). A small proportion of ORs were not estimable due to low symptom prevalence in one or more COVID-19 groups. Whilst there was considerable heterogeneity between studies, many symptoms had a higher prevalence in the ‘COVID-19 in last 12 weeks’ group than in the ‘no COVID-19’ group, with this being most marked for loss of smell (meta-analysed OR 28.6; 95% confidence interval (CI) 16.6, 49.2), loss of taste (20.5; 15.4, 27.4), fever (5.5; 4.3, 7.1), cough (3.6; 2.0, 6.3) and shortness of breath (3.1; 2.6, 3.8). The relative prevalence of all symptoms was lower in the ‘COVID-19 > 12 weeks ago + no functional limitation at 12 weeks’ group than in the ‘COVID-19 in last 12 weeks’, though remained particularly elevated (relative to ‘no COVID-19’ group) for loss of smell (6.8; 4.4, 10.5) and loss of taste (4.2; 3.1, 5.8), with other ORs no greater than 2. In the ‘COVID-19 > 12 weeks ago + functional limitation at 12 weeks’ group there were again many symptoms with raised prevalence relative to the ‘no COVID-19’ group including, in addition to loss of taste (33.1; 9.8, 111.5) and loss of smell (26.3; 7.7, 89.2), fatigue (13.7; 6.9, 27.3), shortness of breath (11.9; 5.3, 26.6), muscle pain or aches (9.7; 6.0, 15.8), chest tightness (7.3; 4.3, 12.3), memory loss (6.3; 3.1, 13.0) and difficulty concentrating (6.2; 1.4, 28.0).

**Fig. 1.**
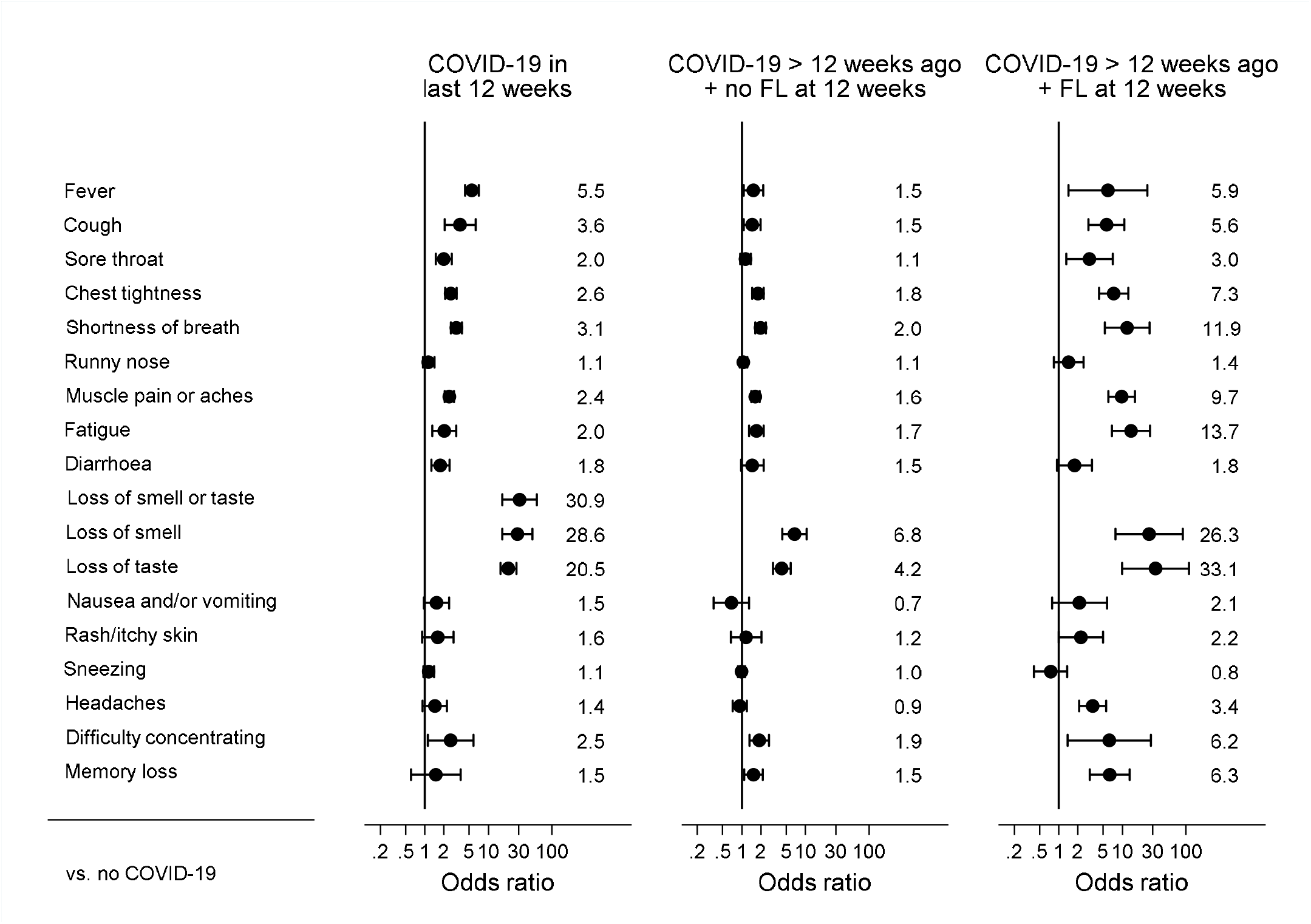
Odds ratio of each symptom for each COVID-19 group relative to the no COVID-19 group. Bars represent 95% confidence intervals. Estimates are from random effects meta-analyses of study-specific estimates.

### Symptom clustering analyses

For the primary analysis using the core symptom set, the data did not support more than two latent classes (symptom patterns) in each COVID-19 group within each study (LCA model fit statistics in Table S6 (Supplementary Material))^2^. The probability of each symptom within each symptom pattern are shown in Fig. S1 (Supplementary Material) for each study. In each instance, we identified one pattern (‘symptom pattern 1’ in the figures) that was characterised by a generally low prevalence of symptoms. A second pattern (‘symptom pattern 2’) was characterised by a higher prevalence of many symptoms, though precisely which symptoms had particularly high prevalence differed by COVID-19 group. The general similarity of symptom pattern 1 across the COVID-19 groups in each study suggests that this pattern identifies subgroups of similar individuals who, although they may have non-negligible probability of common symptoms such as a runny nose or a headache, were essentially well. The higher symptom burden within symptom pattern 2 therefore identifies individuals who are unwell, either due to non-COVID-19-related reasons (in the case of the ‘no COVID-19’ group) or due to a combination of COVID-19- and non-COVID-19-related reasons (as in the ‘COVID-19 in last 12 weeks’ and ‘COVID-19 > 12 weeks ago’ groups). Through a comparison of symptom pattern 2 between the two groups with COVID-19 and the no COVID-19 group we can explore which symptoms have the greatest excess probability relative to the COVID-19-free population, allowing us to identify symptoms that are typical of more acute COVID-19 and of long COVID.

Such comparisons can be more easily made using plots of the absolute probability differences, presented and in Fig. S2 (Supplementary Material) for each study^3^. To further aid cross-study interpretation of these findings, we have plotted the symptom probability differences for all available studies together on a single heatmap (Fig. 2). Although there was variability between studies, some common features were observed. The symptoms most consistently observed to have excess probability among individuals with COVID-19 in the last 12 weeks were loss of taste, loss of smell, fatigue, cough (particularly dry cough), shortness of breath, muscle pains or aches, fever, headaches and difficulty concentrating. The 95% CIs for these excess probabilities almost always excluded the null within each study, providing compelling evidence that these symptoms can be considered characteristic of more acute COVID-19. The symptoms most consistently observed to have excess probability among individuals with COVID-19 > 12 weeks ago were fatigue, shortness of breath, muscle pain or aches, difficulty concentrating, chest tightness, loss of smell, memory loss and loss of taste. The 95% CIs for these excess probabilities did not always exclude the null within each study, but the consistency of the findings across the cohorts provides strong evidence that these symptoms can be considered characteristic of long COVID.

**Fig. 2.**
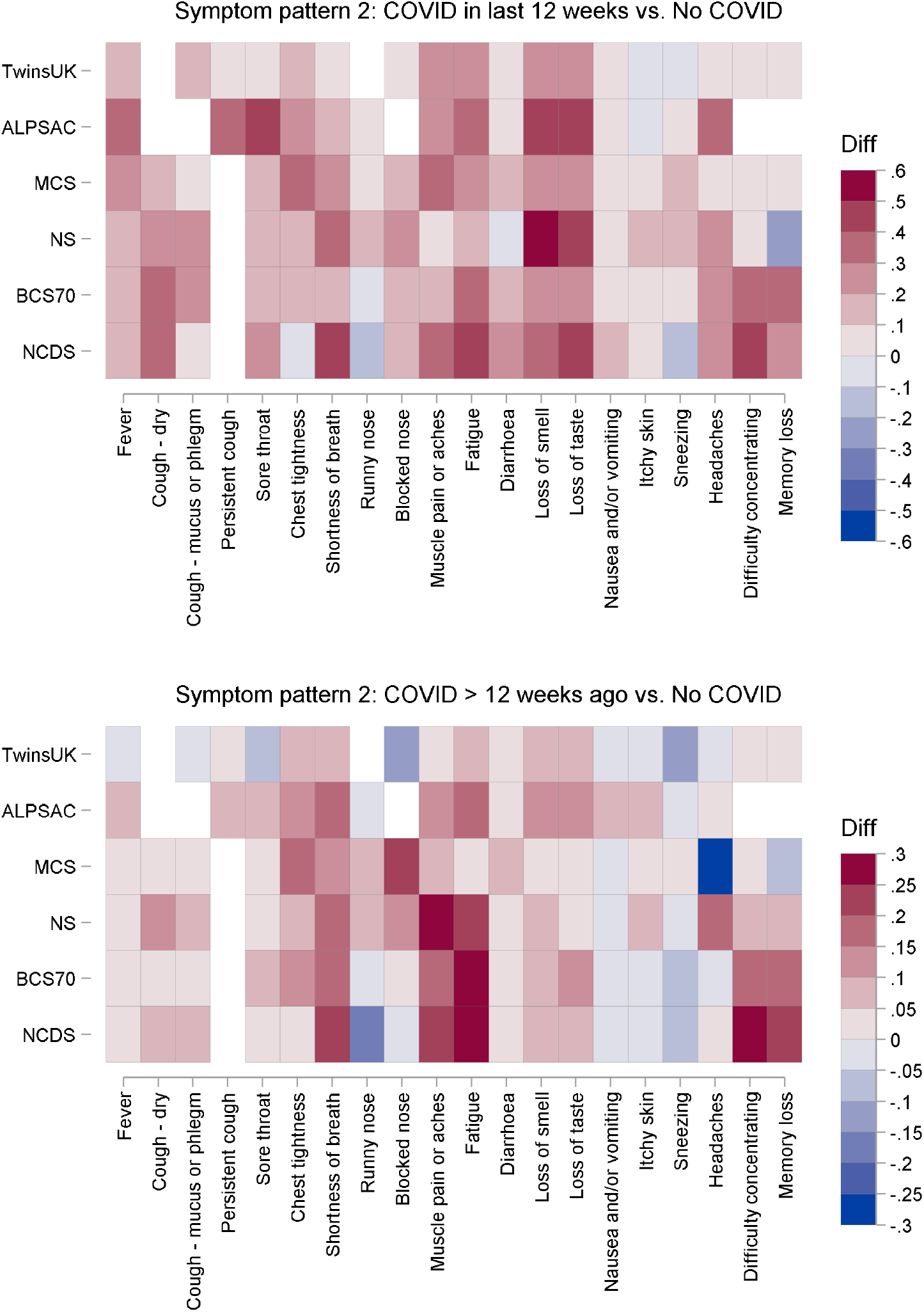
Heatmap of symptom probability differences comparing the two COVID-19 groups with the no COVID-19 group across all studies. NCDS: 1958 National Child Development Study; BCS1970: 1970 British Cohort Study; NS: Next Steps; MCS: Millennium Cohort Study; ALSPAC: Avon Longitudinal Study of Parents and Children. Notes: White cells indicate that the symptom was not asked about in in that study; ALSPAC results for “loss of smell or taste” are duplicated in the “loss of smell” and “loss of taste” cells; TwinsUK results for “confusion” are duplicated in the “difficulty concentrating” and “memory loss” cells.

In the secondary analysis using the maximal symptom set, the data again did not support more than two latent classes (symptom patterns) in each COVID-19 group within each study (Table S7 (Supplementary Material))^4^. The probability of each symptom within each symptom pattern are shown in Fig. S3 (Supplementary Material) for each study, with the absolute probability differences plotted in Fig. S4 (Supplementary Material). Additional symptoms observed to have excess probability among individuals with COVID-19 in the last 12 weeks were chills and heaviness in arms or legs. Among individuals with COVID-19 > 12 weeks ago, only heaviness in arms or legs was additionally identified.

### Associations with symptom patterns

Estimated associations with symptom patterns are shown in Table S8 (Supplementary Material) for each study; meta-analysed ORs are presented in Fig. 3 with the underlying data in Table S9 (Supplementary Material). The majority of individuals who had COVID-19 were unable to function as normal for less than two weeks (between 58.9% and 88.0% across the studies), with relatively few unable to function as normal for 12 weeks or more (1.5% to 7.4%). Across almost all studies there was consistent evidence that symptom pattern 2 (corresponding to a higher symptom burden) was more common among females in each of the COVID-19 groups (e.g. meta-analysed OR 1.6; 95% CI 1.3, 1.9 in the COVID-19 > 12 weeks ago group). In the age-heterogeneous studies there was evidence that symptom pattern 2 was more common at younger ages in the no COVID-19 group and, to a lesser extent, in the COVID-19 > 12 weeks ago group. Findings relating to functional limitation following COVID-19 were clear and consistent: the prevalence of symptom pattern 2 was greater for individuals who were unable to function as normal for longer, being particularly high in those who were unable to function for 12 weeks or more (meta-analysed OR 8.1; 95% CI 4.5, 14.4 relative to always being able to function as normal).

**Fig. 3.**
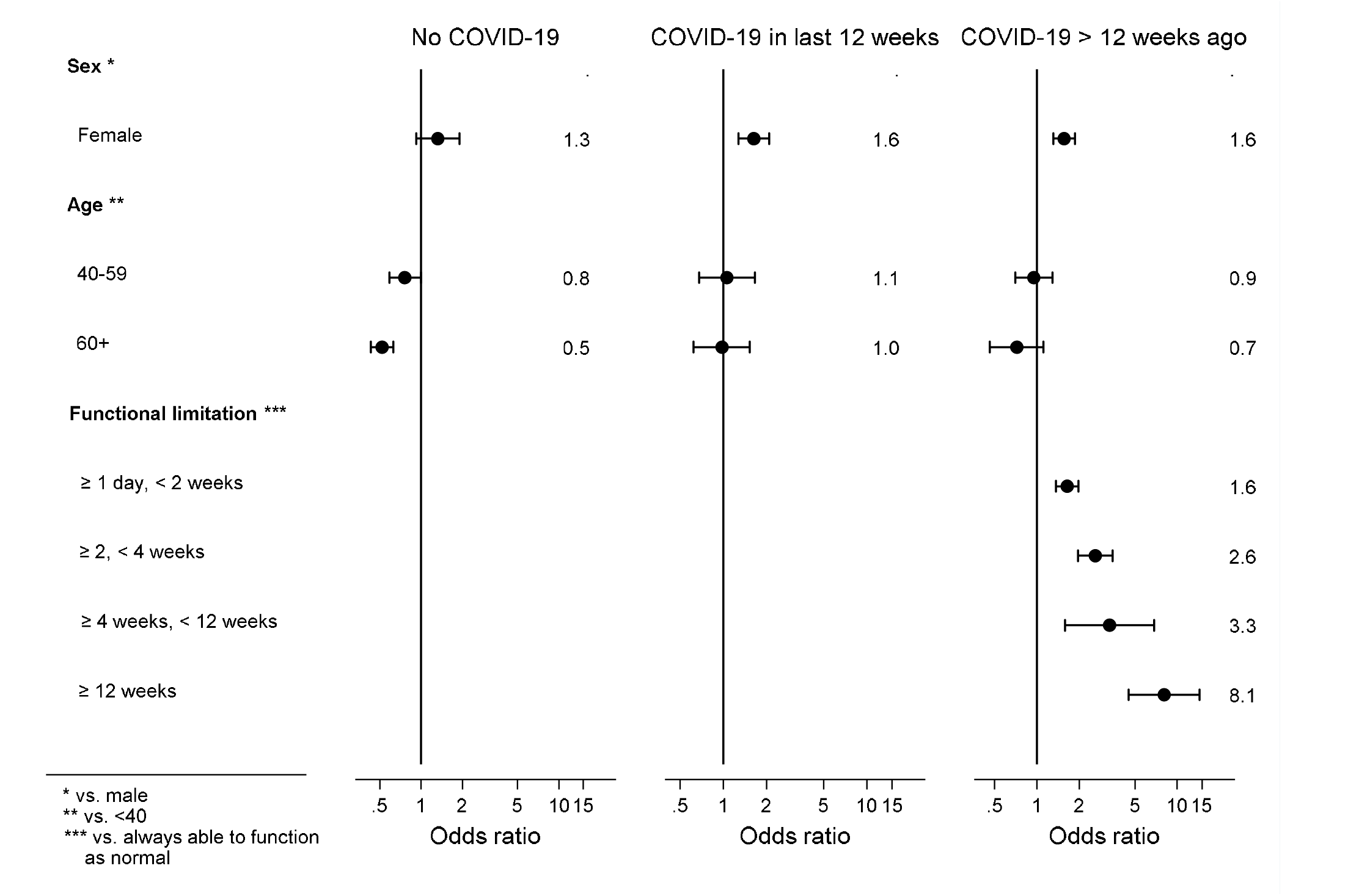
Associations with symptom pattern 2 (vs. symptom pattern 1) meta-analysed across studies. Bars represent 95% confidence intervals. Estimates are from random effects meta-analyses of study-specific estimates.

## DISCUSSION

We have characterised patterns of COVID-19 and long COVID symptoms across nine UK longitudinal studies and examined how patterns differed by key factors such as sex, age and (for long COVID) self-reported functional limitation following COVID-19.

In analyses of individual symptoms, we found replication of known symptoms of COVID-19, in that fever, cough and loss of smell and taste all had highest prevalence in the group with COVID within the past 12 weeks. This suggests that despite using self-reported COVID, and asking people to recall symptoms over varying periods, our results have face validity. The prevalence of some symptoms varied across studies; this could be due to seasonality, the different variants during different stages of the pandemic or just between-study differences in age, geography or other factors. The prevalence of runny nose and sneezing did not seem to differ between those with and without COVID-19, or those with COVID-19 within the past 12 weeks or longer ago, suggesting that these symptoms tend not to be COVID-19-specific.

In the symptom clustering analyses, the data did not support more than two symptom patterns among any of the COVID-19 groups in any of the studies, though relatively small sample sizes in these groups may have affected our ability to identify further symptom patterns of low prevalence. Other studies using differing clustering methods and study designs have found greater than two symptom patterns annotated as distinct symptoms sets when studying acute COVID-19 (32, 33) and long COVID (34). However, some studies have similarly found two symptom patterns to best fit the data: in the Norwegian Mother, Father and Child Cohort Study, Caspersen et al 2022 (1) analysed 73,727 adults followed throughout the pandemic and observed distinct patterns of post-acute symptoms characterised as ‘neurocognitive’ and ‘cardiorespiratory’. The symptoms were captured at 12 months post-infection, so it could be that symptoms become more disaggregated at a longer time interval since initial infection. Reflecting our results, Peluso et al (35) observed patients to group into two clusters, one reflecting high symptom prevalence and the other representing low, although in their approach they first aggregated reported symptoms to seven domains. Considered with our results, this does not support the idea that long COVID may be multiple syndromes discernible by their difference in symptom pattern.

Symptom pattern 2 (characterised by higher symptom burden) was generally more common among individuals with COVID-19 in the last 12 weeks or COVID-19 > 12 weeks ago than among those with no COVID-19, suggesting that, whilst there is a significant symptom burden among those who have never had COVID-19, this is greater in those who have had COVID-19. The presence of a substantial group of individuals without COVID-19 reporting a relatively high symptom burden emphasises the importance of a control group in analyses of COVID-19 symptoms.

Although symptom pattern 2 corresponded to a generally higher symptom burden, the precise symptom profile differed between COVID-19 groups. Although there was some between-study variability, the symptoms identified through comparison of the ‘COVID-19 in last 12 weeks’ and ‘no COVID-19’ groups as being characteristics of acute COVID-19 and ongoing symptomatic COVID-19 were loss of taste, loss of smell, fatigue, cough (particularly dry cough), shortness of breath, muscle pains or aches, fever, headaches and difficulty concentrating. Several meta-analyses have reported similarly, with fatigue, cough and alterations to taste and smell being characteristic of acute COVID-19 (36-38).

Symptoms identified through comparison of the ‘COVID-19 > 12 weeks ago’ and ‘no COVID-19’ groups as being characteristic of long COVID were fatigue, shortness of breath, muscle pain or aches, difficulty concentrating, chest tightness, loss of smell, memory loss and loss of taste. These symptoms are comparable to those identified in the existing literature. A clinical review by Crook and colleagues similarly found shortness of breath, impaired cognition, chest pain and in particular fatigue to characterise long COVID (3). Similarly, a meta-analysis by Martimbianco et al. (14) observed chest pain, fatigue, shortness of breath and cough as the symptoms characterising long COVID. However, these studies were not able to incorporate a ‘no COVID-19’ group to account for baseline population symptoms which may account for the differences observed.

Symptom pattern 2 (characterised by higher symptom burden) was found to be more common in females in all the COVID-19 groups. While this could be interpreted as females having a higher (true) underlying symptom burden than males in each of these groups, an alternative explanation could be differential reporting of (ostensibly similar) symptoms between males and females. Unlike in studies of health care use, differential health-seeking behaviour is unlikely to provide an explanation as symptom information was requested of all participants in each study. Decrease in symptom burden has been previously observed in older age groups compared with younger (e.g. (32)), reflecting our results here. The identified symptom patterns among individuals with COVID-19 > 12 weeks ago were found to be strongly associated with length of time unable to function as normal due to COVID-19 symptoms. This shows that the symptom pattern identified by the LCA relates closely to long COVID.

There are many strengths to our work. Working across multiple studies with different geographic and demographic characteristics allowed us to compare findings and draw more robust conclusions. Co-ordinated standardised analyses minimised methodological heterogeneity and maximised comparability, while appropriately accounting for study designs and characteristics of individual datasets. Meta-analyses of key study-specific estimates maximised statistical power and representativeness. Focussing on functional limitation due to COVID-19 symptoms in order to identify long COVID led to small numbers which caused analytical problems, but we successfully overcame this through a novel application of LCA which allowed us to identify symptoms characteristic of long COVID (as well as acute COVID-19). This was only possible due to the inclusion of a control group of COVID-19-free individuals, which has been a limitation of previous research (15).

There are also some limitations. Although working across multiple studies has its benefits, between-study variability in structure and data availability (particularly which symptoms were reported, how and when) added considerable complexity to the analysis. While each study had a reasonable total analytic sample size, as analyses were conducted separately in COVID-19 groups, small sample sizes, particularly in the groups with COVID-19, may have affected analyses, in particular our ability to identify symptom patterns of low prevalence. This may have been exacerbated by the relatively limited number of symptoms enquired about in some studies, reflected in the core symptom set considered, though analyses using additional symptoms were possible in a smaller number of studies. Further partitioning of the ‘COVID-19 in last 12 weeks’ group into those who had had COVID-19 in the last 4 weeks and those who had had COVID-19 4 to 11 weeks ago would have allowed separate analyses relating to acute COVID-19 and ongoing symptomatic COVID-19, but this was not possible due to low numbers of relevant participants. Because we relied on self-reported COVID-19 status, individuals who had asymptomatic COVID-19 or who had COVID-19 which was misattributed to another cause may have been incorrectly classified as never having had COVID-19. If these misclassified true COVID-19 cases had COVID-19-related symptoms when we observed them, this could attenuate the differences in symptoms between the COVID-19 and no COVID-19 groups, making our findings conservative. We carried out complete-case analyses and were only able to apply non-response weights in studies where these were available. In the remaining studies (and potentially to some extent even in studies with non-response weights due to residual bias), if individuals with more debilitating symptoms were more/less likely to respond to a questionnaire, we would over/under-estimate the prevalence of symptoms. However, unless this happened differentially with respect to COVID-19 status, this would not bias our estimates of differences between COVID-19 groups. Vaccination status was not considered – and given the timing of symptom data collection relative to the vaccination programme rollout would not have been relevant for many studies – but could be of interest in future research. Finally, because data were collected prior to the emergence and dominance of the Omicron variant in the UK, findings may not be generalisable to the current UK circumstances (39).

## Conclusions

Across nine UK longitudinal studies we identified patterns of symptoms in individuals with and without COVID-19 which allowed us to discern symptoms characteristic of acute COVID-19 and long COVID. The symptoms we identified largely replicated those previously identified in the literature. Building the evidence base regarding typical long COVID symptoms will improve diagnosis of this condition and the ability to elicit underlying biological mechanisms, leading to better patient access to treatment and services.

## Supporting information

TableS4. Individual symptom analyses

## Data Availability

Data availability varies depending on the study, and are listed by study in supplementary material TableS2

## FUNDING

This work was supported by the National Core Studies, an initiative funded by UKRI, NIHR and the Health and Safety Executive. The COVID-19 Longitudinal Health and Wellbeing National Core Study was funded by the Medical Research Council (MC_PC_20059). Characterisation, determinants, mechanisms and consequences of the long-term effects of COVID-19: providing the evidence base for health care services (CONVALESCENCE) study is funded by NIHR (COV-LT-0009) (MC_PC_20051).

Data gathered from questionnaire(s) was provided by Wellcome Longitudinal Population Study (LPS) COVID-19 Steering Group and Secretariat (221574/Z/20/Z).

The 1946 National Child Development Study, the 1970 British Cohort Study, Next Steps and the Millennium Cohort Study are supported by the Centre for Longitudinal Studies Resource Centre 2015-20 grant (ES/M001660/1) and a host of other co-funders. The COVID-19 data collections in these five cohorts were funded by the UKRI grant Understanding the economic, social and health impacts of COVID-19 using lifetime data: evidence from 5 nationally representative UK cohorts (ES/V012789/1).

Born in Bradford (BiB) receives core infrastructure funding from the Wellcome Trust (WT101597MA), and a joint grant from the UK Medical Research Council (MRC) and UK Economic and Social Science Research Council (ESRC) (MR/N024397/1),the British Heart Foundation (BHF) (CS/16/4/32482), and The Health Foundation COVID-19 award (2301201). The National Institute for Health Research Yorkshire and Humber Applied Research Collaboration (ARC) (NIHR200166), and Clinical Research Network both provide support for BiB research. Born in Bradford is only possible because of the enthusiasm and commitment of the children and parents in BiB. We are grateful to all the participants, health professionals, schools and researchers who have made Born in Bradford happen.

The UK Medical Research Council and Wellcome (Grant Ref: 217065/Z/19/Z) and the University of Bristol provide core support for ALSPAC. A comprehensive list of grants funding is available on the ALSPAC website (http://www.bristol.ac.uk/alspac/external/documents/grant-acknowledgements.pdf). We are extremely grateful to all the families who took part in this study, the midwives for their help in recruiting them, and the whole ALSPAC team, which includes interviewers, computer and laboratory technicians, clerical workers, research scientists, volunteers, managers, receptionists and nurses. Please note that the study website contains details of all the data that is available through a fully searchable data dictionary and variable search tool” and reference the following webpage: http://www.bristol.ac.uk/alspac/researchers/our-data/. Ethical approval for the study was obtained from the ALSPAC Ethics and Law Committee and the Local Research Ethics Committees. Part of this data was collected using REDCap, see the REDCap website for details https://projectredcap.org/resources/citations/

Generation Scotland received core support from the Chief Scientist Office of the Scottish Government Health Directorates [CZD/16/6] and the Scottish Funding Council [HR03006]. Genotyping of the GS:SFHS samples was carried out by the Genetics Core Laboratory at the Wellcome Trust Clinical Research Facility, Edinburgh, Scotland and was funded by the Medical Research Council UK and the Wellcome Trust (Wellcome Trust Strategic Award “STratifying Resilience and Depression Longitudinally” (STRADL) Reference 104036/Z/14/Z). Generation Scotland is funded by the Wellcome Trust (216767/Z/19/Z) and (221574/Z/20/Z).

Understanding Society is an initiative funded by the Economic and Social Research Council and various Government Departments, with scientific leadership by the Institute for Social and Economic Research, University of Essex, and survey delivery by NatCen Social Research and Kantar Public. The Understanding Society COVID-19 study is funded by the Economic and Social Research Council (ES/K005146/1) and the Health Foundation (2076161). The research data are distributed by the UK Data Service.

TwinsUK receives funding from the Wellcome Trust (WT212904/Z/18/Z), the National Institute for Health Research (NIHR) Biomedical Research Centre based at Guy’s and St Thomas’ NHS Foundation Trust and King’s College London. The TwinsUK COVID-19 personal experience study was funded by the King’s Together Rapid COVID-19 Call award, under the projects original title ‘Keeping together through coronavirus: The physical and mental health implications of self-isolation due to the Covid-19 TwinsUK is also supported by the Chronic Disease Research Foundation and Zoe Global Ltd. The funders had no role in study design, data collection and analysis, decision to publish, or preparation of the manuscript.

NJT is a Wellcome Trust Investigator (202802/Z/16/Z), is the PI of the Avon Longitudinal Study of Parents and Children (MRC & WT 217065/Z/19/Z), is supported by the University of Bristol NIHR Biomedical Research Centre, the MRC Integrative Epidemiology Unit (MC_UU_00011/1) and works within the CRUK Integrative Cancer Epidemiology Programme (C18281/A29019). ASFK acknowledges funding from the ESRC (ES/V011650/1).

GBP acknowledges funding from the Economic and Social Research Council (ES/V012789/1). KT works in a Unit that is supported by the University of Bristol and UK Medical Research Council (MC_UU_00011/3). DMW is supported by funding from UK Medical Research Council (MC_PC_20030). NC is supported by funding from the UK Medical Research Council (MC_UU_00019/2).

## DATA AVAILABILITY

Data access for NCDS (SN 6137), BCS70 (SN 8547), Next Steps (SN 5545), MCS (SN 8682) and all four COVID-19 surveys (SN 8658) can be obtained through the UK Data Service. ALSPAC data is available to researchers through an online proposal system. Information regarding access can be found on the ALSPAC website (http://www.bristol.ac.uk/media-library/sites/alspac/documents/researchers/data-access/ALSPAC_Access_Policy.pdf). Data from the various Born in Braford family studies are available to researchers; see the study website for information on how to access data (https://borninbradford.nhs.uk/research/how-to-access-data/). Generation Scotland data are available through the UK Data Service (SN 6614 and SN 8644). Access to data is approved by the Generation Scotland Access Committee. See https://www.ed.ac.uk/generation-scotland/for-researchers/access or for further details. TwinsUK data are available on request from the TwinsUK Resource Executive Committee (TREC). Access to TwinsUK data can be obtained via a standard application procedure. Data requests should be submitted via https://twinsuk.ac.uk/resources-for-researchers/access-our-data/.

## Footnotes

1 We included both the ALSPAC cohort (ALSPAC-G1; born 1991-92) and their parents (ALSPAC-G0) as a pooled sample.

2 Analyses were not possible for either group with COVID-19 in BiB due to insufficient sample size.

3 With the exception of GS and USoc which lack a no COVID-19 group to act as a comparator.

4 Analyses were again not possible for either group with COVID-19 in BiB due to insufficient sample size.

