## Supplementary Material for "Characterising patterns of COVID-19 and long COVID symptoms: Evidence from nine UK longitudinal studies"

### **Methods S1**: Study-specific methods details.

**1958 National Child Development Study (NCDS), 1970 British Cohort Study (BCS70), Next Steps (NS) and Millennium Cohort Study (MCS)**

Note: These studies are described together as a common questionnaire was issued to all of them and an identical approach to variable derivation used.

Data

COVID-19 and symptoms data come from the CLS COVID-19 Survey wave 3, which was conducted between February and March 2021.

Variables

Symptoms were reported for the two-week period prior to the questionnaire being completed. Presence and date of COVID-19 was reported retrospectively, with dates between February 2020 and March 2021. Sex was recorded at previous cohort waves. The CLS cohorts are age-homogeneous, so age was not considered in these analyses. Functional limitation following COVID-19 was obtained from the question “For how long were you unable to function as normal due to Coronavirus symptoms?”

Analysis

As symptoms data were only available at a single timepoint, a cross-sectional LCA was undertaken. Adjustment was made for calendar period of symptom reporting using a binary month variable (February 2021 vs. March 2021).

**Avon Longitudinal Study of Parents and Children (ALSPAC)**

Data

COVID-19 and symptoms data are from the wave 4 data collection, which was conducted in March - April 2021.

Variables

COVID-19 symptoms data reported for the most recent month in wave 4 data are chosen for analyses. Presence and month of COVID-19 infection was directly asked in the wave 4 questionnaire and additionally sourced from retrospective wave’s data and concurrent laboratory test results. Age and sex data of the cohorts were accessed from previously existing data sources of the cohort.

Analysis

A cross sectional LCA of the longitudinal reduced data set is undertaken for the most recent reported month’s symptoms data. Adjustments were done for age and sex in the logistic models of each symptom.

**TwinsUK**

Data

COVID-19 and symptoms data come from the COVID-19 Personal Experience Survey Wave 2 (July–August 2020), wave 3 (September–December 2020) and wave 4 (April–June 2021).

Variables

Symptoms were reported for two month periods prior to questionnaire being completed. Presence, date and duration of COVID-19 incidence were reported retrospectively. Sex was recorded at previous waves. Age was calculated as difference between year of symptom reporting and year of birth, reported in previous data collection. Functional limitation was assessed from the question *For how long were you unable to function as normal due to COVID-19 symptoms?* Collected at each wave. A dummy variable was created to account for family relatedness, where identical twins were grouped to the same unique identifier, and dizygotic twins were given singular identifiers.

Analysis

For each wave, the symptoms reported for the 2 months preceding the response date were used. In LCA analysis, family relatedness was included as a cluster variable, and a set of three binary dummy variables reflecting the wave of reporting were included as covariates. Due to study design using a mixed method of data collection (questionnaires were administered online and via post for participants requesting it) some individuals reported twice within the same wave; in these cases the earliest response within wave with complete data on COVID-19 duration was used.

**Born in Bradford (BiB)**

Data

COVID-19 and symptom data were collected in Born in Bradford COVID-19 Phase 2 survey from October to December 2020.

Variables

Symptom data were reported retrospectively. In total, 27 symptoms were asked in one question. Participants were asked about each symptom every month from March to September 2020. No functional limitation variable was available. Age and sex information were from Born in Bradford baseline survey.

Analysis

To simplify the analysis, a cross-sectional LCA was undertaken. Symptoms data used were from the last reporting period – Sep 2020. No adjustment for calendar period was made.

**Understanding Society (USoc)**

Data

COVID-19 and symptoms data come from the Understanding Society COVID-19 Study Wave 7 (January 2021), Wave 8 (March 2021) and Wave 9 (September 2021).

Variables

Participants were asked to report COVID-19 symptoms in two circumstances. First participants were asked if they had experienced symptoms that could be caused by coronavirus since they had last completed the survey. Participants responding affirmatively were then asked a list of 22 symptoms. An additional two symptoms, relating to loss of concentration and remembering things, were also asked of those who also reported having COVID-19 in the previous survey. Second, participants who responded not reported having had COVID-19 since the last interview, but had reported that they had COVID-19 in previous survey, were asked if they had recovered from COVID-19, those who had not were asked the full list of 24 symptoms. Participants who reported having had COVID-19 or not recovering from COVID-19 were asked how many weeks that they had experienced for. No symptoms data was collected for participants who did not report COVID-19, and no functional limitation information was available. Given that loss of concentration and remembering things were not asked of everybody who had COVID-19 these have been omitted from the analyses.

Analysis

For each wave, symptoms reported since the last survey were used, with survey being indicated with a dummy variable. Given that the analyses were for multiple waves and symptoms were restricted to those who reported Covid, there was not a clear theoretical or methodological basis to calculate appropriate weights for this study.

**Generation Scotland (GS)**

Data

COVID-19 and symptoms data come from the Generation Scotland CovidLife Survey Wave 3, which was conducted across February 2021.

Variables

Participants were asked if they ever had or current had COVID-19. Those who answered that they had or had ever had COVID-19 (included suspected cases not confirmed by test) then reported what symptoms they had experienced. This included symptoms at any time point. No symptoms data was collected for participants who reported no COVID-19, no functional limitation variable was available, and participants did not report when symptoms occurred. Participants were asked how long they experienced COVID-19 symptoms. Sex and age was recorded as Wave 1 of the CovidLife participants.

Analysis
As symptoms were only available at a single time-point, a cross-sectional LCA was undertaken. No adjustment for calendar period was made, as data were all collected during February 2021. Analyses were restricted only to CovidLife participants who were Generation Scotland cohort members. Final class was taken based on acute COVID-19 models for the participants with symptoms less than 12 weeks, and long COVID models for the participants with symptoms lasting more than 12 weeks. Dates of infection were only available for a small subset of the data, so analyses were mostly based on length of symptom, rather than time since infection.

### **Table S1**: Details of included studies.

| Study population | Design and sample frame | Age(s) in 2020 | Details of COVID-19 surveys (response rate) |
| --- | --- | --- | --- |
| *Age homogenous cohorts* | |  |  |
| 1958 National Child Development Study (NCDS) | Cohort of all children born in Great Britain (i.e. England, Wales & Scotland) in one week in 1958, with regular follow-up surveys from birth. | 62 | February-March 2021 (58.5%) |
| 1970 British Cohort Study (BCS70) | Cohort of all children born in Great Britain (i.e. England, Wales & Scotland) in one week in 1970, with regular follow-up surveys from birth. | 50 | February-March 2021 (45.4%) |
| Next Steps (NS) | Sample recruited via secondary schools in England at around age 13 with regular follow-up surveys thereafter. | 29-31 | February-March 2021 (34.3%) |
| Millennium Cohort Study (MCS) | Cohort of UK children born between Sept 2000 and Jan 2002 with regular follow-up surveys from birth. | 18-20 | February-March 2021 (33.1%) |
| Avon Longitudinal Study of Parents and Children (ALSPAC) - Generation 1 (G1) | Cohort of children born in the South-West of England between April 1991 and Dec 1992, with regular follow-up surveys from birth. | 27-29 | Three questionnaires: April 2020 (19%), June 2020 (17.4%), December 2020 (26.4%) |
| *Age heterogeneous studies* | |  |  |
| Avon Longitudinal Study of Parents and Children (ALSPAC) - Generation 0 (G0) | Parents of the ALSPAC (G1) cohort described above, treated as a separate age-heterogenous study population. | 45-81 | Three questionnaires: April 2020 (12.4%), June 2020 (12.2%), December 2020 (14.3%) |
| TwinsUK: the UK Adult Twin Registry | A cohort of UK volunteer adult twins (55% monozygotic and 43% dizygotic) who were sampled between 18-101 years of age. | 22-96 | Four surveys: April 2020 (64.3%), July 2020 (77.6%), November 2020 (76.1%), March 2021 (76%) |
| Born in Bradford (BiB) | Birth cohort recruiting pregnant women and their children between 2007 and 2011 | 28-55 | Two surveys: April-June 2020 (30.7%) & October-November 2020 (39.9%) |
| Understanding Society: the UK Household Longitudinal Survey (USoc) | A nationally representative longitudinal household panel study, based on a clustered-stratified probability sample of UK households, with all adults aged 16+ in chosen households surveyed annually. | 16-96 | Three surveys (full/partial interview): January 2021 (28.5%), March 2021(30.2%), September 2021(30.6%) |
| Generation Scotland: the Scottish Family Health Study (GS) | A family-structured, population-based Scottish cohort, with participants aged 18-99 recruited between 2006-2011 | 27-100 | Three surveys: April-June 2020 (21.3%), July-August 2020 (15.4%), February 2021 (14.3%) |

### **Table S2**: Ethics and data access statements for each study.

| Study | Ethics and data access statements |
| --- | --- |
| 1958 National Child Development Study (NCDS), 1970 British Cohort Study (BCS70), Next Steps (NS), Millennium Cohort Study (MCS) | The most recent sweeps of the NCDS, BCS70, Next Steps and MCS have all been granted ethical approval by the National Health Service (NHS) Research Ethics Committee and all participants have given informed consent. Data for NCDS (SN 6137), BCS70 (SN 8547), Next Steps (SN 5545), MCS (SN 8682) and all four COVID-19 surveys (SN 8658) are available through the UK Data Service. |
| Avon Longitudinal Study of Parents and Children (ALSPAC) | Ethical approval was obtained from the ALSPAC Ethics and Law Committee and the Local Research Ethics Committees. The study website contains details of all the data that is available through a fully searchable data dictionary and variable search tool: <http://www.bristol.ac.uk/alspac/researchers/our-data>. ALSPAC data is available to researchers through an online proposal system. Information regarding access can be found on the ALSPAC website (<http://www.bristol.ac.uk/media-library/sites/alspac/documents/researchers/data-access/ALSPAC_Access_Policy.pdf>). |
| TwinsUK: the UK Adult Twin Registry | All waves of TwinsUK have received ethical approval associated with TwinsUK Biobank (19/NW/0187), TwinsUK (EC04/015) or Healthy Ageing Twin Study (H.A.T.S) (07/H0802/84) studies from NHS Research Ethics Committees at the Department of Twin Research and Genetic Epidemiology, King’s College London. The TwinsUK Resource Executive Committee (TREC) oversees management, data sharing and collaborations involving the TwinsUK registry (for further details see <https://twinsuk.ac.uk/resources-for-researchers/access-our-data/>). |
| Born in Bradford (BiB) | Ethical approval for Born in Bradford was granted by the National Health Service Health Research Authority Yorkshire and the Humber (Bradford Leeds) Research Ethics Committee (reference: 16/YH/0320). Data from the various BiB family studies are available to researchers; see the study website for information on how to access data (<https://borninbradford.nhs.uk/research/how-to-access-data/>). |
| Understanding Society: the UK Household Longitudinal Survey (USoc) | The University of Essex Ethics Committee has approved all data collection for the Understanding Society main study and COVID-19 waves (ETH1920-1271). The March 2021 survey was approved by the NHS Health Authority, London – City & East East Research Committee (21/HRA/0644). No additional ethical approval was necessary for this secondary data analysis. All data are available through the UK Data Service (SN 6614 and SN 8644). |
| Generation Scotland: the Scottish Family Health Study (GS) | Generation Scotland obtained ethical approval from the East of Scotland Committee on Medical Research Ethics (on behalf of the National Health Service). Reference number 20/ES/0021. Access to data is approved by the Generation Scotland Access Committee. See <https://www.ed.ac.uk/generation-scotland/for-researchers/access> or for further details. |

### **Table S3**. Symptom sets within each study.

|  | NCDS, BCS70, NS, MCS | |  | ALSPAC | |  | TwinsUK | |  | BiB | |  | USoc | |  | GS | |
| --- | --- | --- | --- | --- | --- | --- | --- | --- | --- | --- | --- | --- | --- | --- | --- | --- | --- |
| Symptom | Core  set | Maximal  set |  | Core  set | Maximal  set |  | Core  set | Maximal  set |  | Core  set | Maximal  set |  | Core  set | Maximal  set |  | Core  set | Maximal  set |
| Fever | 1 | 1 |  | 1 | 1 |  | 1 | 1 |  | 1 | 1 |  | 1 | 1 |  | 1 | 1 |
| Cough - dry | 1 | 1 |  |  |  |  |  |  |  |  |  |  |  |  |  | 1 | 1 |
| Cough - mucus or phlegm | 1 | 1 |  |  |  |  |  |  |  |  |  |  |  |  |  |  |  |
| Persistent cough (non-specific) |  |  |  | 1 | 1 |  | 1 | 1 |  | 1 | 1 |  | 1 | 1 |  |  |  |
| Chesty cough |  |  |  |  |  |  | 1 | 1 |  |  |  |  |  |  |  |  |  |
| Sore throat | 1 | 1 |  | 1 | 1 |  | 1 | 1 |  | 1 | 1 |  | 1 | 1 |  | 1 | 1 |
| Chest tightness | 1 | 1 |  | 1 | 1 |  | 1 | 1 |  | 1 | 1 |  | 1 | 1 |  |  |  |
| Shortness of breath | 1 | 1 |  | 1 | 1 |  | 1 | 1 |  | 1 | 1 |  | 1 | 1 |  | 1 | 1 |
| Runny nose | 1 | 1 |  | 1 | 1 |  | 1 | 1 |  | 1 | 1 |  |  |  |  | 1 | 1 |
| Blocked nose | 1 | 1 |  | 1 | 1 |  | 1 | 1 |  | 1 | 1 |  |  |  |  | 1 | 1 |
| Runny or stuffy nose |  |  |  |  |  |  |  |  |  |  |  |  | 1 | 1 |  |  |  |
| Muscle or body aches | 1 | 1 |  | 1 | 1 |  | 1 | 1 |  | 1 | 1 |  | 1 | 1 |  | 1 | 1 |
| Fatigue | 1 | 1 |  | 1 | 1 |  | 1 | 1 |  |  |  |  | 1 | 1 |  | 1 | 1 |
| Severe fatigue |  |  |  |  | 1 |  |  | 1 |  |  | 1 |  |  |  |  |  |  |
| Diarrhoea | 1 | 1 |  | 1 | 1 |  | 1 | 1 |  | 1 | 1 |  | 1 | 1 |  | 1 | 1 |
| Loss of smell | 1 | 1 |  |  |  |  | 1 | 1 |  |  |  |  |  |  |  |  |  |
| Loss of taste | 1 | 1 |  |  |  |  | 1 | 1 |  |  |  |  |  |  |  |  |  |
| Loss of smell or taste |  |  |  | 1 | 1 |  |  |  |  | 1 | 1 |  | 1 | 1 |  | 1 | 1 |
| Nausea and/or vomiting | 1 | 1 |  | 1 | 1 |  | 1 | 1 |  | 1 | 1 |  |  |  |  | 1 | 1 |
| Raised, red, itchy areas on the skin | 1 | 1 |  | 1 | 1 |  |  |  |  | 1 | 1 |  |  |  |  |  |  |
| Sneezing | 1 | 1 |  | 1 | 1 |  | 1 | 1 |  | 1 | 1 |  | 1 | 1 |  |  |  |
| Headaches | 1 | 1 |  | 1 | 1 |  | 1 | 1 |  | 1 | 1 |  | 1 | 1 |  | 1 | 1 |
| Decrease in appetite |  |  |  |  | 1 |  |  | 1 |  |  | 1 |  |  | 1 |  |  | 1 |
| Abdominal pain/tummy ache |  |  |  |  | 1 |  |  | 1 |  |  | 1 |  |  |  |  |  | 1 |
| Sore eyes |  |  |  |  | 1 |  |  | 1 |  |  | 1 |  |  | 1 |  |  | 1 |
| Hoarse voice |  |  |  |  | 1 |  |  | 1 |  |  | 1 |  |  | 1 |  |  |  |
| Dizziness |  |  |  |  | 1 |  |  | 1 |  |  | 1 |  |  | 1 |  |  |  |
| Chest pain |  |  |  |  | 1 |  |  | 1 |  |  | 1 |  |  | 1 |  |  |  |
| Chills |  |  |  |  | 1 |  |  | 1 |  |  | 1 |  |  | 1 |  |  |  |
| Difficulty sleeping |  |  |  |  | 1 |  |  | 1 |  |  | 1 |  |  | 1 |  |  |  |
| Numbness or tingling somewhere in the body |  |  |  |  | 1 |  |  | 1 |  |  | 1 |  |  | 1 |  |  |  |
| Feeling of heaviness in arms or legs |  |  |  |  | 1 |  |  | 1 |  |  | 1 |  |  | 1 |  |  |  |
| Sudden swelling of the face or lips |  |  |  |  | 1 |  |  | 1 |  |  | 1 |  |  |  |  |  |  |
| Difficulty concentrating | 1 | 1 |  |  |  |  | 1 | 1 |  |  |  |  |  |  |  | 1 | 1 |
| Memory loss | 1 | 1 |  |  |  |  | 1 | 1 |  |  |  |  |  |  |  |  |  |
| **Total** | **19** | **19** |  | **15** | **27** |  | **18** | **30** |  | **14** | **26** |  | **12** | **21** |  | **13** | **16** |

NCDS: 1958 National Child Development Study; BCS1970: 1970 British Cohort Study; NS: Next Steps; MCS: Millennium Cohort Study; ALSPAC: Avon Longitudinal Study of Parents and Children; BiB: Born in Bradford; USoc Understanding Society; GS: Generation Scotland. Colour coding represents symptoms relating to a common or overlapping concept.

### **Table S4**. Individual symptom analyses in each study.

See separate file: [Individual symptom analyses.xlsx](https://liveuclac.sharepoint.com/:x:/r/sites/LHWStudyGroup/Shared%20Documents/Long%20Covid%20(ARQ2)/CONVALESCENCE%20-%20Work%20Package%201%20-%20Definition/Symptom%20clustering/Manuscript/Individual%20symptom%20analyses.xlsx?d=w3f76fb2dca0e41b5ac75a018041bb87a&csf=1&web=1&e=iPfv8b)

FL: Functional limitation. Small cells counts suppressed if <5 or <10 depending on study policy.

### **Table S5**. Individual symptom analyses meta-analysed across studies.

|  | COVID-19 in last 12 weeks  vs. No COVID-19 | | | | | |  | COVID-19 > 12 weeks ago + no FL at 12 weeks  vs. No COVID-19 | | | | | |  | COVID-19 > 12 weeks ago + FL at 12 weeks  vs. No COVID-19 | | | | | |
| --- | --- | --- | --- | --- | --- | --- | --- | --- | --- | --- | --- | --- | --- | --- | --- | --- | --- | --- | --- | --- |
| Symptom | OR | OR 95%  CI lower | OR 95%  CI upper | I^2^ | Q | No.  studies |  | OR | OR 95%  CI lower | OR 95%  CI upper | I^2^ | Q | No.  studies |  | OR | OR 95%  CI lower | OR 95%  CI upper | I^2^ | Q | No.  studies |
| Fever | 5.5 | 4.3 | 7.1 | 0.0 | 4.6 | 6 |  | 1.5 | 1.1 | 2.2 | 7.9 | 4.3 | 5 |  | 5.9 | 1.4 | 24.6 | 53.2 | 4.3 | 3 |
| Cough | 3.6 | 2.0 | 6.3 | 83.6 | 30.4 | 6 |  | 1.5 | 1.1 | 2.0 | 45.0 | 7.3 | 5 |  | 5.6 | 2.9 | 10.8 | 0.0 | 3.7 | 5 |
| Sore throat | 2.0 | 1.5 | 2.7 | 53.5 | 12.9 | 7 |  | 1.2 | 1.0 | 1.4 | 22.0 | 5.1 | 5 |  | 3.0 | 1.3 | 7.1 | 45.2 | 7.3 | 5 |
| Chest tightness | 2.6 | 2.1 | 3.2 | 0.0 | 2.8 | 6 |  | 1.8 | 1.5 | 2.2 | 0.0 | 2.7 | 5 |  | 7.3 | 4.3 | 12.3 | 11.8 | 4.5 | 5 |
| Shortness of breath | 3.1 | 2.6 | 3.8 | 0.0 | 3.4 | 6 |  | 2.0 | 1.6 | 2.4 | 0.0 | 1.5 | 5 |  | 11.9 | 5.3 | 26.6 | 63.4 | 10.9 | 5 |
| Runny nose | 1.1 | 0.9 | 1.4 | 35.0 | 9.2 | 7 |  | 1.1 | 0.9 | 1.2 | 0.0 | 3.5 | 5 |  | 1.4 | 0.8 | 2.5 | 0.0 | 2.8 | 5 |
| Muscle pain or aches | 2.4 | 2.1 | 2.9 | 0.0 | 4.8 | 6 |  | 1.6 | 1.4 | 1.9 | 0.0 | 1.6 | 5 |  | 9.7 | 6.0 | 15.8 | 0.0 | 3.0 | 5 |
| Fatigue | 2.0 | 1.3 | 3.1 | 88.8 | 53.7 | 7 |  | 1.7 | 1.3 | 2.2 | 75.2 | 16.1 | 5 |  | 13.7 | 6.9 | 27.3 | 47.4 | 7.6 | 5 |
| Diarrhoea | 1.8 | 1.3 | 2.4 | 47.6 | 11.4 | 7 |  | 1.5 | 1.0 | 2.2 | 69.7 | 13.2 | 5 |  | 1.8 | 0.9 | 3.3 | 22.7 | 5.2 | 5 |
| Loss of smell or taste | 30.9 | 16.5 | 57.9 | 0.0 | 0.0 | 1 |  |  |  |  |  |  |  |  |  |  |  |  |  |  |
| Loss of smell | 28.6 | 16.6 | 49.2 | 58.2 | 9.6 | 5 |  | 6.8 | 4.4 | 10.5 | 38.9 | 6.5 | 5 |  | 26.3 | 7.7 | 89.2 | 64.3 | 11.2 | 5 |
| Loss of taste | 20.5 | 15.4 | 27.4 | 0.0 | 2.7 | 5 |  | 4.2 | 3.1 | 5.8 | 0.0 | 1.1 | 5 |  | 33.1 | 9.8 | 111.5 | 59.8 | 10.0 | 5 |
| Nausea and/or vomiting | 1.5 | 1.0 | 2.4 | 50.8 | 12.2 | 7 |  | 0.7 | 0.4 | 1.3 | 63.3 | 10.9 | 5 |  | 2.1 | 0.8 | 5.7 | 9.7 | 2.2 | 3 |
| Rash/itchy skin | 1.6 | 0.9 | 2.9 | 67.5 | 18.4 | 7 |  | 1.2 | 0.7 | 2.0 | 72.2 | 14.4 | 5 |  | 2.2 | 1.0 | 5.0 | 0.0 | 3.5 | 5 |
| Sneezing | 1.2 | 1.0 | 1.4 | 28.3 | 8.4 | 7 |  | 1.0 | 0.9 | 1.1 | 0.0 | 2.5 | 5 |  | 0.8 | 0.4 | 1.4 | 11.8 | 4.5 | 5 |
| Headaches | 1.4 | 0.9 | 2.2 | 88.2 | 51.0 | 7 |  | 0.9 | 0.7 | 1.2 | 72.1 | 14.4 | 5 |  | 3.4 | 2.1 | 5.6 | 0.0 | 0.5 | 5 |
| Difficulty concentrating | 2.5 | 1.1 | 5.8 | 91.1 | 33.8 | 4 |  | 1.9 | 1.3 | 2.7 | 68.4 | 9.5 | 4 |  | 6.2 | 1.4 | 28.0 | 84.3 | 19.1 | 4 |
| Memory loss | 1.5 | 0.6 | 3.7 | 91.2 | 34.1 | 4 |  | 1.5 | 1.1 | 2.1 | 60.0 | 7.5 | 4 |  | 6.3 | 3.1 | 13.0 | 24.9 | 4.0 | 4 |

### **Table S6**. Latent class analysis model fit statistics by COVID-19 status in each study (core symptom set).

#### **Table S6A**. 1958 National Child Development Study.

|  | Classes | | | | |  |
| --- | --- | --- | --- | --- | --- | --- |
|  | 1 | 2 | 3 | 4 | 5 | IC plot |
| No COVID-19 (n = 6,005) | | | | | | 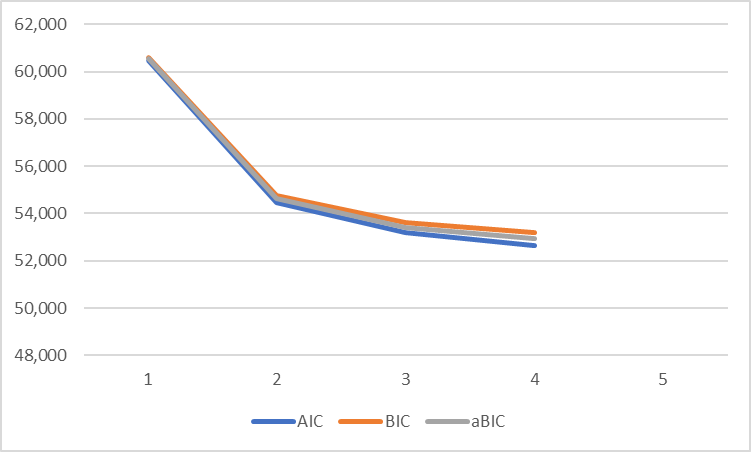 |
| No. parameters | 19 | 40 | 61 | 82 | - |  |
| Log likelihood | -30,217 | -27,201 | -26.535 | -26,236 | - |  |
| AIC | 60,471 | 54,482 | 53,193 | 52,637 | - |  |
| BIC | 60,599 | 54,750 | 53,601 | 53,186 | - |  |
| aBIC | 60,538 | 54,623 | 53408 | 52,926 | - |  |
| Smallest class % | n/a | 24.8 | 8.7 | 5.6 | - |  |
| Entropy | n/a | 0.79 | 0.74 | 0.74 | - |  |
| COVID-19 in last 12 weeks (n = 203) | | | | | | 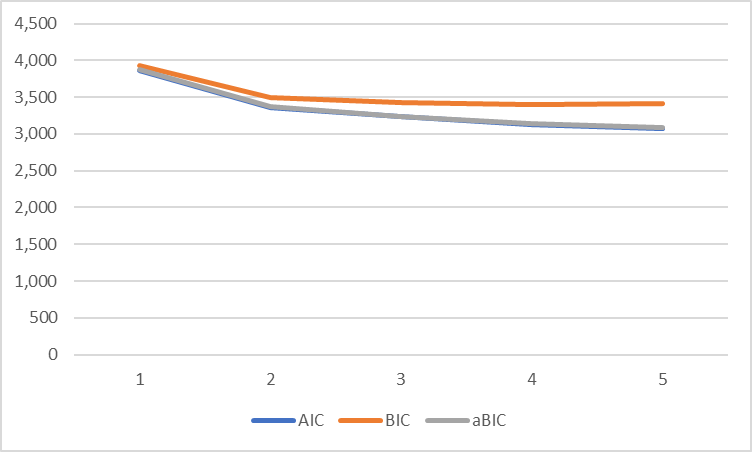 |
| No. parameters | 19 | 40 | 61 | 82 | 103 |  |
| Log likelihood | -1,915 | -1,641 | -1,554 | -1,485 | -1,435 |  |
| AIC | 3,867 | 3,363 | 3,231 | 3,134 | 3,077 |  |
| BIC | 3,930 | 3,495 | 3,433 | 3,406 | 3,418 |  |
| aBIC | 3,870 | 3,368 | 3,239 | 3,146 | 3,092 |  |
| Smallest class % | n/a | 39.0 | 3.4 | 3.3 | 3.3 |  |
| Entropy | n/a | 0.90 | 0.92 | 0.93 | 0.90 |  |
| COVID-19 > 12 weeks ago (n = 503) | | | | | | 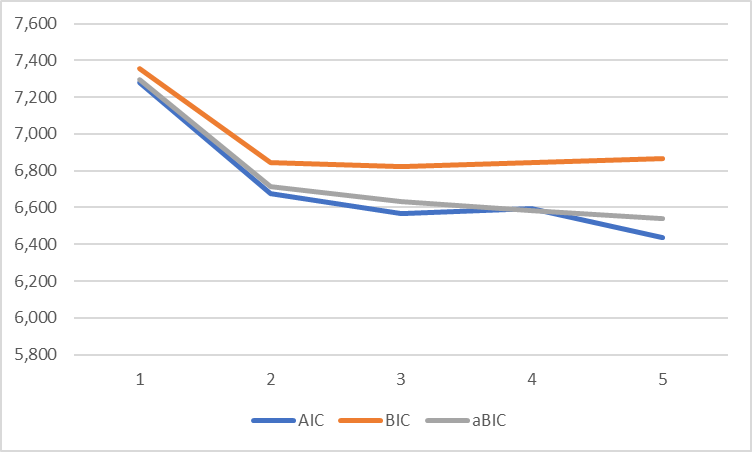 |
| No. parameters | 19 | 40 | 61 | 82 | 103 |  |
| Log likelihood | -3,620 | -3,297 | -3,222 | -3,166 | -3,114 |  |
| AIC | 7,278 | 6,674 | 6,566 | 6,595 | 6,435 |  |
| BIC | 7,358 | 6,843 | 6,824 | 6,842 | 6,869 |  |
| aBIC | 7,297 | 6,716 | 6,630 | 6,581 | 6,542 |  |
| Smallest class % | n/a | 33.3 | 3.3 | 2.9 | 2.9 |  |
| Entropy | n/a | 0.81 | 0.88 | 0.81 | 0.80 |  |

AIC: Akaike information criterion; BIC: Bayesian information criterion; aBIC: Sample size-adjusted Bayesian information criterion; n/a: not applicable.

#### **Table S6B**. 1970 British Cohort Study.

|  | Classes | | | | |  |
| --- | --- | --- | --- | --- | --- | --- |
|  | 1 | 2 | 3 | 4 | 5 | IC plot |
| No COVID-19 (n = 4,746) | | | | | | 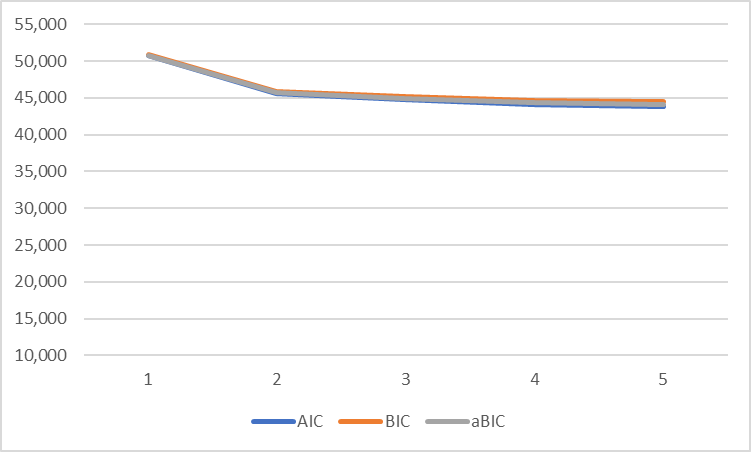 |
| No. parameters | 19 | 40 | 61 | 82 | 103 |  |
| Log likelihood | -25,347 | -22,778 | -22,303 | -21,988 | -21,807 |  |
| AIC | 50,732 | 45,636 | 44,728 | 44,140 | 43,819 |  |
| BIC | 50,855 | 45,895 | 45,123 | 44,670 | 44,485 |  |
| aBIC | 50,794 | 45,767 | 44,929 | 44,410 | 44,158 |  |
| Smallest class % | n/a | 28.2 | 6.8 | 5.9 | 0.7 |  |
| Entropy | n/a | 0.78 | 0.76 | 0.74 | 0.77 |  |
| COVID-19 in last 12 weeks (n = 266) | | | | | | 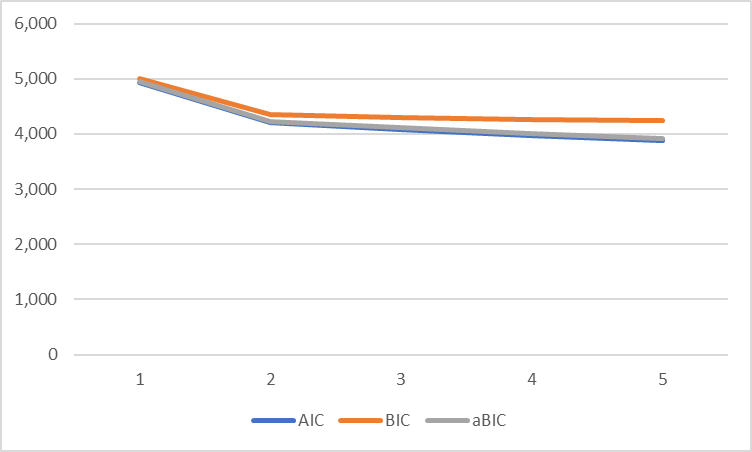 |
| No. parameters | 19 | 40 | 61 | 82 | 103 |  |
| Log likelihood | -2,448 | -2,064 | -1,983 | -1,906 | -1,838 |  |
| AIC | 4,934 | 4,207 | 4,087 | 3,977 | 3,881 |  |
| BIC | 5,002 | 4,350 | 4,306 | 4,270 | 4,251 |  |
| aBIC | 4,942 | 4,224 | 4,112 | 4,010 | 3,924 |  |
| Smallest class % | n/a | 41.5 | 28.2 | 7.8 | 3.9 |  |
| Entropy | n/a | 0.91 | 0.88 | 0.92 | 0.93 |  |
| COVID-19 > 12 weeks ago (n = 618) | | | | | | 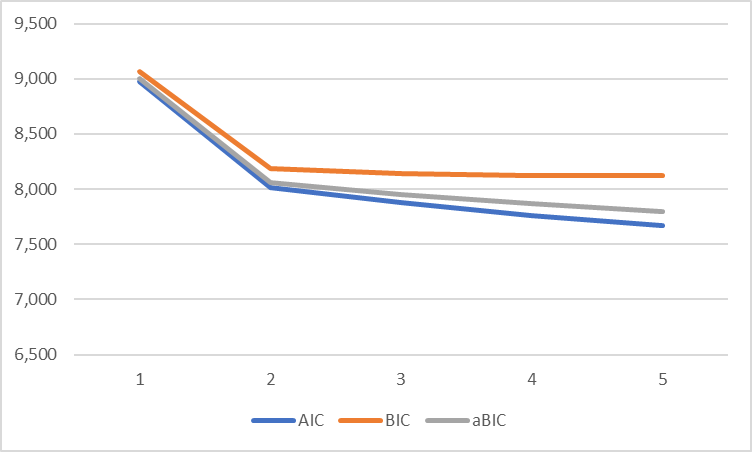 |
| No. parameters | 19 | 40 | 61 | 82 | 103 |  |
| Log likelihood | -4,470 | -3,965 | -3,876 | -3,800 | -3,732 |  |
| AIC | 8,979 | 8,010 | 7,874 | 7,765 | 7,669 |  |
| BIC | 9,063 | 8,187 | 8,144 | 8,127 | 8,125 |  |
| aBIC | 9,003 | 8,060 | 7,950 | 7,867 | 7,798 |  |
| Smallest class % | n/a | 39.4 | 15.4 | 11.3 | 1.6 |  |
| Entropy | n/a | 0.84 | 0.84 | 0.84 | 0.90 |  |

AIC: Akaike information criterion; BIC: Bayesian information criterion; aBIC: Sample size-adjusted Bayesian information criterion; n/a: not applicable.

#### **Table S6C**. Next Steps.

|  | Classes | | | | |  |
| --- | --- | --- | --- | --- | --- | --- |
|  | 1 | 2 | 3 | 4 | 5 | IC plot |
| No COVID-19 (n = 3,263) | | | | | | 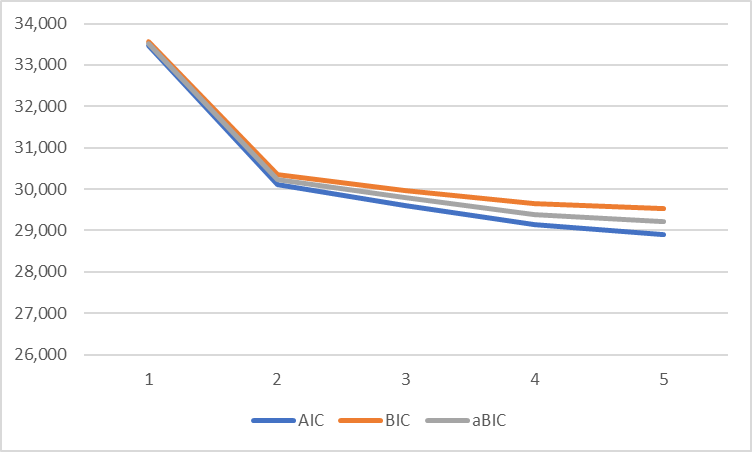 |
| No. parameters | 19 | 40 | 61 | 82 | 103 |  |
| Log likelihood | -16,711 | -15,016 | -14,742 | -14,496 | -14,354 |  |
| AIC | 33,461 | 30,111 | 29,607 | 29,156 | 28,914 |  |
| BIC | 33,577 | 30,355 | 29,978 | 29,656 | 29,542 |  |
| aBIC | 33,516 | 30,228 | 29,785 | 29,395 | 29,214 |  |
| Smallest class % | n/a | 26.7 | 13.4 | 7.5 | 6.7 |  |
| Entropy | n/a | 0.79 | 0.76 | 0.74 | 0.75 |  |
| COVID-19 in last 12 weeks (n = 260) | | | | | | 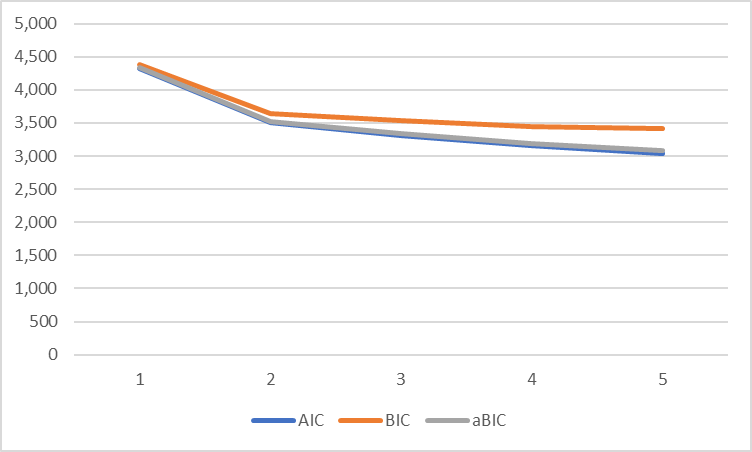 |
| No. parameters | 19 | 40 | 61 | 82 | 103 |  |
| Log likelihood | -2,142 | -1,711 | -1,597 | -1,495 | -1,418 |  |
| AIC | 4,322 | 3,502 | 3,316 | 3,155 | 3,041 |  |
| BIC | 4,389 | 3,645 | 3,533 | 3,447 | 3,408 |  |
| aBIC | 4,329 | 3,518 | 3,339 | 3,187 | 3,082 |  |
| Smallest class % | n/a | 34.2 | 9.4 | 4.6 | 4.6 |  |
| Entropy | n/a | 0.95 | 0.94 | 0.97 | 0.95 |  |
| COVID-19 > 12 weeks ago (n = 565) | | | | | | 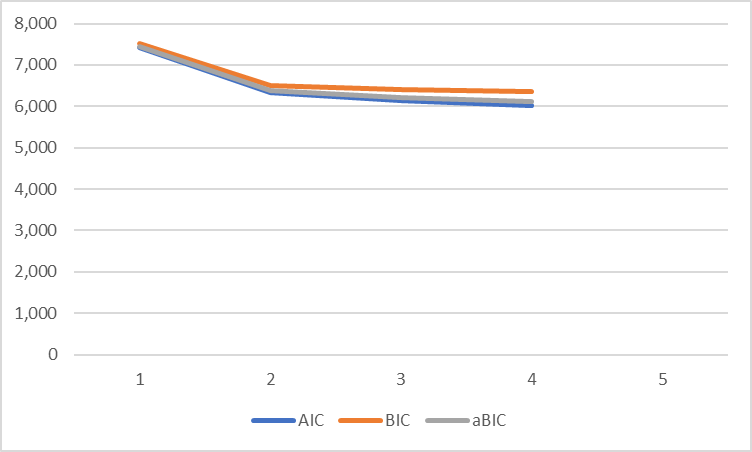 |
| No. parameters | 19 | 40 | 61 | 82 | 103 |  |
| Log likelihood | -3,694 | -3,129 | -3,012 | -2,923 | - |  |
| AIC | 7,427 | 6,338 | 6,147 | 6,010 | - |  |
| BIC | 7,509 | 6,511 | 6,411 | 6,366 | - |  |
| aBIC | 7,449 | 6,384 | 6,218 | 6,105 | - |  |
| Smallest class % | n/a | 30.5 | 2.2 | 2.3 | - |  |
| Entropy | n/a | 0.88 | 0.93 | 0.91 | - |  |

AIC: Akaike information criterion; BIC: Bayesian information criterion; aBIC: Sample size-adjusted Bayesian information criterion; n/a: not applicable.

#### **Table S6D**. Millennium Cohort Study.

|  | Classes | | | | |  |
| --- | --- | --- | --- | --- | --- | --- |
|  | 1 | 2 | 3 | 4 | 5 | IC plot |
| No COVID-19 (n = 3,283) | | | | | | 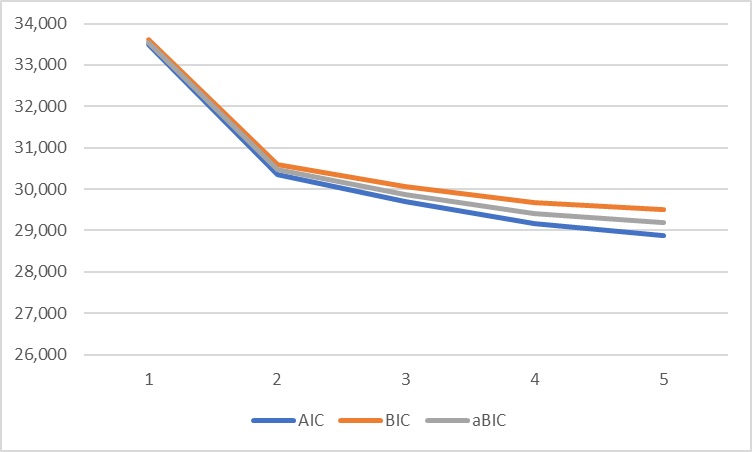 |
| No. parameters | 19 | 40 | 61 | 82 | 103 |  |
| Log likelihood | -16,730 | -15,138 | -14,790 | -14,504 | -14,342 |  |
| AIC | 33,498 | 30,357 | 29,702 | 29,172 | 28,889 |  |
| BIC | 33,613 | 30,601 | 30,074 | 29,672 | 29,517 |  |
| aBIC | 33,553 | 30,474 | 29,880 | 29,411 | 29,190 |  |
| Smallest class % | n/a | 34.0 | 19.0 | 5.7 | 0.6 |  |
| Entropy | n/a | 0.75 | 0.76 | 0.76 | 0.78 |  |
| COVID-19 in last 12 weeks (n = 254) | | | | | | 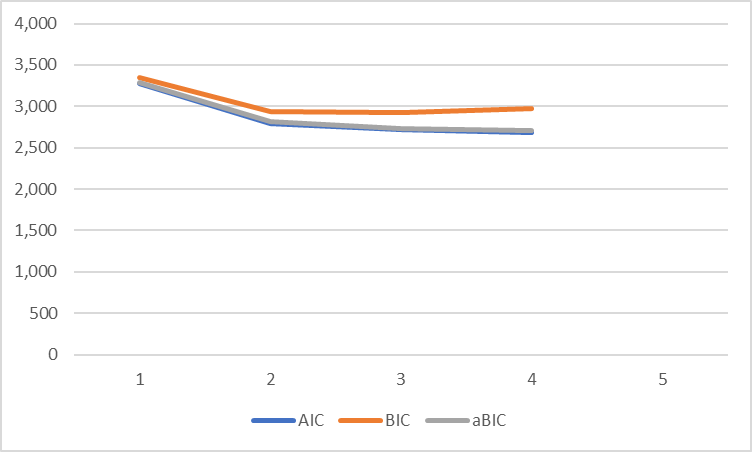 |
| No. parameters | 19 | 40 | 61 | 82 | 103 |  |
| Log likelihood | -1,620 | -1,359 | -1,296 | -1,258 | - |  |
| AIC | 3,277 | 2,797 | 2,714 | 2,681 | - |  |
| BIC | 3,345 | 2,939 | 2,930 | 2,971 | - |  |
| aBIC | 3,284 | 2,812 | 2,737 | 2,711 | - |  |
| Smallest class % | n/a | 20.7 | 8.6 | 6.5 | - |  |
| Entropy | n/a | 0.91 | 0.86 | 0.87 | - |  |
| COVID-19 > 12 weeks ago (n = 800) | | | | | | 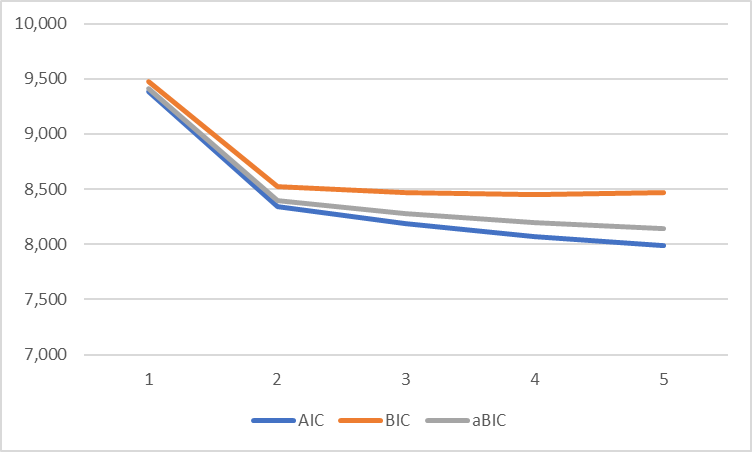 |
| No. parameters | 19 | 40 | 61 | 82 | 103 |  |
| Log likelihood | -4,674 | -4,130 | -4,031 | -3,953 | -3,890 |  |
| AIC | 9,385 | 8,339 | 8,185 | 8,069 | 7,986 |  |
| BIC | 9,474 | 8,526 | 8,470 | 8,453 | 8,469 |  |
| aBIC | 9,414 | 8,399 | 8,277 | 8,193 | 8,142 |  |
| Smallest class % | n/a | 36.8 | 3.8 | 2.1 | 2.1 |  |
| Entropy | n/a | 0.83 | 0.86 | 0.88 | 0.90 |  |

AIC: Akaike information criterion; BIC: Bayesian information criterion; aBIC: Sample size-adjusted Bayesian information criterion; n/a: not applicable.

#### **Table S6E**. Avon Longitudinal Study of Parents and Children.

|  | Classes | | | | |  |
| --- | --- | --- | --- | --- | --- | --- |
|  | 1 | 2 | 3 | 4 | 5 | IC plot |
| No COVID-19 (n = 4,877) | | | | | | 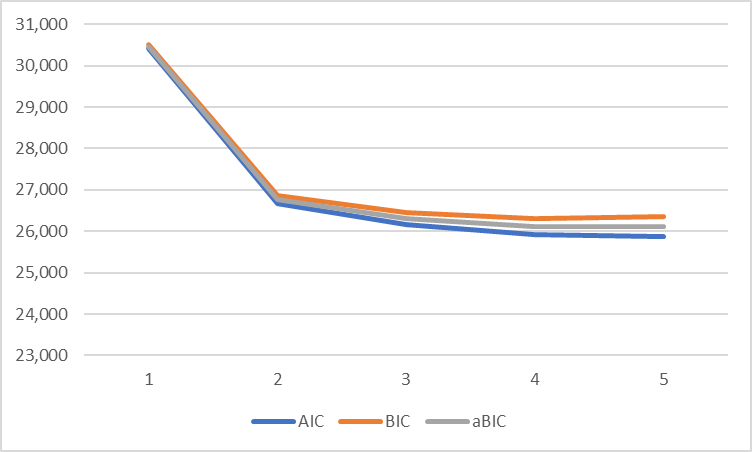 |
| No. parameters | 14 | 29 | 44 | 59 | 74 |  |
| Log likelihood | -15,193 | -13,311 | -13,036 | -12,903 | -12,867 |  |
| AIC | 30,414 | 26,680 | 26,159 | 25,924 | 25,881 |  |
| BIC | 30,505 | 26,869 | 26,445 | 26,307 | 26,361 |  |
| aBIC | 30,460 | 26,776 | 26,305 | 26,120 | 26,126 |  |
| Smallest class % | n/a | 23.6 | 10.2 | 3.7 | 1.9 |  |
| Entropy | n/a | 0.75 | 0.76 | 0.69 | 0.69 |  |
| COVID-19 in last 12 weeks (n = 181) | | | | | | 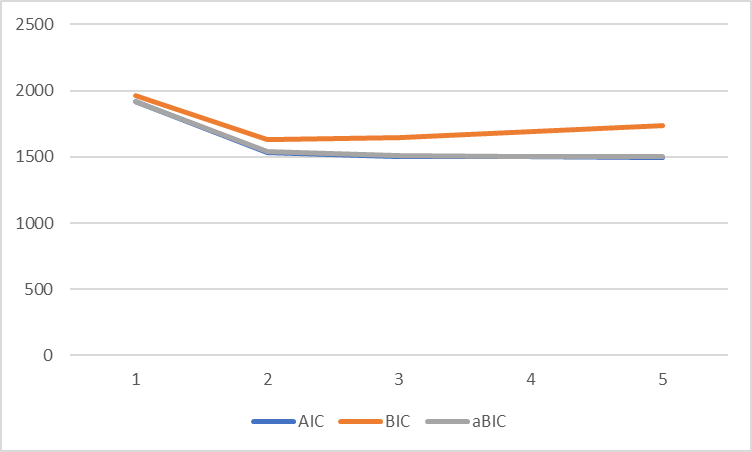 |
| No. parameters | 14 | 29 | 44 | 59 | 74 |  |
| Log likelihood | -945 | -738 | -708 | -692 | -674 |  |
| AIC | 1,918 | 1,535 | 1,504 | 1,501 | 1,497 |  |
| BIC | 1,963 | 1,628 | 1,645 | 1,690 | 1,734 |  |
| aBIC | 1,919 | 1,536 | 1,506 | 1,503 | 1,499 |  |
| Smallest class % | n/a | 21.6 | 10.5 | 7.2 | 2.8 |  |
| Entropy | n/a | 0.93 | 0.91 | 0.87 | 1.0 |  |
| COVID-19 > 12 weeks ago (n = 631) | | | | | | 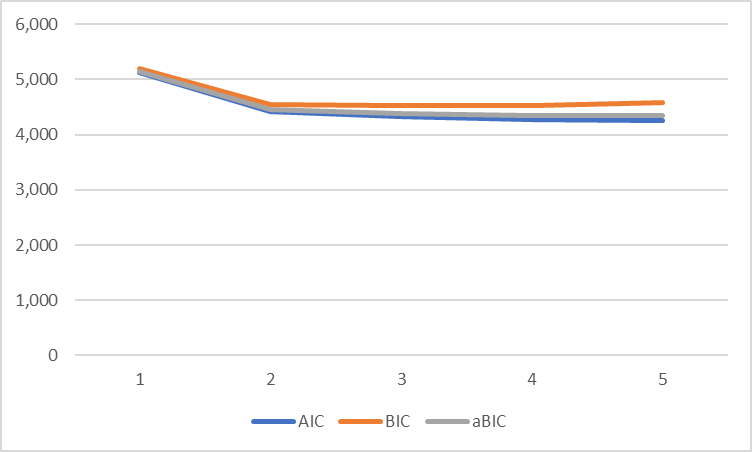 |
| No. parameters | 14 | 29 | 44 | 59 | 74 |  |
| Log likelihood | -2,552 | -2,182 | -2,120 | -2,074 | -2,056 |  |
| AIC | 5,132 | 4,422 | 4,328 | 4,266 | 4,260 |  |
| BIC | 5,195 | 4,551 | 4,524 | 4,529 | 4,589 |  |
| aBIC | 5,150 | 4,459 | 4,384 | 4,341 | 4,354 |  |
| Smallest class % | n/a | 20.9 | 3.8 | 4.1 | 1.1 |  |
| Entropy | n/a | 0.80 | 0.75 | 0.79 | 0.86 |  |

AIC: Akaike information criterion; BIC: Bayesian information criterion; aBIC: Sample size-adjusted Bayesian information criterion; n/a: not applicable.

#### **Table S6F**. TwinsUK

|  | Classes | | | | |  |
| --- | --- | --- | --- | --- | --- | --- |
|  | 1 | 2 | 3 | 4 | 5 | IC plot |
| No COVID-19 (n = 11,380) | | | | | | 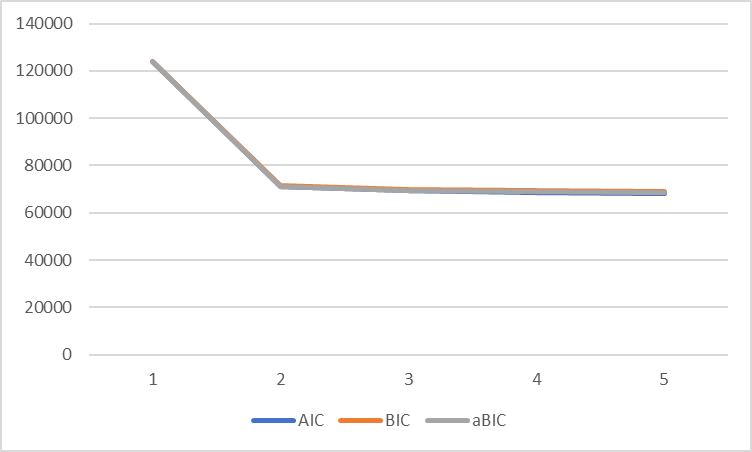 |
| No. parameters | 23 | 38 | 59 | 80 | 101 |  |
| Log likelihood | -61,955 | -35,492 | -34,552 | -34,270 | -34,031 |  |
| AIC | 123,955 | 71,060 | 69,222 | 68,700 | 68,264 |  |
| BIC | 124,124 | 71,339 | 69,655 | 69,287 | 69,005 |  |
| aBIC | 124,051 | 71,218 | 69,468 | 69,033 | 68,684 |  |
| Smallest class % | n/a | 18.65 | 0.39 | 0.38 | 0.38 |  |
| Entropy | n/a | 0.79 | 0.82 | 0.725 | 0.72 |  |
| COVID-19 in last 12 weeks (n = 652) | | | | | | 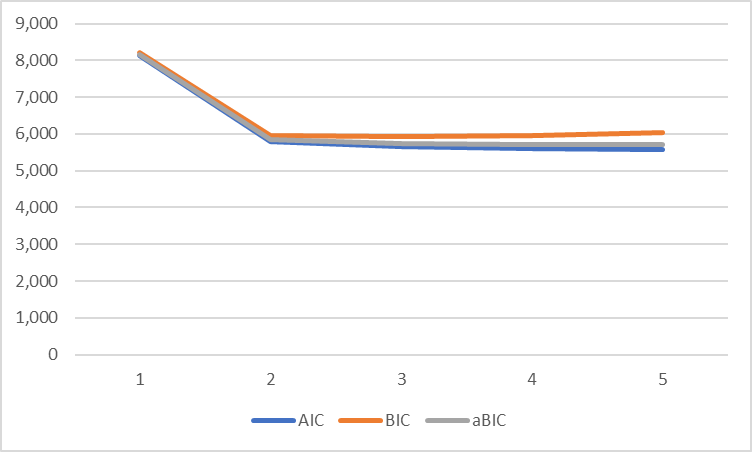 |
| No. parameters | 23 | 38 | 59 | 80 | 101 |  |
| Log likelihood | -4,037 | -2,862 | -2,773 | -2,722 | -2,688 |  |
| AIC | 8,120 | 5,799 | 5,664 | 5,604 | 5,579 |  |
| BIC | 8,223 | 5,970 | 5,928 | 5,962 | 6,031 |  |
| aBIC | 8,150 | 5,849 | 5,741 | 5,708 | 5,711 |  |
| Smallest class % | n/a | 32.46 | 11.45 | 6.65 | 3.75 |  |
| Entropy | n/a | 0.82 | 0.89 | 0.85 | 0.86 |  |
| COVID-19 > 12 weeks ago (n = 946) | | | | | | 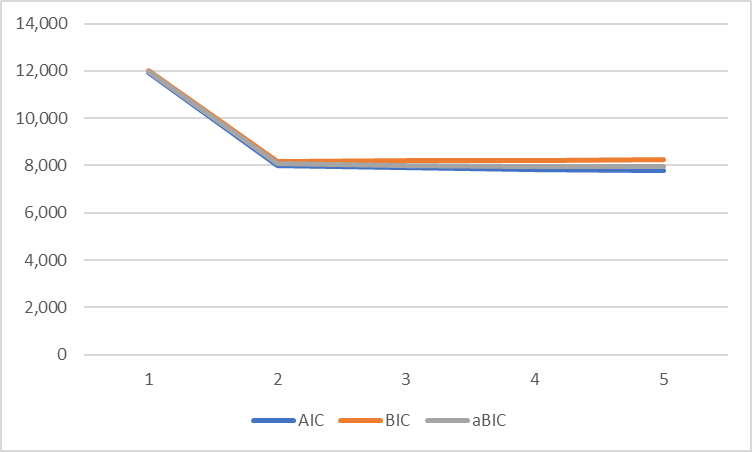 |
| No. parameters | 23 | 38 | 59 | 80 | 101 |  |
| Log likelihood | -5,939 | -3,963 | -3,894 | -3,834 | -3,790 |  |
| AIC | 11,924 | 8,001 | 7,907 | 7,828 | 7,781 |  |
| BIC | 12,035 | 8,185 | 8,193 | 8,217 | 8,271 |  |
| aBIC | 11,962 | 8,065 | 8,005 | 7,963 | 7,951 |  |
| Smallest class % | n/a | 31.71 | 3.97 | 3.43 | 3.15 |  |
| Entropy | n/a | 0.80 | 0.87 | 0.84 | 0.82 |  |

AIC: Akaike information criterion; BIC: Bayesian information criterion; aBIC: Sample size-adjusted Bayesian information criterion; n/a: not applicable.

#### **Table S6G**. Born in Bradford.

|  | Classes | | | | |  |
| --- | --- | --- | --- | --- | --- | --- |
|  | 1 | 2 | 3 | 4 | 5 | IC plot |
| No COVID-19 (n=561) | | | | | | 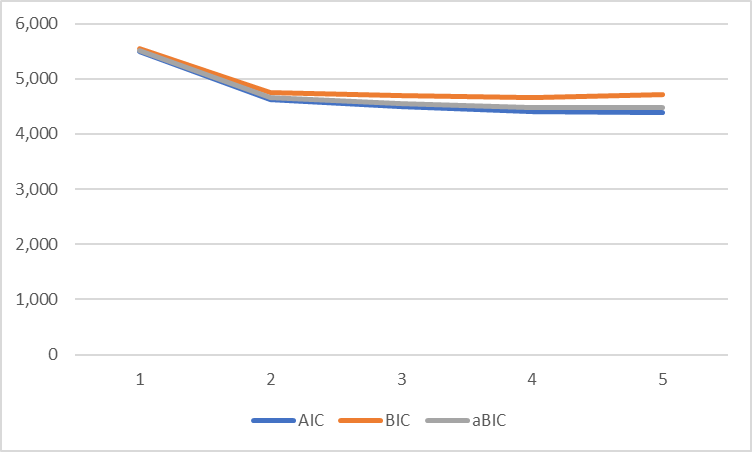 |
| No. parameters | 14 | 29 | 44 | 59 | 74 |  |
| Log likelihood | -2,731 | -2,281 | -2,206 | -2,143 | -2,121 |  |
| AIC | 5,490 | 4,619 | 4,501 | 4,403 | 4,391 |  |
| BIC | 5,550 | 4,745 | 4,691 | 4,659 | 4,711 |  |
| aBIC | 5,506 | 4,653 | 4,552 | 4,472 | 4,476 |  |
| Smallest class % | n/a | 0.30 | 0.09 | 0.07 | 0.04 |  |
| Entropy | n/a | 0.87 | 0.88 | 0.86 | 0.84 |  |

AIC: Akaike information criterion; BIC: Bayesian information criterion; aBIC: Sample size-adjusted Bayesian information criterion; n/a: not applicable. Note: Analyses were not possible in either the COVID-19 in last 12 weeks or COVID-19 > 12 weeks ago groups due to insufficient sample size (n = 11 and 58 respectively).

#### **Table S6H**. Understanding Society.

|  | Classes | | | | |  |
| --- | --- | --- | --- | --- | --- | --- |
|  | 1 | 2 | 3 | 4 | 5 | IC plot |
| Symptoms < 12 Weeks (n = 2,815) | | | | | | 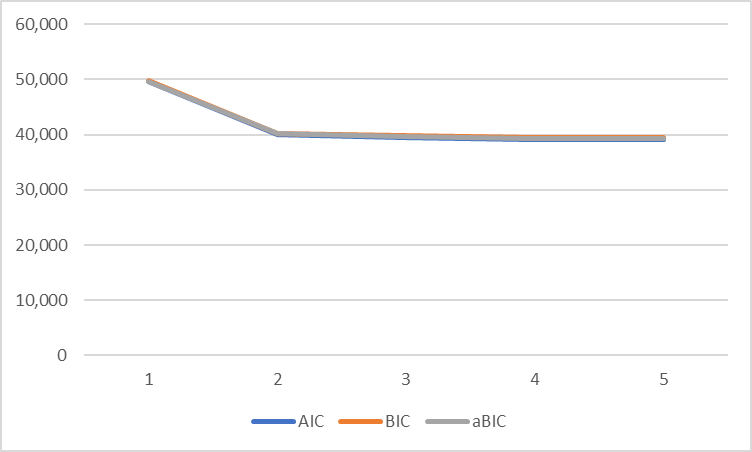 |
| No. parameters | 16 | 27 | 42 | 57 | 72 |  |
| Log likelihood | -24,800 | -20,032 | -19,727 | -19,548 | -19,453 |  |
| AIC | 49,632 | 40,117 | 39,538 | 39,210 | 39,051 |  |
| BIC | 49,727 | 40,278 | 39,787 | 39,549 | 39,479 |  |
| aBIC | 49,676 | 40,192 | 39,654 | 39,368 | 39,250 |  |
| Smallest class % | n/a | 43.1 | 25.9 | 19.3 | 10.5 |  |
| Entropy | n/a | 0.68 | 0.67 | 0.65 | 0.66 |  |
| Symptoms > 12 weeks (n = 410) | | | | | | 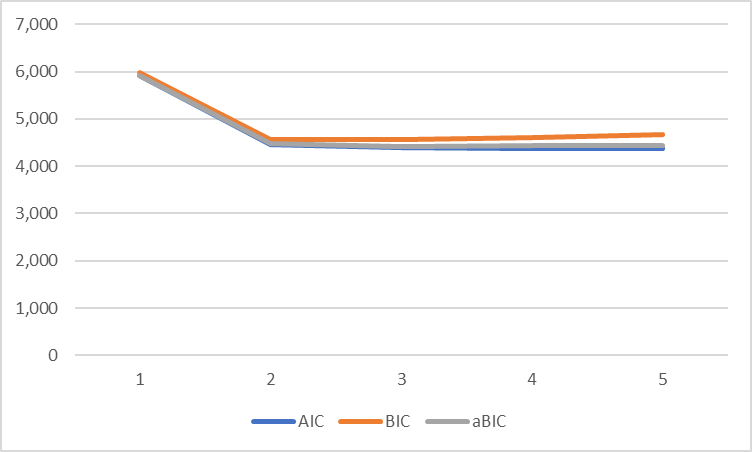 |
| No. parameters | 16 | 27 | 42 | 57 | 72 |  |
| Log likelihood | -2,938 | -2,204 | -2,153 | -2,132 | -2,115 |  |
| AIC | 5,909 | 4,462 | 4,389 | 4,379 | 4,374 |  |
| BIC | 5,973 | 4,571 | 4,558 | 4,608 | 4,663 |  |
| aBIC | 5,922 | 4,485 | 4,425 | 4,427 | 4,434 |  |
| Smallest class % | n/a | 38.4 | 28.8 | 13.3 | 2.2 |  |
| Entropy | n/a | 0.71 | 0.71 | 0.76 | 0.81 |  |

AIC: Akaike information criterion; BIC: Bayesian information criterion; aBIC: Sample size-adjusted Bayesian information criterion; n/a: not applicable.

#### **Table S6I**. Generation Scotland.

|  | Classes | | | | |  |
| --- | --- | --- | --- | --- | --- | --- |
|  | 1 | 2 | 3 | 4 | 5 | IC plot |
| Symptoms for less than 12 weeks (n = 960) | | | | | | 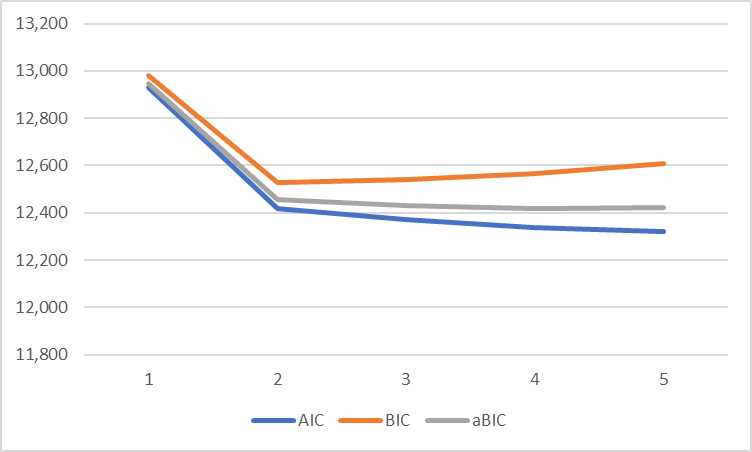 |
| No. parameters | 11 | 23 | 35 | 47 | 59 |  |
| Log likelihood | -6,453 | -6,185 | -6,150 | -6,122 | -6,101 |  |
| AIC | 12,929 | 12,416 | 12,370 | 12,337 | 12,321 |  |
| BIC | 12,982 | 12,528 | 12,540 | 12,566 | 12,608 |  |
| aBIC | 12,947 | 12,455 | 12,429 | 12,417 | 12,421 |  |
| Smallest class % | n/a | 48.6 | 17.1 | 14.6 | 14.6 |  |
| Entropy | n/a | 0.62 | 0.62 | 0.56 | 0.57 |  |
| Symptoms for over 12 weeks (n = 239) | | | | | | 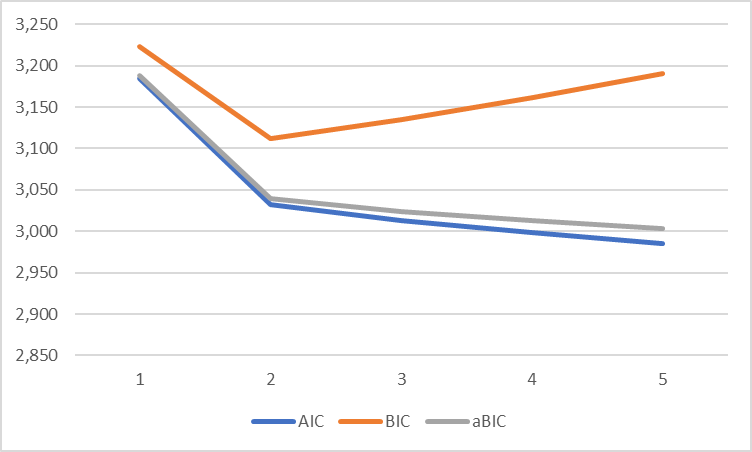 |
| No. parameters | 11 | 23 | 35 | 47 | 59 |  |
| Log likelihood | -1,581 | -1,493 | -1,471 | -1,452 | -1,433 |  |
| AIC | 3,185 | 3,032 | 3,013 | 2,999 | 2,985 |  |
| BIC | 3,223 | 3,112 | 3,135 | 3,162 | 3,190 |  |
| aBIC | 3,188 | 3,039 | 3,024 | 3,013 | 3,003 |  |
| Smallest class % | n/a | 41.8 | 19.4 | 6.2 | 4.4 |  |
| Entropy | n/a | 0.67 | 0.66 | - | - |  |

AIC: Akaike information criterion; BIC: Bayesian information criterion; aBIC: Sample size-adjusted Bayesian information criterion; n/a: not applicable.

### **Table S7**. Latent class analysis model fit statistics by COVID-19 status in each study (maximal symptom set).

#### **Table S7A**. Born in Bradford.

|  | Classes | | | | |  |
| --- | --- | --- | --- | --- | --- | --- |
|  | 1 | 2 | 3 | 4 | 5 | IC plot |
| No COVID-19 (n = 561) | | | | | | 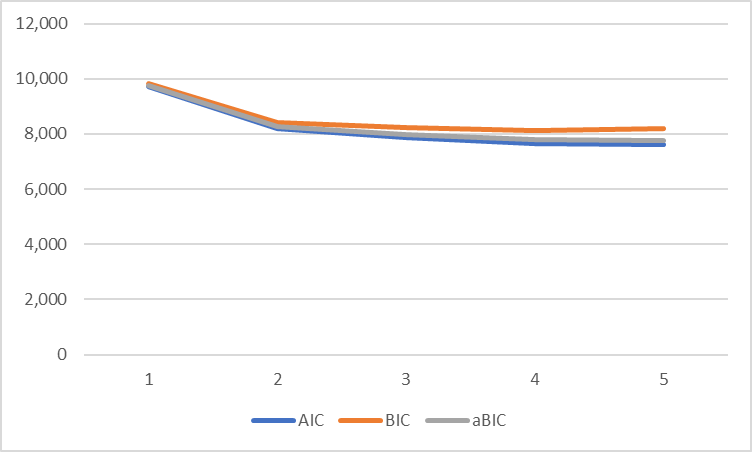 |
| No. parameters | 26 | 53 | 80 | 107 | 134 |  |
| Log likelihood | -4,838 | -4,044 | -3,858 | -3,725 | -3,672 |  |
| AIC | 9,727 | 8,193 | 7,876 | 7,663 | 7,612 |  |
| BIC | 9,840 | 8,423 | 8,223 | 8,127 | 8,192 |  |
| aBIC | 9,758 | 8,254 | 7,969 | 7,787 | 7,767 |  |
| Smallest class % | n/a | 0.33 | 0.07 | 0.07 | 0.04 |  |
| Entropy | n/a | 0.90 | - | - | - |  |

AIC: Akaike information criterion; BIC: Bayesian information criterion; aBIC: Sample size-adjusted Bayesian information criterion; not applicable. Note: Analyses were not possible in either the COVID-19 in last 12 weeks or COVID-19 > 12 weeks ago groups due to insufficient sample size (n = 11 and 58 respectively).

#### **Table S7B**. Avon Longitudinal Study of Parents and Children.

|  | Classes | | | | |  |
| --- | --- | --- | --- | --- | --- | --- |
|  | 1 | 2 | 3 | 4 | 5 | IC plot |
| No COVID-19 (n = 4,877) | | | | | | 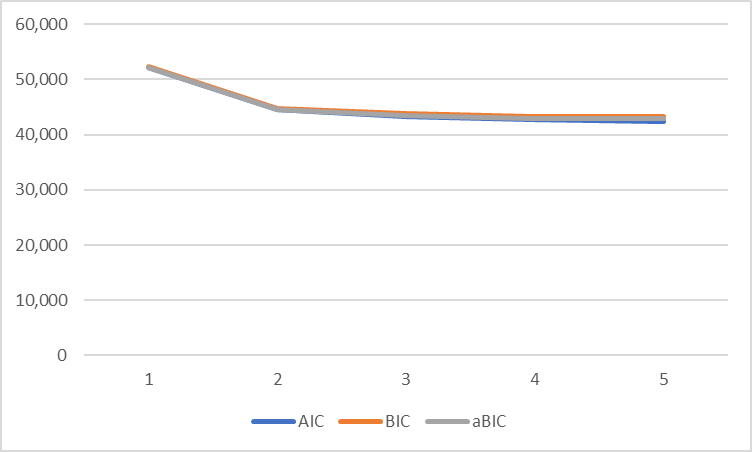 |
| No. parameters | 26 | 53 | 80 | 103 | 134 |  |
| Log likelihood | -26,052 | -22,182 | -21,544 | -21,222 | -21,088 |  |
| AIC | 52,155 | 44,469 | 43,247 | 42,658 | 42,444 |  |
| BIC | 52,324 | 44,813 | 43,767 | 43,353 | 43,314 |  |
| aBIC | 52,241 | 44,645 | 43,512 | 43,013 | 42,888 |  |
| Smallest class % | n/a | 23.1 | 5.6 | 5.0 | 4.2 |  |
| Entropy | n/a | 0.83 | 0.76 | 0.77 | 0.76 |  |
| COVID-19 in last 12 weeks (n = 181) | | | | | | 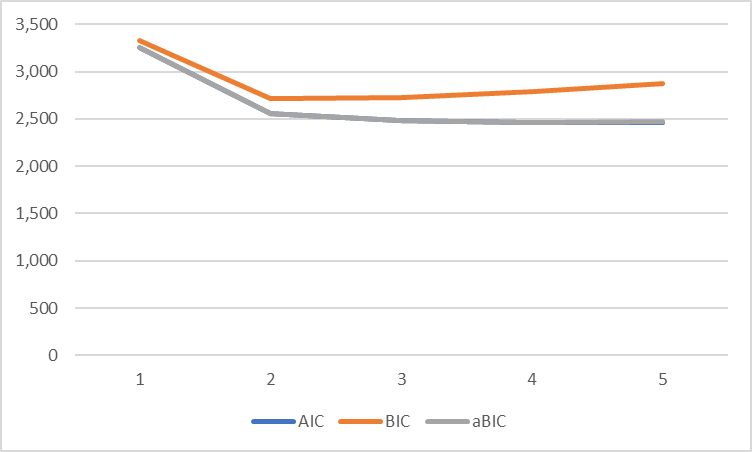 |
| No. parameters | 25 | 51 | 77 | 107 | 129 |  |
| Log likelihood | -1,601 | -1,226 | -1,164 | -1,127 | -1,104 |  |
| AIC | 3,253 | 2,554 | 2,482 | 2,459 | 2,466 |  |
| BIC | 3,333 | 2,717 | 2,728 | 2,789 | 2,879 |  |
| aBIC | 3,253 | 2,556 | 2,484 | 2,463 | 2,471 |  |
| Smallest class % | n/a | 0.23 | 12.2 | 8.8 | 6.1 |  |
| Entropy | n/a | 0.95 | 0.87 | 0.67 | 0.65 |  |
| COVID-19 > 12 weeks ago (n = 631) | | | | | | 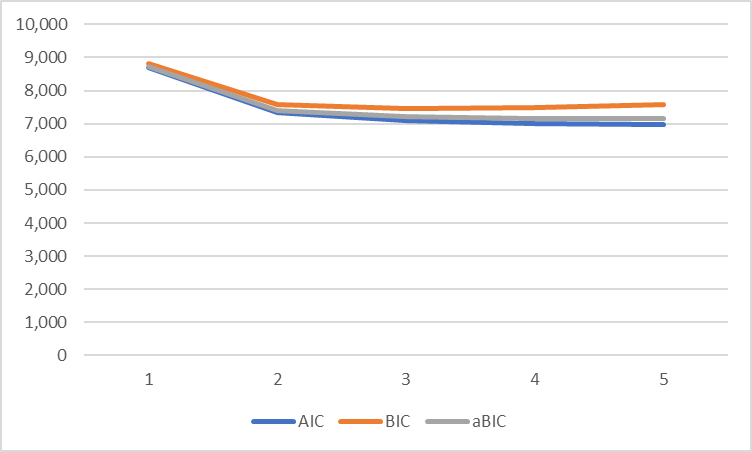 |
| No. parameters | 26 | 53 | 80 | 103 | 134 |  |
| Log likelihood | -4,320 | -3,611 | -3,472 | -3,395 | -3,353 |  |
| AIC | 8,692 | 7,328 | 7,103 | 7,004 | 6,975 |  |
| BIC | 8,807 | 7,564 | 7,459 | 7,480 | 7,571 |  |
| aBIC | 8,725 | 7,396 | 7,205 | 7,141 | 7,145 |  |
| Smallest class % | n/a | 25.4 | 5.4 | 5.4 | 3.0 |  |
| Entropy | n/a | 0.87 | 0.85 | 0.86 | 0.86 |  |

AIC: Akaike information criterion; BIC: Bayesian information criterion; aBIC: Sample size-adjusted Bayesian information criterion; n/a: not applicable.

#### **Table S7C**. TwinsUK

|  | Classes | | | | |  |
| --- | --- | --- | --- | --- | --- | --- |
|  | 1 | 2 | 3 | 4 | 5 | IC plot |
| No COVID-19 (n = 11383) | | | | | | 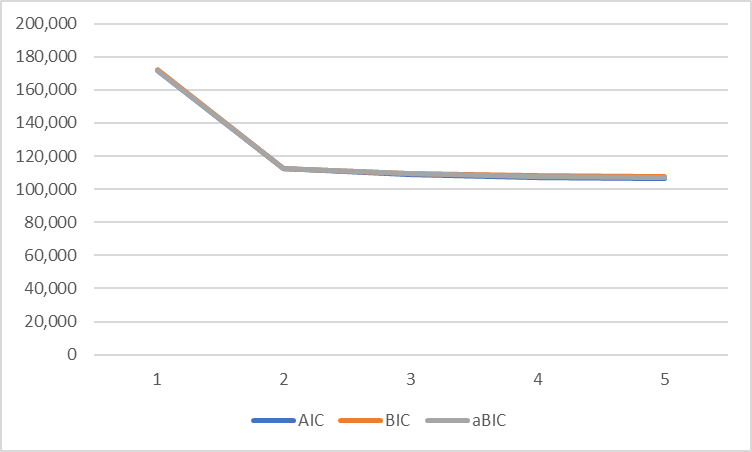 |
| No. parameters | 35 | 62 | 95 | 128 | 161 |  |
| Log likelihood | -85,848 | -56,013 | -54,293 | -53,385 | -52,944 |  |
| AIC | 171,767 | 112,150 | 108,775 | 107,025 | 106,211 |  |
| BIC | 172,024 | 112,605 | 109,472 | 107,965 | 107,392 |  |
| aBIC | 171,913 | 112,408 | 109,171 | 107,558 | 106,881 |  |
| Smallest class % | n/a | 19.37 | 2.44 | 0.38 | 0.38 |  |
| Entropy | n/a | 0.84 | 0.81 | 0.78 | 0.74 |  |
| COVID-19 in last 12 weeks (n = 653) | | | | | | 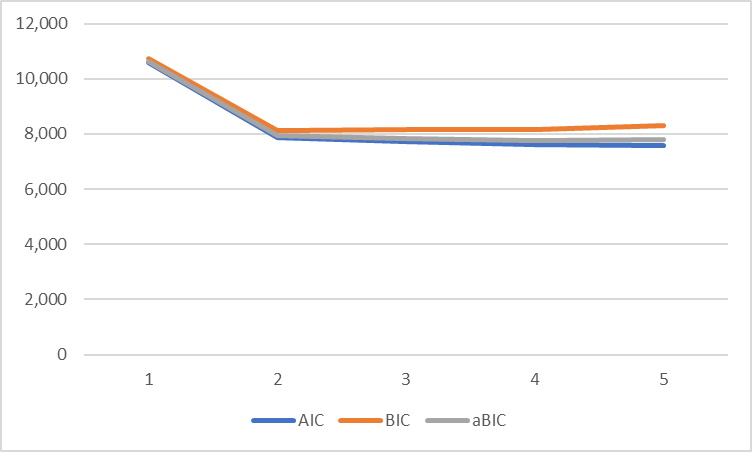 |
| No. parameters | 35 | 62 | 95 | 128 | 161 |  |
| Log likelihood | -5,256 | -3,870 | -3,765 | -3,672 | -3,630 |  |
| AIC | 10,582 | 7,864 | 7,721 | 7,601 | 7,581 |  |
| BIC | 10,739 | 8,141 | 8,147 | 8,174 | 8,302 |  |
| aBIC | 10,628 | 7,944 | 7,845 | 7,768 | 7,791 |  |
| Smallest class % | n/a | 31.79 | 7.18 | 6.77 | 4.63 |  |
| Entropy | n/a | 0.87 | 0.85 | 0.89 | 0.90 |  |
| COVID-19 > 12 weeks ago (n = 949) | | | | | | 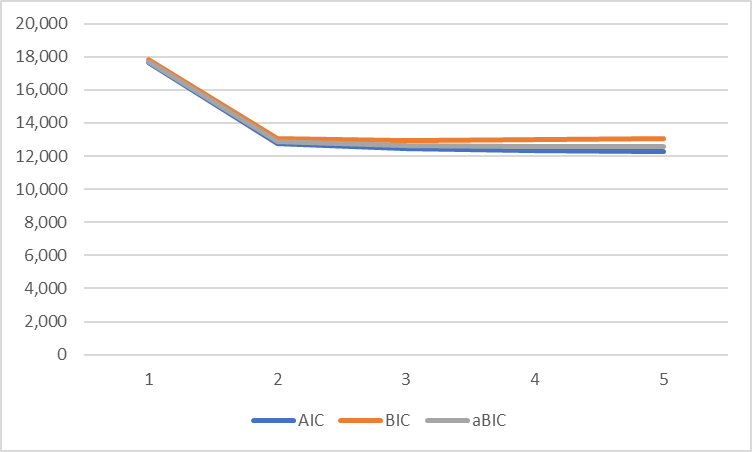 |
| No. parameters | 35 | 62 | 95 | 128 | 161 |  |
| Log likelihood | -8,793 | -6,314 | -6,131 | -6,050 | -5,987 |  |
| AIC | 17,657 | 12,752 | 12,453 | 12,357 | 12,297 |  |
| BIC | 17,827 | 13,053 | 12,914 | 12,978 | 13,078 |  |
| aBIC | 17,715 | 12,856 | 12,612 | 12,571 | 12,567 |  |
| Smallest class % | n/a | 31.59 | 6.46 | 3.78 | 2.99 |  |
| Entropy | n/a | 0.87 | 0.86 | 0.89 | 0.89 |  |

AIC: Akaike information criterion; BIC: Bayesian information criterion; aBIC: Sample size-adjusted Bayesian information criterion; n/a: not applicable.

#### **Table S7D**. Understanding Society.

|  | Classes | | | | |  |
| --- | --- | --- | --- | --- | --- | --- |
|  | 1 | 2 | 3 | 4 | 5 | IC plot |
| Symptoms < 12 Weeks (n = 2,815) | | | | | | 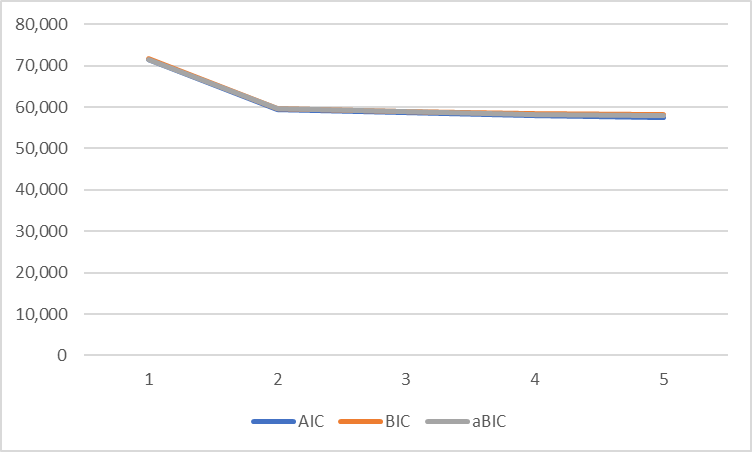 |
| No. parameters | 25 | 45 | 69 | 93 | 117 |  |
| Log likelihood | -35,743 | -29,666 | -29,239 | -28,853 | -28,682 |  |
| AIC | 71,537 | 59,421 | 58,617 | 57,892 | 57,598 |  |
| BIC | 71,685 | 59,688 | 59,027 | 58,444 | 58,294 |  |
| aBIC | 71,606 | 59,545 | 58,808 | 58,149 | 57,922 |  |
| Smallest class % | n/a | 35.5 | 16.7 | 17.9 | 11.1 |  |
| Entropy | n/a | 0.801 | 0.736 | 0.719 | 0.715 |  |
| Symptoms > 12 weeks (n = 410) | | | | | | 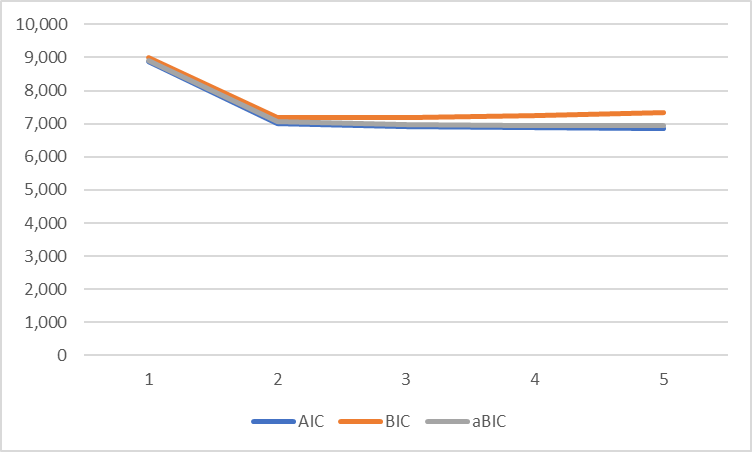 |
| No. parameters | 25 | 45 | 69 | 93 | 117 |  |
| Log likelihood | -4,417 | -3,464 | -3,385 | -3,346 | -3,309 |  |
| AIC | 8,884 | 7,018 | 6,909 | 6,879 | 6,852 |  |
| BIC | 8,985 | 7,198 | 7,186 | 7,252 | 7,322 |  |
| aBIC | 8,905 | 7,056 | 6,967 | 6,957 | 6,950 |  |
| Smallest class % | n/a | 36.7 | 27.4 | 12.9 | 10.2 |  |
| Entropy | n/a | 0.841 | 0.806 | 0.825 | 0.824 |  |

AIC: Akaike information criterion; BIC: Bayesian information criterion; aBIC: Sample size-adjusted Bayesian information criterion; aLRT: n/a: not applicable.

#### **Table S7E**. Generation Scotland.

|  | Classes | | | | |  |
| --- | --- | --- | --- | --- | --- | --- |
|  | 1 | 2 | 3 | 4 | 5 | IC plot |
| Symptoms for less than 12 weeks (n = 960) | | | | | | 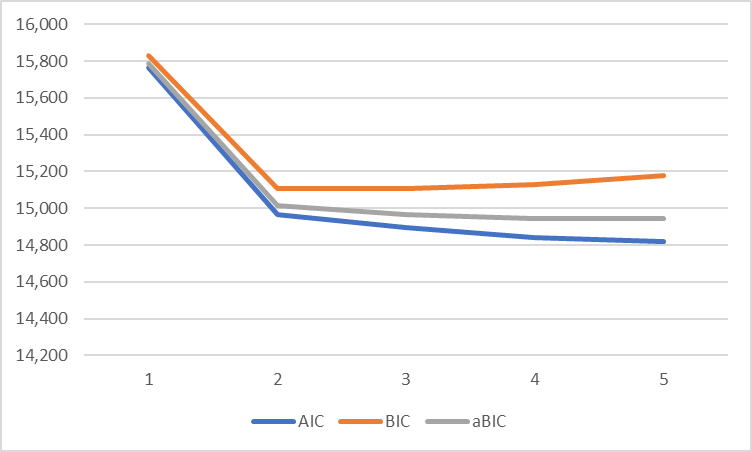 |
| No. parameters | 14 | 29 | 44 | 59 | 74 |  |
| Log likelihood | -7,867 | -7,455 | -7,402 | -7,362 | -7,335 |  |
| AIC | 15,763 | 14,968 | 14,893 | 14,843 | 14,817 |  |
| BIC | 15,831 | 15,109 | 15,107 | 15,130 | 15,177 |  |
| aBIC | 15,786 | 15,017 | 14,967 | 14,943 | 14,942 |  |
| Smallest class % | n/a | 49.4 | 10.1 | 9.5 | 3.5 |  |
| Entropy | n/a | 0.68 | 0.70 | - | - |  |
| Symptoms for over 12 weeks (n = 239) | | | | | | 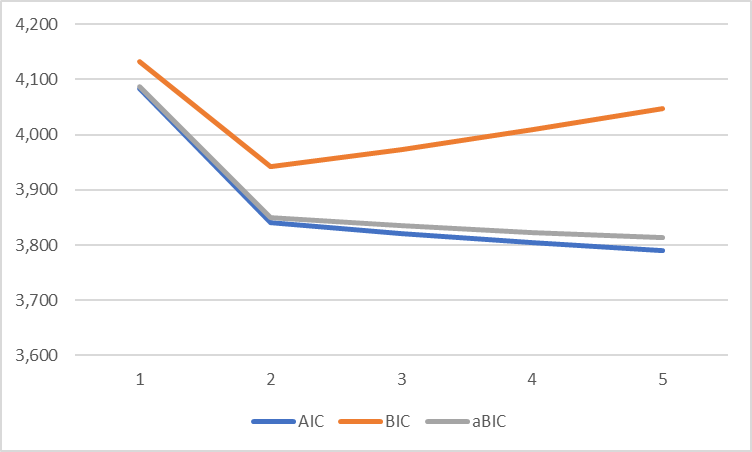 |
| No. parameters | 14 | 29 | 44 | 59 | 74 |  |
| Log likelihood | -2,028 | -1,892 | -1,867 | -1,844 | -1,821 |  |
| AIC | 4,084 | 3,841 | 3,821 | 3,805 | 3,790 |  |
| BIC | 4,132 | 3,942 | 3,974 | 4,010 | 4,048 |  |
| aBIC | 4,088 | 3,850 | 3,835 | 3,823 | 3,813 |  |
| Smallest class % | n/a | 47.6 | 15.2 | 4.4 | 4.5 |  |
| Entropy | n/a | 0.74 | 0.76 | - | - |  |

AIC: Akaike information criterion; BIC: Bayesian information criterion; aBIC: Sample size-adjusted Bayesian information criterion; n/a: not applicable.

### **Table S8**. Associations with symptom pattern 2 (vs. symptom pattern 1) in each study.

#### **Table S8A**. Associations with symptom patterns in the 1958 National Child Development Study.

| COVID-19 group | Sex | n | % of  total | n in symptom  pattern 2 | % in symptom  pattern 2 | OR | 95% CI  lower | 95% CI  upper |
| --- | --- | --- | --- | --- | --- | --- | --- | --- |
| No COVID-19 | Male | 2,797 | 46.6 | 602 | 21.5 | 1.00 | (ref) | (ref) |
| (n = 6,005) | Female | 3,208 | 53.4 | 889 | 27.2 | 1.40 | 1.24 | 1.57 |
| COVID-19 in last 12 weeks | Male | 89 | 43.8 | 26 | 29.2 | 1.00 | (ref) | (ref) |
| (n = 203) | Female | 114 | 56.2 | 52 | 45.6 | 2.03 | 1.13 | 3.66 |
| COVID-19 > 12 weeks ago | Male | 230 | 45.7 | 68 | 29.6 | 1.00 | (ref) | (ref) |
| (n = 503) | Female | 273 | 54.3 | 122 | 44.7 | 1.92 | 1.33 | 2.79 |
| COVID-19 group | Functional limitation following COVID-19 | n | % of  total | n in symptom  pattern 2 | % in symptom  pattern 2 | OR | 95% CI  lower | 95% CI  upper |
| COVID-19 > 12 weeks ago | Always able to function as normal | 98 | 19.5 | 24 | 24.5 | 1.00 | (ref) | (ref) |
| (n = 503) | 1 day or more, less than 2 weeks | 198 | 39.4 | 63 | 31.8 | 1.44 | 0.83 | 2.49 |
|  | 2 weeks or more, less than 4 weeks | 112 | 22.3 | 50 | 44.6 | 2.49 | 1.38 | 4.50 |
|  | 4 weeks or more, less than 12 weeks | 66 | 13.1 | 29 | 43.9 | 2.42 | 1.24 | 4.72 |
|  | 12 weeks or more | 29 | 5.8 | 24 | 82.8 | 14.80 | 5.09 | 43.06 |

#### **Table S6B**. 1970 British Cohort Study.

| COVID-19 group | Sex | n | % of  total | n in symptom  pattern 2 | % in symptom  pattern 2 | OR | 95% CI  lower | 95% CI  upper |
| --- | --- | --- | --- | --- | --- | --- | --- | --- |
| No COVID-19 | Male | 2,009 | 42.3 | 481 | 23.9 | 1.00 | (ref) | (ref) |
| (n = 4,746) | Female | 2,737 | 57.7 | 801 | 29.3 | 1.31 | 1.15 | 1.50 |
| COVID-19 in last 12 weeks | Male | 117 | 44.0 | 34 | 29.1 | 1.00 | (ref) | (ref) |
| (n = 266) | Female | 149 | 56.0 | 71 | 47.7 | 2.22 | 1.33 | 3.71 |
| COVID-19 > 12 weeks ago | Male | 262 | 42.4 | 87 | 33.2 | 1.00 | (ref) | (ref) |
| (n = 618) | Female | 356 | 57.6 | 163 | 45.8 | 1.70 | 1.22 | 2.37 |
| COVID-19 group | Functional limitation following COVID-19 | n | % of  total | n in symptom  pattern 2 | % in symptom  pattern 2 | OR | 95% CI  lower | 95% CI  upper |
| COVID-19 > 12 weeks ago | Always able to function as normal | 142 | 23.0 | 31 | 21.8 | 1.00 | (ref) | (ref) |
| (n = 618) | 1 day or more, less than 2 weeks | 275 | 44.5 | 97 | 35.3 | 1.95 | 1.22 | 3.12 |
|  | 2 weeks or more, less than 4 weeks | 111 | 18.0 | 55 | 49.6 | 3.52 | 2.04 | 6.06 |
|  | 4 weeks or more, less than 12 weeks | 44 | 7.1 | 33 | 75.0 | 10.74 | 4.87 | 23.67 |
|  | 12 weeks or more | 46 | 7.4 | 34 | 73.9 | 10.15 | 4.70 | 21.89 |

#### **Table S8C**. Next Steps.

| COVID-19 group | Sex | n | % of  total | n in symptom  pattern 2 | % in symptom  pattern 2 | OR | 95% CI  lower | 95% CI  upper |
| --- | --- | --- | --- | --- | --- | --- | --- | --- |
| No COVID-19 | Male | 1,234 | 37.8 | 277 | 22.5 | 1.00 | (ref) | (ref) |
| (n = 3,263) | Female | 2,029 | 62.2 | 643 | 31.7 | 1.60 | 1.36 | 1.89 |
| COVID-19 in last 12 weeks | Male | 78 | 30.0 | 24 | 30.8 | 1.00 | (ref) | (ref) |
| (n = 260) | Female | 182 | 70.0 | 65 | 35.7 | 1.25 | 0.71 | 2.21 |
| COVID-19 > 12 weeks ago | Male | 226 | 40.0 | 54 | 23.9 | 1.00 | (ref) | (ref) |
| (n = 565) | Female | 339 | 60.0 | 120 | 35.4 | 1.75 | 1.20 | 2.55 |
| COVID-19 group | Functional limitation following COVID-19 | n | % of  total | n in symptom  pattern 2 | % in symptom  pattern 2 | OR | 95% CI  lower | 95% CI  upper |
| COVID-19 > 12 weeks ago | Always able to function as normal | 135 | 23.9 | 26 | 19.3 | 1.00 | (ref) | (ref) |
| (n = 565) | 1 day or more, less than 2 weeks | 298 | 52.7 | 86 | 28.9 | 1.70 | 1.04 | 2.79 |
|  | 2 weeks or more, less than 4 weeks | 76 | 13.5 | 30 | 39.5 | 2.73 | 1.46 | 5.12 |
|  | 4 weeks or more, less than 12 weeks | 34 | 6.0 | 19 | 55.9 | 5.31 | 2.38 | 11.83 |
|  | 12 weeks or more | 22 | 3.9 | 13 | 59.1 | 6.06 | 2.34 | 15.68 |

#### **Table S8D**. Millennium Cohort Study.

| COVID-19 group | Sex | n | % of  total | n in symptom  pattern 2 | % in symptom  pattern 2 | OR | 95% CI  lower | 95% CI  upper |
| --- | --- | --- | --- | --- | --- | --- | --- | --- |
| No COVID-19 | Male | 1,304 | 39.7 | 319 | 24.5 | 1.00 | (ref) | (ref) |
| (n = 3,283) | Female | 1,979 | 60.3 | 816 | 41.2 | 2.17 | 1.86 | 2.53 |
| COVID-19 in last 12 weeks | Male | 89 | 35.0 | 18 | 20.2 | 1.00 | (ref) | (ref) |
| (n = 254) | Female | 165 | 65.0 | 55 | 33.3 | 1.97 | 1.07 | 3.63 |
| COVID-19 > 12 weeks ago | Male | 314 | 39.3 | 110 | 35.0 | 1.00 | (ref) | (ref) |
| (n = 800) | Female | 486 | 60.7 | 211 | 43.4 | 1.42 | 1.06 | 1.91 |
| COVID-19 group | Functional limitation following COVID-19 | n | % of  total | n in symptom  pattern 2 | % in symptom  pattern 2 | OR | 95% CI  lower | 95% CI  upper |
| COVID-19 > 12 weeks ago | Always able to function as normal | 288 | 36.0 | 85 | 29.5 | 1.00 | (ref) | (ref) |
| (n = 800) | 1 day or more, less than 2 weeks | 416 | 52.0 | 183 | 44.0 | 1.88 | 1.36 | 2.58 |
|  | 2 weeks or more, less than 4 weeks | 60 | 7.5 | 35 | 58.3 | 3.34 | 1.89 | 5.93 |
|  | 4 weeks or more, less than 12 weeks | 24 | 3.0 | 12 | 50.0 | 2.39 | 1.03 | 5.52 |
|  | 12 weeks or more | 12 | 1.5 | 6 | 50.0 | 2.39 | 0.75 | 7.62 |

#### **Table S8E**. Avon Longitudinal Study of Parents and Children.

| COVID-19 group | Sex | n | % of  total | n in symptom  pattern 2 | % in symptom  pattern 2 | OR | 95% CI  lower | 95% CI  upper |
| --- | --- | --- | --- | --- | --- | --- | --- | --- |
| No COVID-19 | Male | 2233 | 45.8 | 403 | 18.0 | 1.00 | (ref) | (ref) |
| (n = 4,877) | Female | 2644 | 54.2 | 751 | 28.4 | 1.80 | 1.57 | 2.07 |
| COVID-19 in last 12 weeks | Male | 52 | 28.7 | 11 | 21.2 | 1.00 | (ref) | (ref) |
| (n = 181) | Female | 129 | 71.3 | 28 | 21.7 | 1.03 | 0.48 | 2.34 |
| COVID-19 > 12 weeks ago | Male | 232 | 36.8 | 48 | 20.7 | 1.00 | (ref) | (ref) |
| (n = 631) | Female | 399 | 63.2 | 84 | 21.1 | 1.02 | 0.69 | 1.53 |
| COVID-19 group | Age (years) | n | % of  total | n in symptom  pattern 2 | % in symptom  pattern 2 | OR | 95% CI  lower | 95% CI  upper |
| No COVID-19 | <40 | 3454 | 70.8 | 901 | 26.1 | 1.00 | (ref) | (ref) |
| (n = 4,877) | 40-59 | 595 | 12.2 | 112 | 18.8 | 0.66 | 0.53 | 0.81 |
|  | 60+ | 828 | 17.0 | 141 | 17.0 | 0.58 | 0.48 | 0.71 |
| COVID-19 in last 12 weeks | <40 | * | * | * | 21.4 | 1.00 | (ref) | (ref) |
| (n = 181) | 40-59 | <5 | * | * | 20.0 | 0.92 | 0.05 | 6.45 |
|  | 60+ | <5 | * | * | 33.3 | 1.84 | 0.08 | 19.61 |
| COVID-19 > 12 weeks ago | <40 | 507 | 80.3 | 109 | 21.5 | 1.00 | (ref) | (ref) |
| (n = 631) | 40-59 | 71 | 11.3 | 17 | 23.9 | 1.15 | 0.62 | 2.03 |
|  | 60+ | 53 | 8.4 | 6 | 11.3 | 0.47 | 0.18 | 1.04 |

#### **Table S8F**. TwinsUK

| COVID-19 group | Sex | n | % of  total | n in symptom  pattern 2 | % in symptom  pattern 2 | OR | 95% CI  lower | 95% CI  upper |
| --- | --- | --- | --- | --- | --- | --- | --- | --- |
| No COVID-19 | Male | 1,281 | 11.3 | 146 | 11.4 | 1.00 | (ref) | (ref) |
| (n = 11,289) | Female | 10,008 | 87.9 | 1773 | 17.7 | 1.67 | 1.40 | 2.01 |
| COVID-19 in last 12 weeks | Male | 74 | 11.3 | 61 | 27.0 | 1.00 | (ref) | (ref) |
| (n = 631) | Female | 557 | 85.4 | 437 | 33.6 | 1.36 | 0.81 | 2.40 |
| COVID-19 > 12 weeks ago | Male | 108 | 11.4 | 22 | 20.4 | 1.00 | (ref) | (ref) |
| (n = 943) | Female | 835 | 88.3 | 264 | 31.6 | 1.81 | 1.13 | 3.02 |
| COVID-19 group | Age (years) | n | % of  total | n in symptom  pattern 2 | % in symptom  pattern 2 | OR | 95% CI  lower | 95% CI  upper |
| No COVID-19 | <40 | 1247 | 11.0 | 325 | 26.1 | 1.00 | (ref) | (ref) |
| (n = 11,294) | 40-59 | 3122 | 27.4 | 599 | 19.2 | 0.67 | 0.58 | 0.79 |
|  | 60+ | 6925 | 60.9 | 995 | 14.4 | 0.48 | 0.41 | 0.55 |
| COVID-19 in last 12 weeks | <40 | 129 | 19.8 | 42 | 32.6 | 1.00 | (ref) | (ref) |
| (n = 631) | 40-59 | 259 | 39.7 | 88 | 34.0 | 1.07 | 0.68 | 1.68 |
|  | 60+ | 243 | 37.3 | 77 | 31.7 | 0.96 | 0.61 | 1.52 |
| COVID-19 > 12 weeks ago | <40 | 215 | 22.7 | 71 | 33.0 | 1.00 | (ref) | (ref) |
| (n = 944) | 40-59 | 388 | 41.0 | 118 | 30.4 | 0.89 | 0.62 | 1.27 |
|  | 60+ | 341 | 36.0 | 97 | 28.4 | 0.81 | 0.56 | 1.17 |
| COVID-19 group | Functional limitation following COVID-19 | n | % of  total | n in symptom  pattern 2 | % in symptom  pattern 2 | OR | 95% CI  lower | 95% CI  upper |
| COVID-19 > 12 weeks ago | Always able to function as normal | 218 | 23.0 | 53 | 24.3 | 1.00 | (ref) | (ref) |
| (n = 943) | 1 day or more, less than 2 weeks | 523 | 55.3 | 154 | 29.4 | 1.30 | 0.91 | 1.88 |
|  | 2 weeks or more, less than 4 weeks | 118 | 12.5 | 41 | 34.7 | 1.66 | 1.01 | 2.70 |
|  | 4 weeks or more, less than 12 weeks | 58 | 6.1 | 17 | 29.3 | 1.29 | 0.67 | 2.43 |
|  | 12 weeks or more | 26 | 2.7 | 21 | 80.8 | 13.08 | 5.05 | 40.71 |

#### **Table S8G**. Born in Bradford.

| COVID-19 group | Sex | n | % of  total | n in symptom  pattern 2 | % in symptom  pattern 2 | OR | 95% CI  lower | 95% CI  upper |
| --- | --- | --- | --- | --- | --- | --- | --- | --- |
| No COVID-19 | Male | 32 | 5.7 | 9 | 28.1 | 1.00 | (ref) | (ref) |
| (n = 561) | Female | 529 | 94.3 | 159 | 30.1 | 1.10 | 0.50 | 2.43 |
| COVID-19 group | Age (years) | n | % of  total | n in symptom  pattern 2 | % in symptom  pattern 2 | OR | 95% CI  lower | 95% CI  upper |
| No COVID-19 | <40 | 211 | 39.89 | 59 | 28.0 | 1.00 | (ref) | (ref) |
| (n = 561) | 40+ | 318 | 60.11 | 100 | 31.4 | 1.18 | 0.81 | 1.73 |

Notes: Age information on male partners is missing, only females included in the age adjusted models. Analyses were not possible in either the COVID-19 in last 12 weeks or COVID-19 > 12 weeks ago groups due to insufficient sample size (n = 11 and 58 respectively).

#### **Table S8H**. Understanding Society.

| COVID-19 group | Sex | n | % of  total | n in symptom  pattern 2 | % in symptom  pattern 2 | OR | 95% CI  lower | 95% CI  upper |
| --- | --- | --- | --- | --- | --- | --- | --- | --- |
| COVID-19 symptoms < 12 weeks | Male | 1,030 | 36.6 | 360 | 35.0 | 1.00 | (ref) | (ref) |
| (n = 2,815) | Female | 1,785 | 63.4 | 848 | 47.5 | 1.65 | 1.41 | 1.95 |
| COVID symptoms > 12 weeks | Male | 140 | 34.1 | 44 | 31.4 | 1.00 | (ref) | (ref) |
| (n = 410) | Female | 270 | 65.9 | 110 | 40.7 | 1.24 | 0.75 | 2.06 |
| COVID-19 group | Age (years) | n | % of  total | n in symptom  pattern 2 | % in symptom  pattern 2 | OR | 95% CI  lower | 95% CI  upper |
| COVID-19 symptoms < 12 weeks | <40 | 959 | 34.1 | 444 | 46.3 | 1.00 | (ref) | (ref) |
| (n = 2,815) | 40-59 | 1,282 | 45.5 | 573 | 44.7 | 0.98 | 0.82 | 1.16 |
|  | 60+ | 574 | 20.4 | 191 | 33.3 | 0.61 | 0.49 | 0.76 |
| COVID symptoms > 12 weeks | <40 | 74 | 18.0 | 26 | 35.1 | 1.00 | (ref) | (ref) |
| (n = 410) | 40-59 | 205 | 50.0 | 81 | 39.5 | 1.24 | 0.66 | 2.33 |
|  | 60+ | 131 | 32.0 | 47 | 35.9 | 0.98 | 0.49 | 1.96 |

#### **Table S8I**. Generation Scotland.

| COVID-19 group | Sex | n | % of  total | n in symptom  pattern 2 | % in symptom  pattern 2 | OR | 95% CI  lower | 95% CI  upper |
| --- | --- | --- | --- | --- | --- | --- | --- | --- |
| COVID-19 symptoms < 12 weeks | Male | 272 | 28.3 | 125 | 46.0 | 1.00 | (ref) | (ref) |
| (n = 960) | Female | 688 | 71.7 | 387 | 56.3 | 1.51 | 1.14 | 2.01 |
| COVID symptoms > 12 weeks | Male | 55 | 23.0 | 29 | 52.7 | 1.00 | (ref) | (ref) |
| (n = 239) | Female | 184 | 77.0 | 111 | 60.3 | 1.36 | 0.74 | 2.50 |
| COVID-19 group | Age (years) | n | % of  total | n in symptom  pattern 2 | % in symptom  pattern 2 | OR | 95% CI  lower | 95% CI  upper |
| COVID-19 symptoms < 12 weeks | <40 | 136 | 14.2 | 80 | 58.8 | 1.00 | (ref) | (ref) |
| (n = 954) | 40-59 | 450 | 46.9 | 260 | 57.8 | 0.96 | 0.65 | 1.41 |
|  | 60+ | 368 | 38.3 | 170 | 46.2 | 0.60 | 0.40 | 0.89 |
| COVID symptoms > 12 weeks | <40 | 38 | 15.9 | 25 | 65.8 | 1.00 | (ref) | (ref) |
| (n = 239) | 40-59 | 125 | 52.3 | 84 | 67.2 | 1.07 | 0.48 | 2.27 |
|  | 60+ | 76 | 31.8 | 31 | 40.8 | 0.36 | 0.16 | 0.80 |

### **Table S9**. Associations with symptom pattern 2 (vs. symptom pattern 1) meta-analysed across studies.

|  | No COVID-19 | | | | | |  | COVID-19 in last 12 weeks | | | | | |  | COVID-19 > 12 weeks ago | | | | | |
| --- | --- | --- | --- | --- | --- | --- | --- | --- | --- | --- | --- | --- | --- | --- | --- | --- | --- | --- | --- | --- |
|  | OR | OR 95%  CI lower | OR 95%  CI upper | I^2^ | Q | No.  studies |  | OR | OR 95%  CI lower | OR 95%  CI upper | I^2^ | Q | No.  studies |  | OR | OR 95%  CI lower | OR 95%  CI upper | I^2^ | Q | No.  studies |
| Sex |  |  |  |  |  |  |  |  |  |  |  |  |  |  |  |  |  |  |  |  |
| Male | 1.00 | (ref) | (ref) |  |  |  |  | 1.00 | (ref) | (ref) |  |  |  |  | 1.00 | (ref) | (ref) |  |  |  |
| Female | 1.32 | 0.92 | 1.89 | 97.1 | 206.9 | 7 |  | 1.63 | 1.28 | 2.09 | 4.4 | 5.2 | 6 |  | 1.56 | 1.31 | 1.86 | 27.4 | 6.9 | 6 |
| Age |  |  |  |  |  |  |  |  |  |  |  |  |  |  |  |  |  |  |  |  |
| <40 | 1.00 | (ref) | (ref) |  |  |  |  | 1.00 | (ref) | (ref) |  |  |  |  | 1.00 | (ref) | (ref) |  |  |  |
| 40-59 | 0.76 | 0.59 | 1.00 | 74.3 | 7.8 | 3 |  | 1.06 | 0.68 | 1.66 | 0.0 | 0.0 | 2 |  | 0.95 | 0.70 | 1.29 | 0.0 | 0.5 | 2 |
| 60+ | 0.52 | 0.43 | 0.63 | 60.1 | 2.5 | 2 |  | 0.98 | 0.62 | 1.53 | 0.0 | 0.2 | 2 |  | 0.72 | 0.46 | 1.11 | 19.0 | 1.2 | 2 |
| Functional limitation following COVID-19 |  |  |  |  |  |  |  |  |  |  |  |  |  |  |  |  |  |  |  |  |
| Always able to function as normal |  |  |  |  |  |  |  |  |  |  |  |  |  |  | 1.00 | (ref) | (ref) |  |  |  |
| 1 day or more, less than 2 weeks |  |  |  |  |  |  |  |  |  |  |  |  |  |  | 1.64 | 1.37 | 1.98 | 0.0 | 3.0 | 5 |
| 2 weeks or more, less than 4 weeks |  |  |  |  |  |  |  |  |  |  |  |  |  |  | 2.61 | 1.96 | 3.47 | 22.6 | 5.2 | 5 |
| 4 weeks or more, less than 12 weeks |  |  |  |  |  |  |  |  |  |  |  |  |  |  | 3.29 | 1.59 | 6.82 | 79.2 | 19.2 | 5 |
| 12 weeks or more |  |  |  |  |  |  |  |  |  |  |  |  |  |  | 8.08 | 4.52 | 14.44 | 42.6 | 7.0 | 5 |

### **Fig. S1**. Probability of each symptom in each symptom pattern by COVID-19 status in each study (core symptom set).

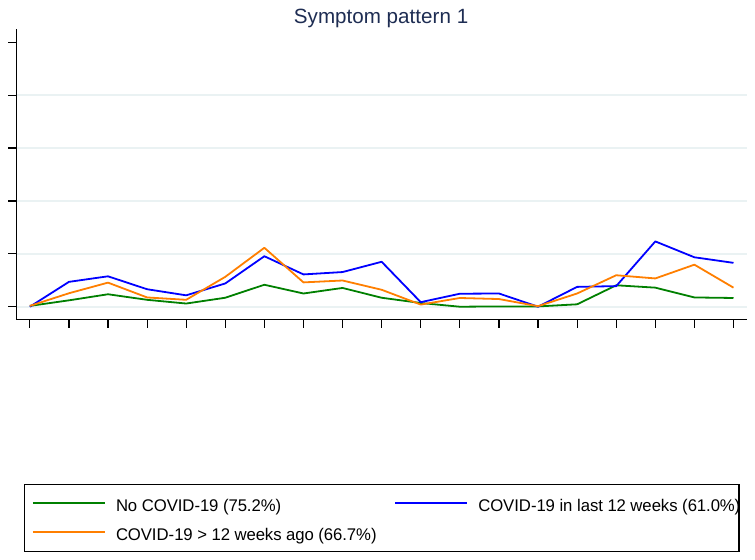

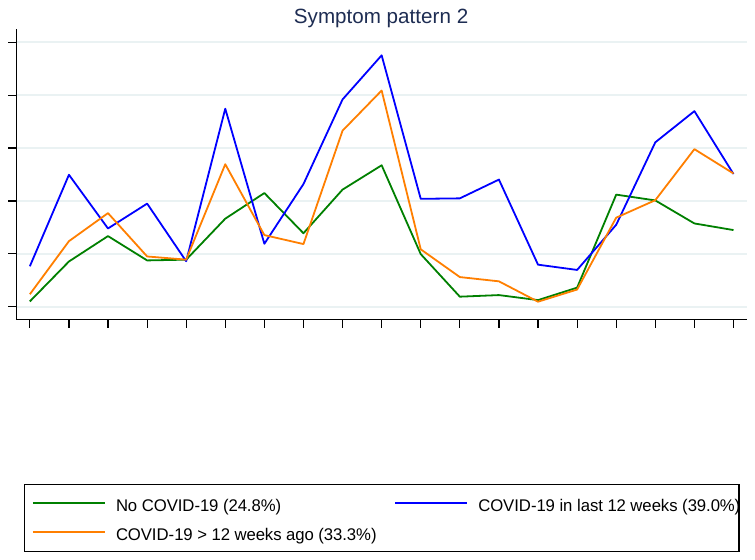

#### **Fig. S1A**. 1958 National Child Development Study.

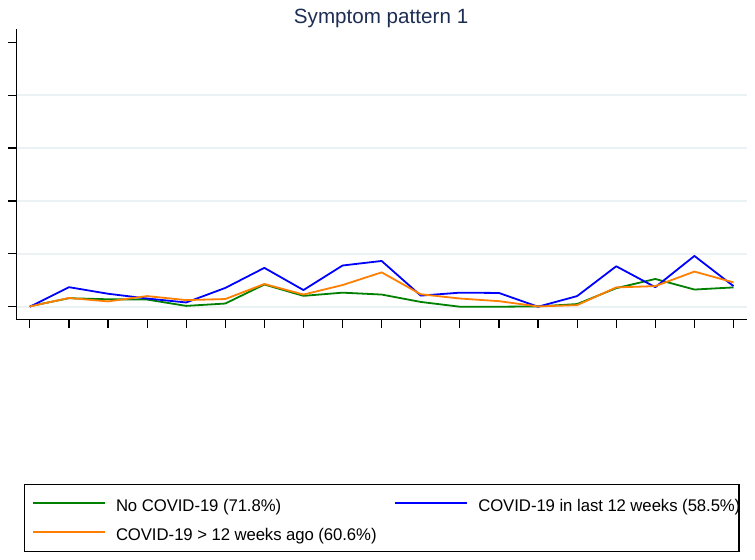

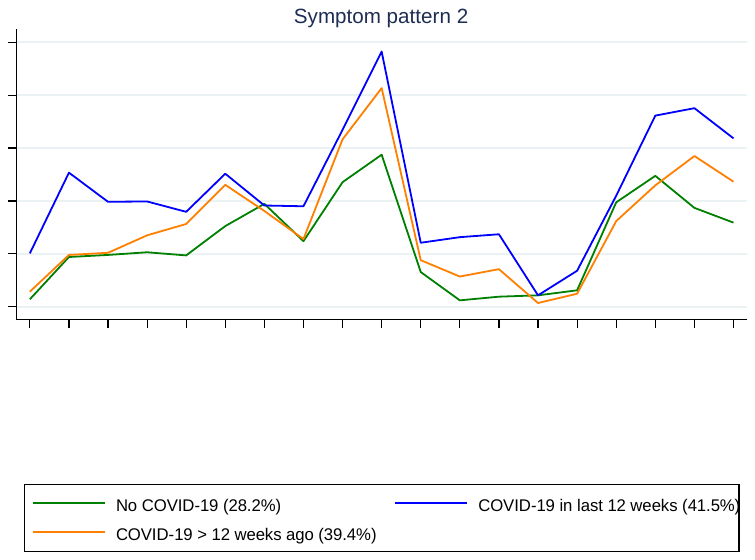

#### **Fig. S1B**. 1970 British Cohort Study.

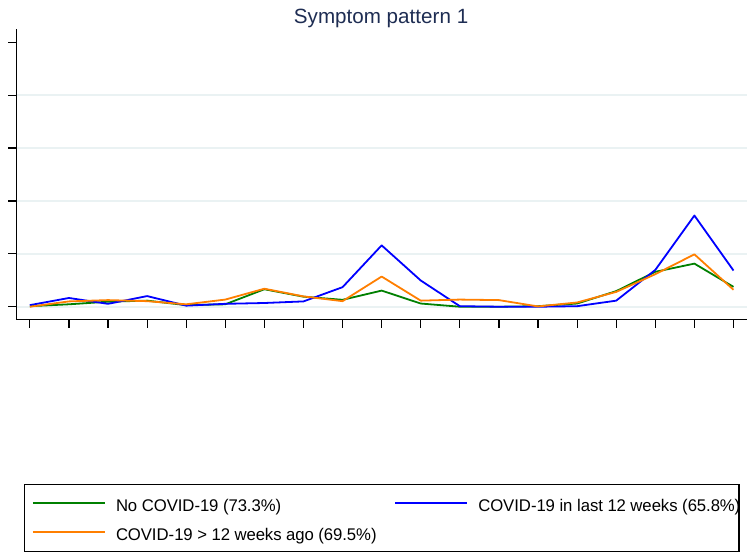

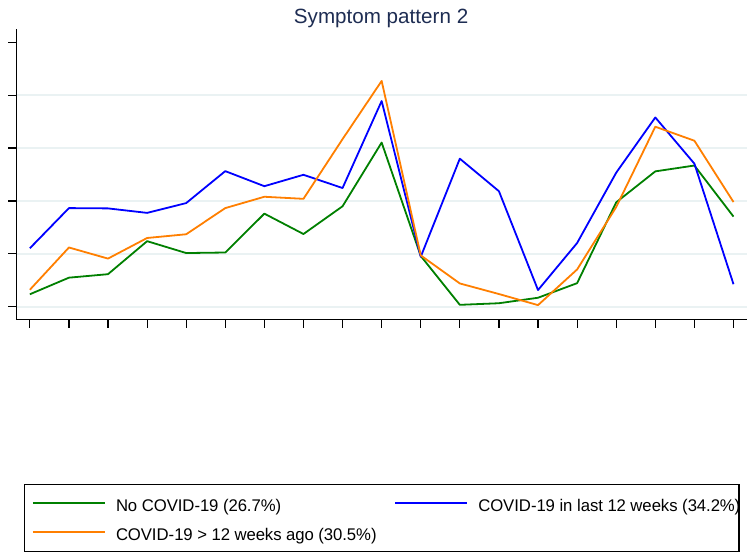

#### **Fig. S1C**. Next Steps.

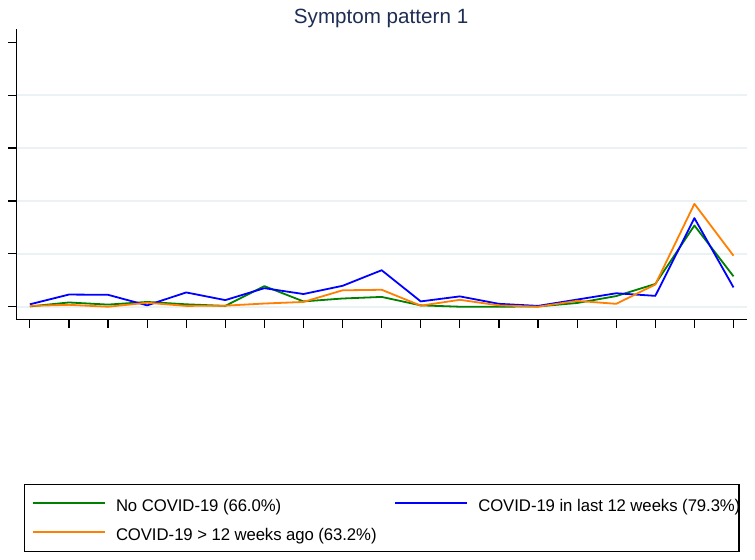

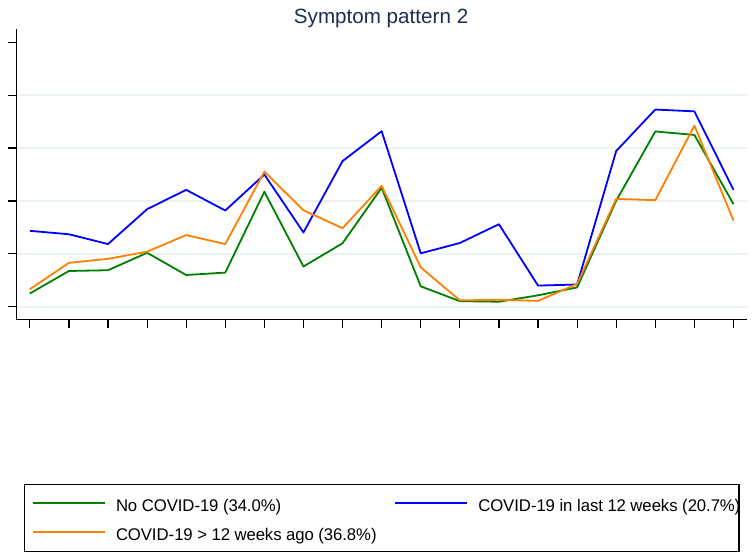

#### **Fig. S1D**. Millennium Cohort Study.

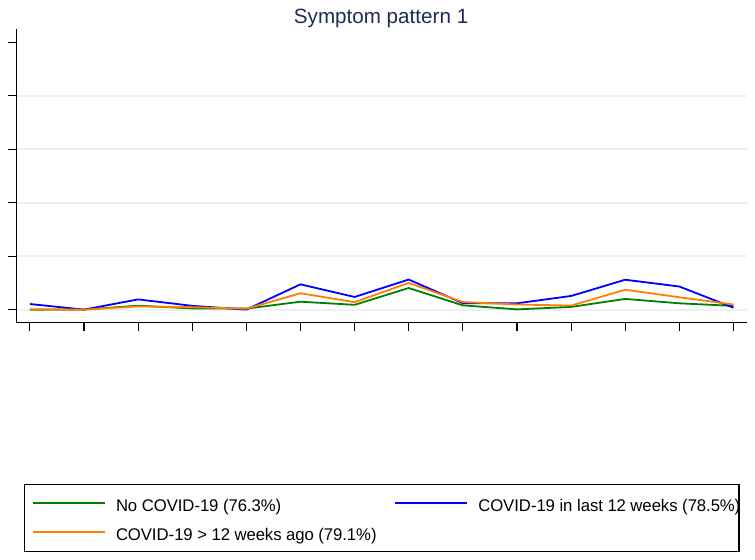

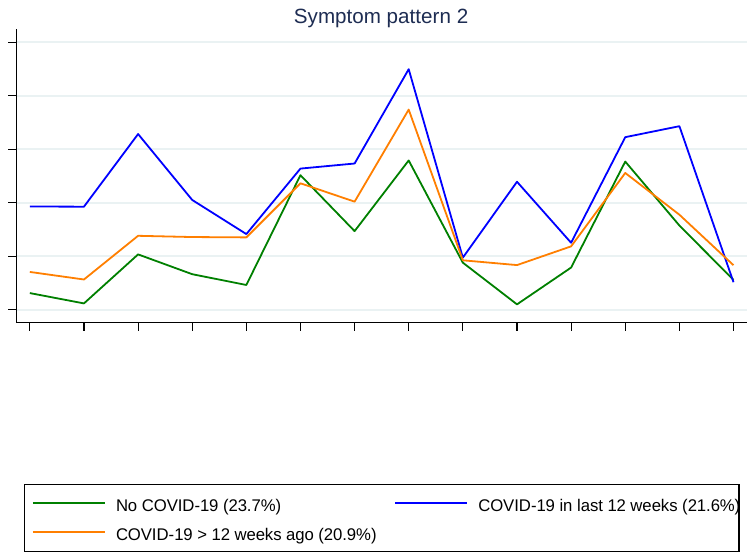

#### **Fig. S1E**. Avon Longitudinal Study of Parents and Children.

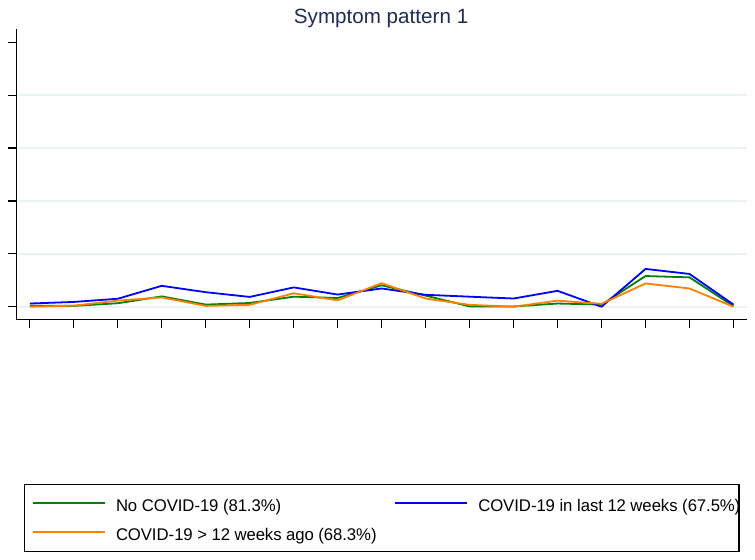

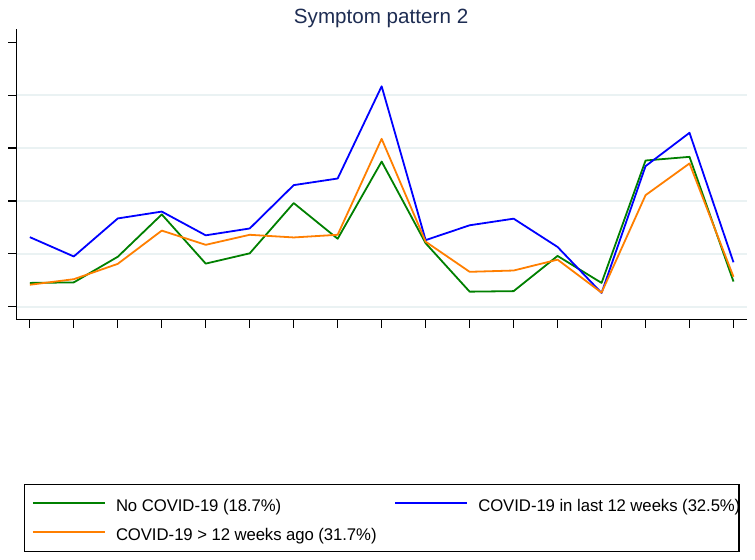

#### **Fig. S1F**. TwinsUK.

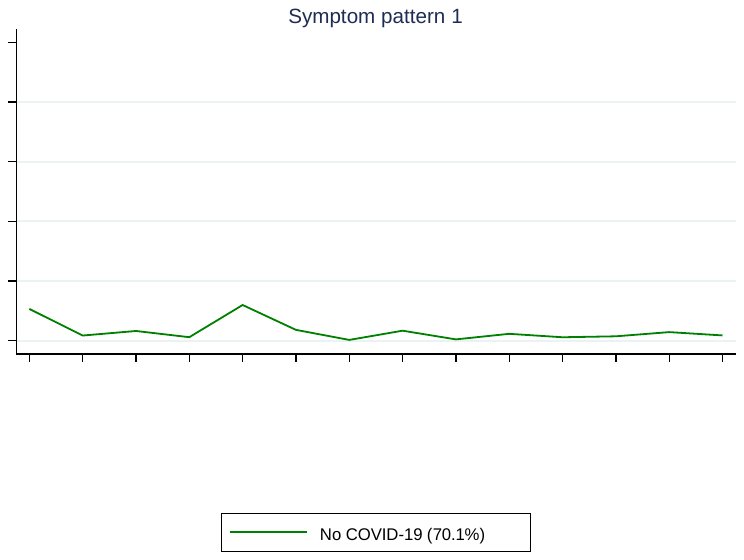

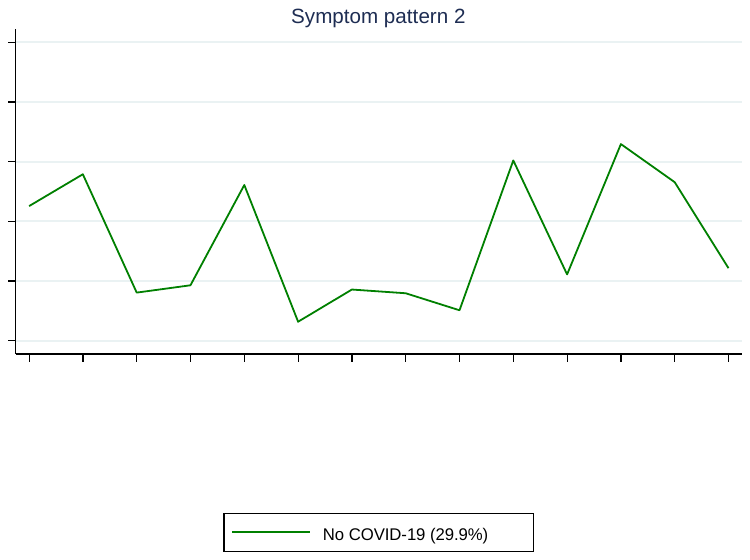

#### **Fig. S1G**. Born in Bradford.

Note: Analyses were not possible in either the COVID-19 in last 12 weeks or COVID-19 > 12 weeks ago groups due to insufficient sample size (n = 11 and 58 respectively).

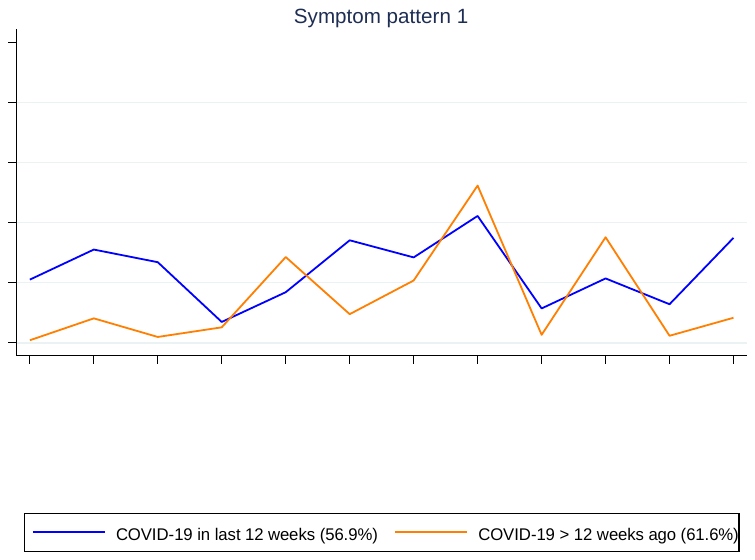

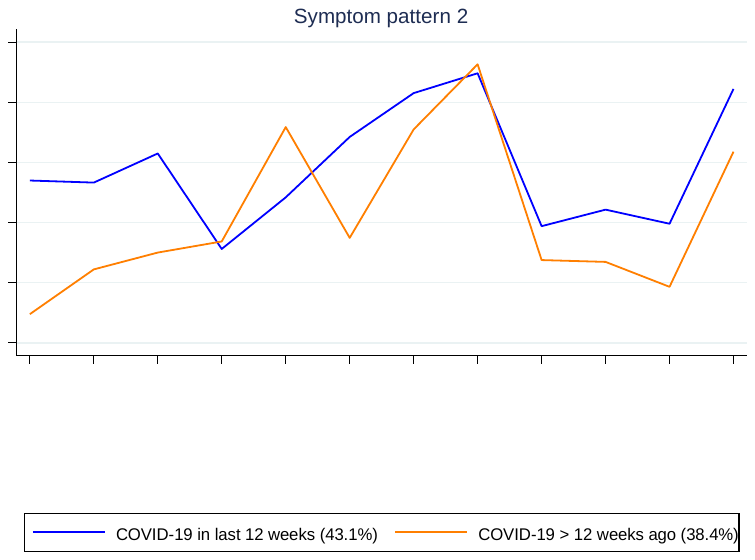

#### **Fig. S1H**. Understanding Society.

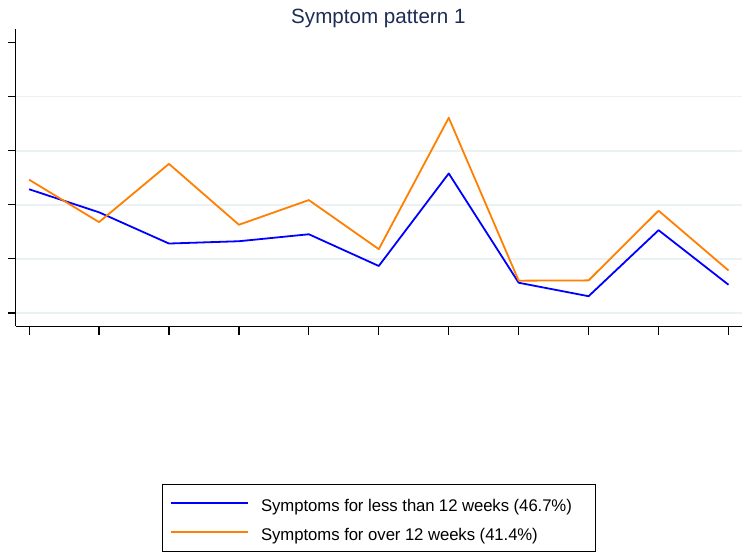

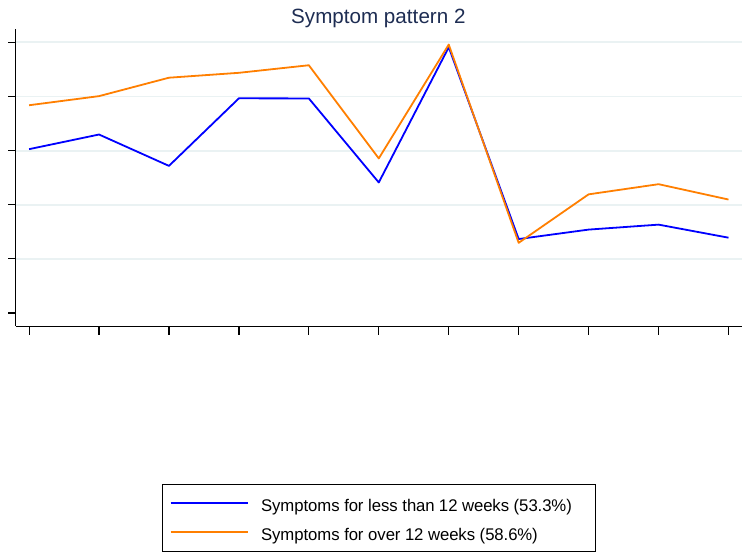

#### **Fig. S1I**. Generation Scotland.

### **Fig. S2**. Symptom probability differences comparing the two COVID-19 groups with the no COVID-19 group in each study (core symptom set). Bars represent 95% confidence intervals.

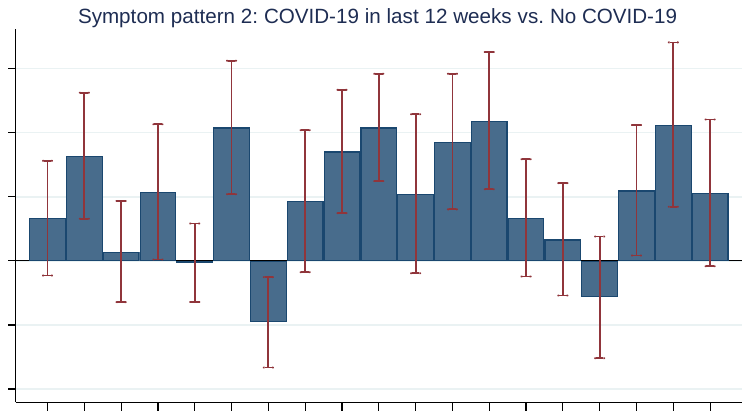

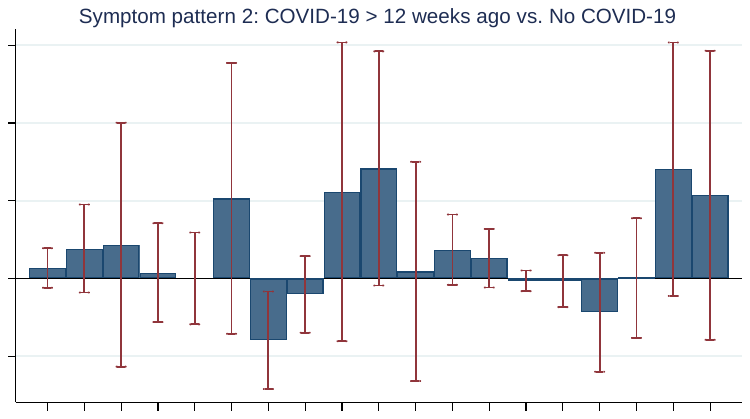

#### **Fig. S2A**. 1958 National Child Development Study.

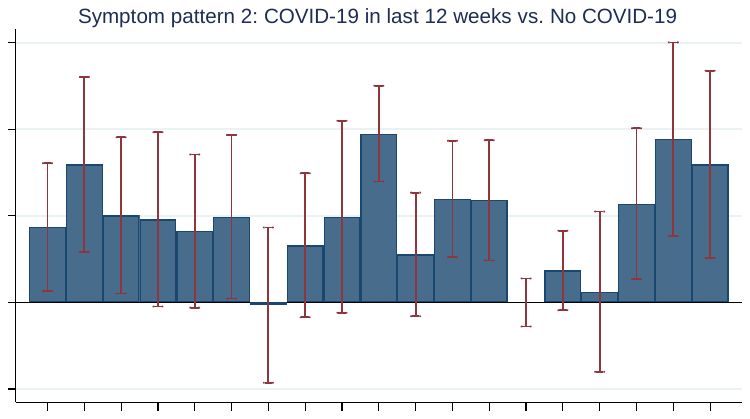

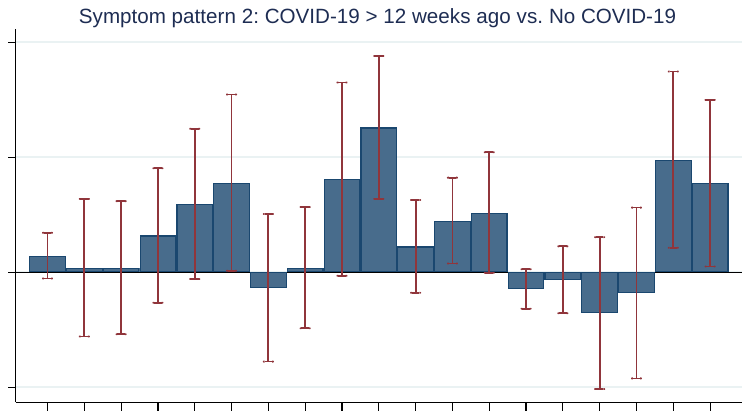

#### **Fig. S2B**. 1970 British Cohort Study.

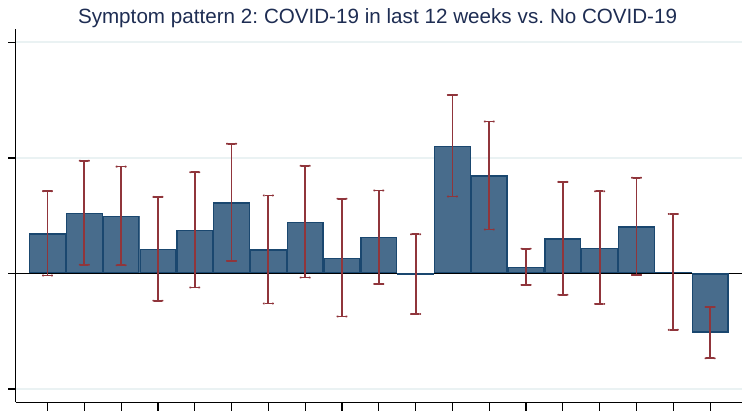

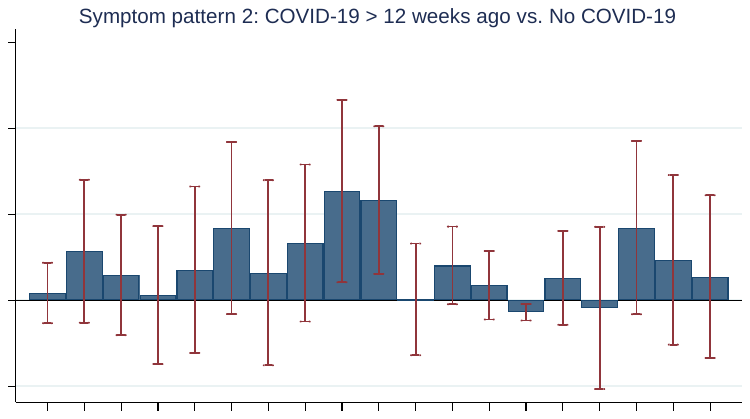

#### **Fig. S2C**. Next Steps.

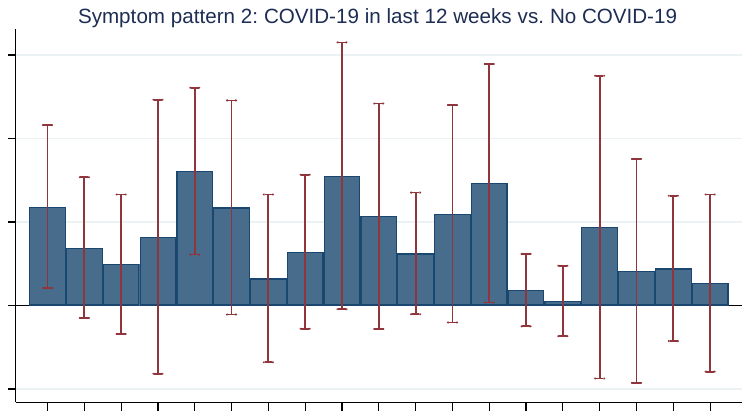

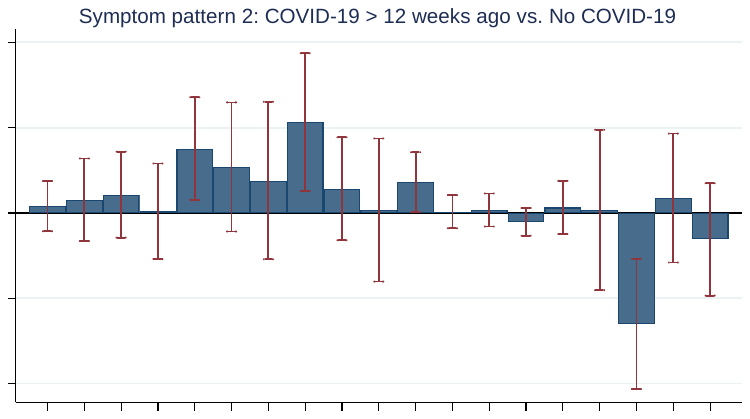

#### **Fig. S2D**. Millennium Cohort Study.

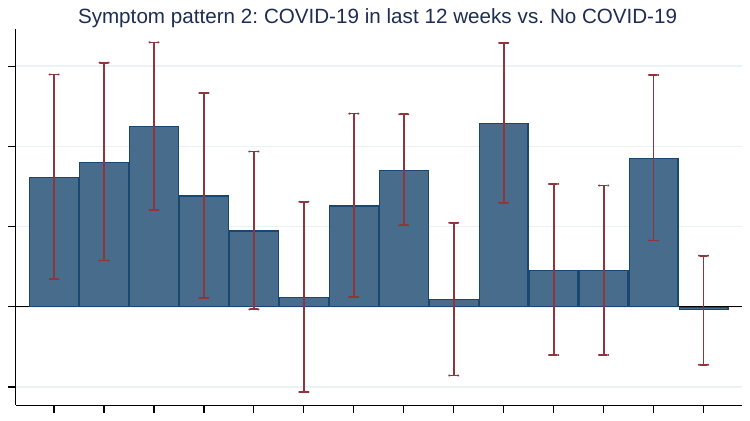

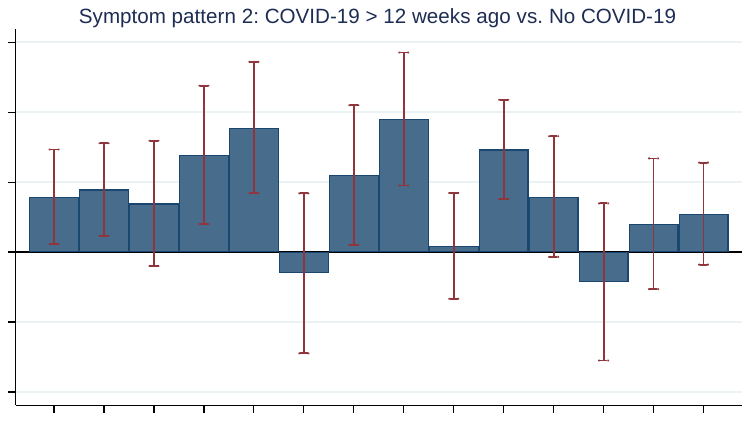

#### **Fig. S2E**. Avon Longitudinal Study of Parents and Children.

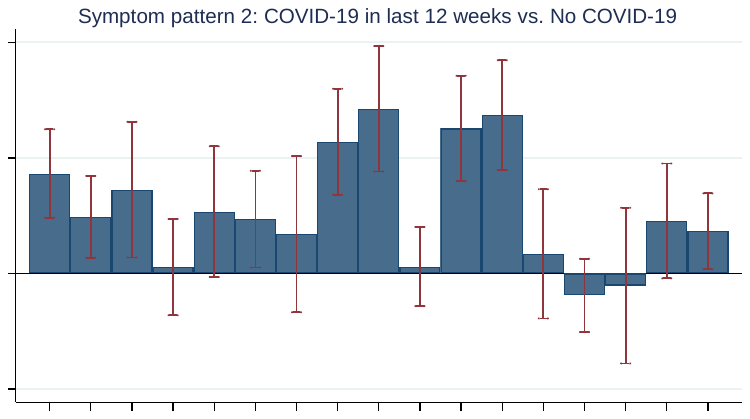

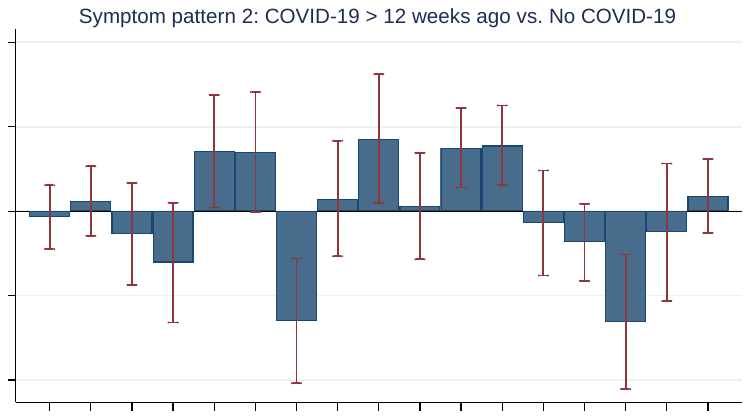

#### **Fig. S2F**. TwinsUK

### **Fig. S3**. Probability of each symptom in each symptom pattern by COVID-19 status in each study (maximal symptom set).

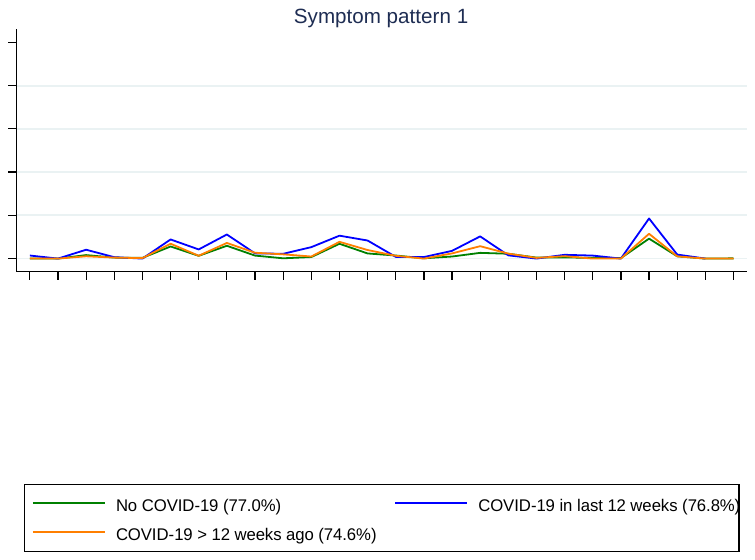

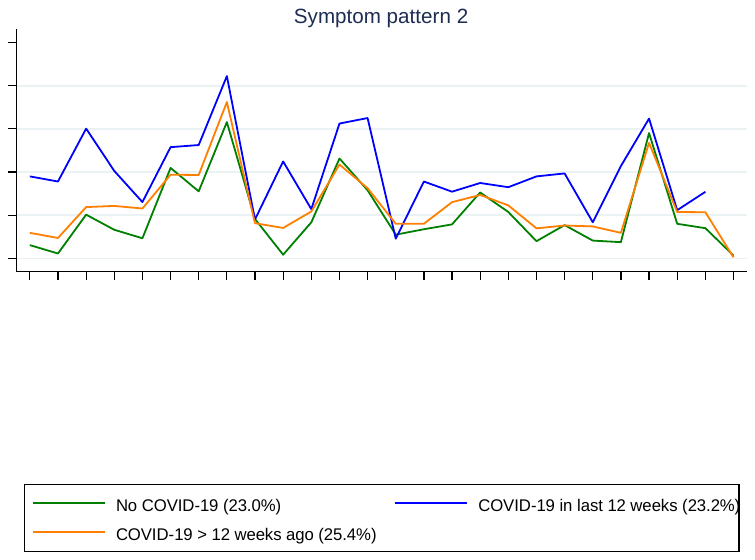

#### **Fig. S3A**. Avon Longitudinal Study of Parents and Children.

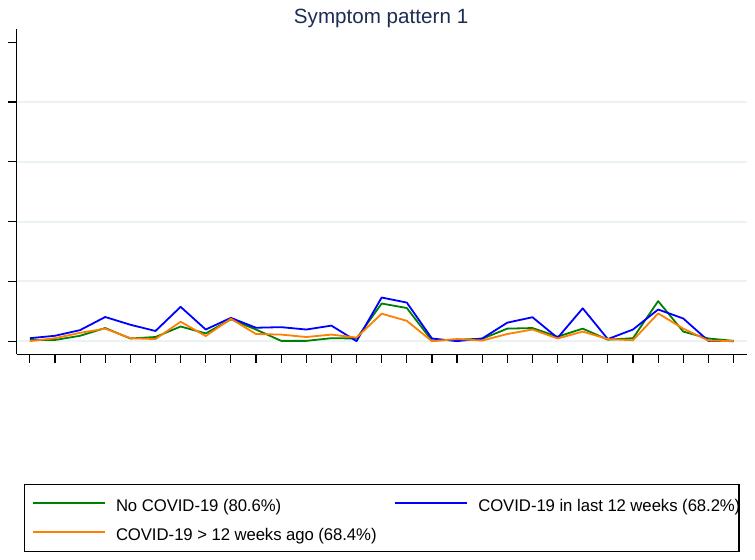

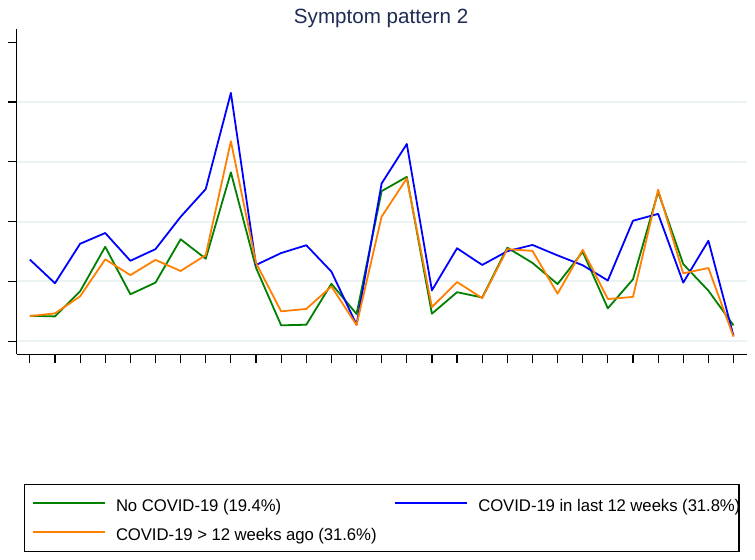

#### **Fig. S3B**. TwinsUK.

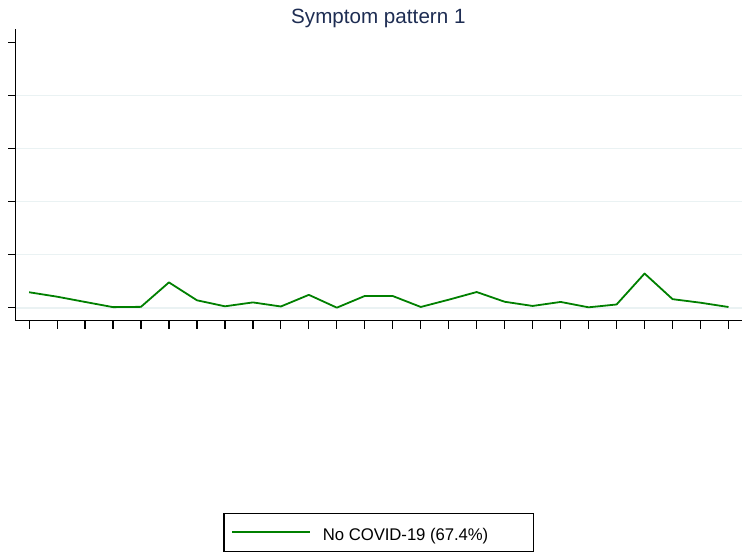

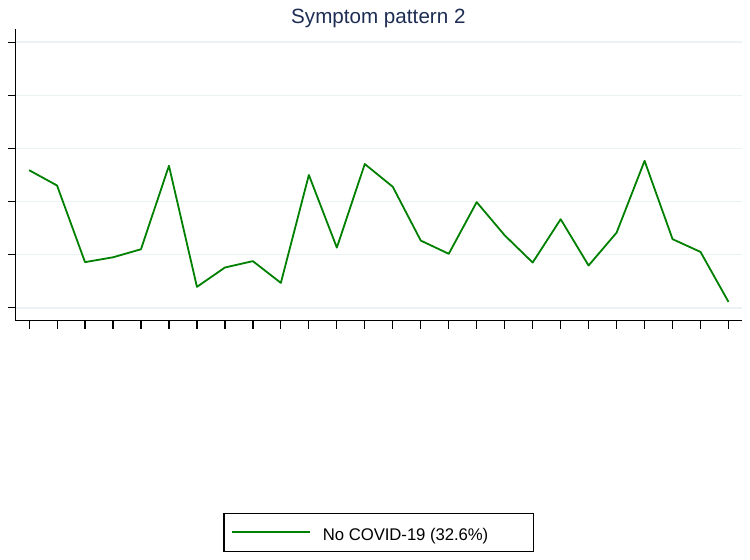

#### **Fig. S3C**. Born in Bradford.

Note: Analyses were not possible in either the COVID-19 in last 12 weeks or COVID-19 > 12 weeks ago groups due to insufficient sample size (n = 11 and 58 respectively).

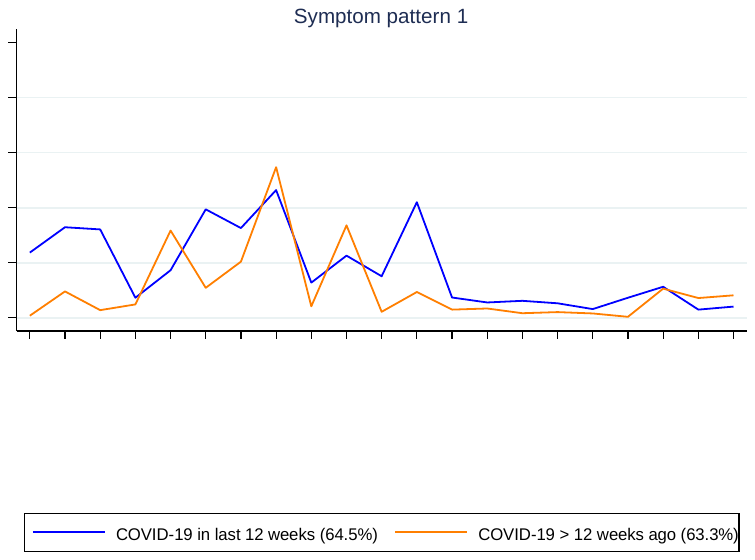

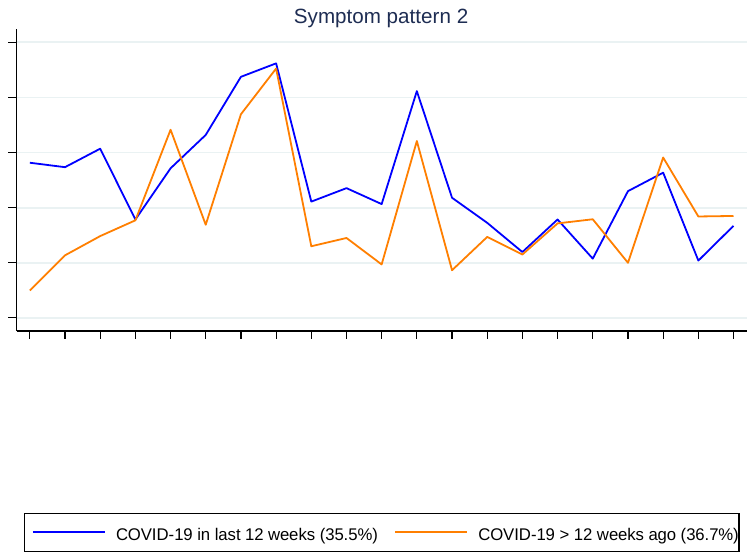

#### **Fig. S3D**. Understanding Society.

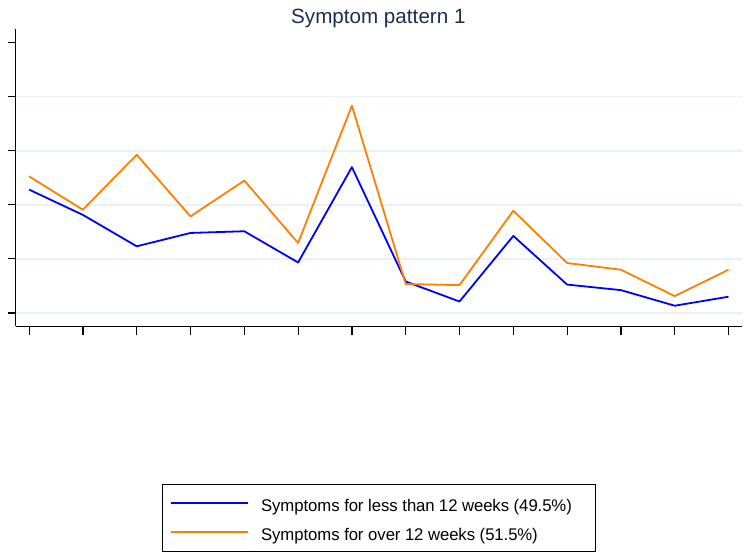

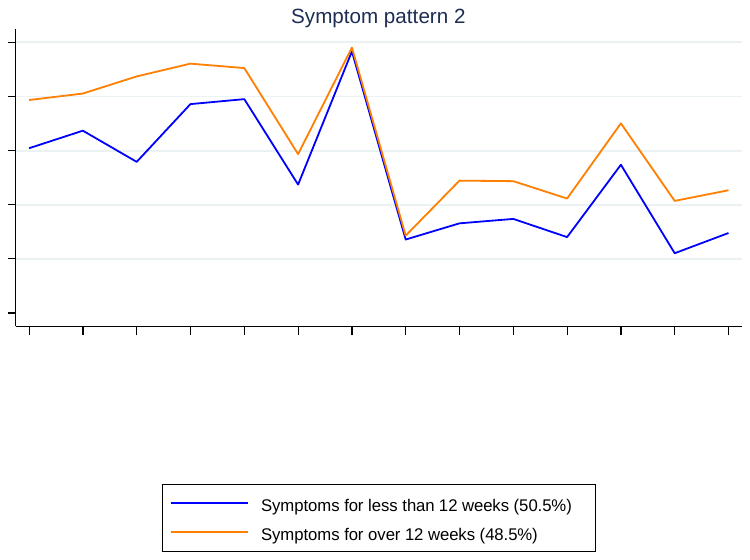

#### **Fig. S3E**. Generation Scotland.

### **Fig. S4**. Symptom probability differences comparing the two COVID-19 groups with the no COVID-19 group in each study (maximal symptom set). Bars represent 95% confidence intervals.

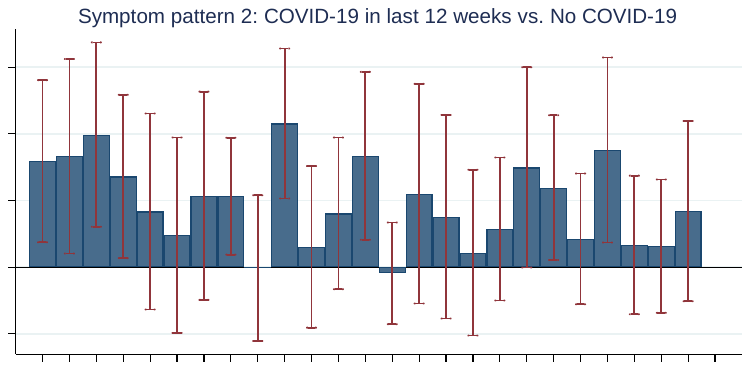

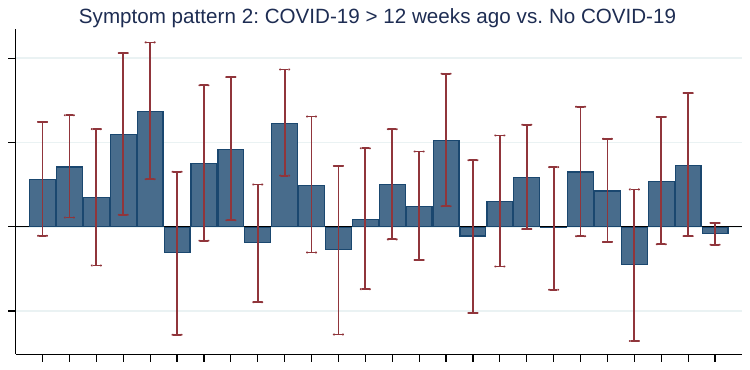

#### **Fig. S4A**. Avon Longitudinal Study of Parents and Children.

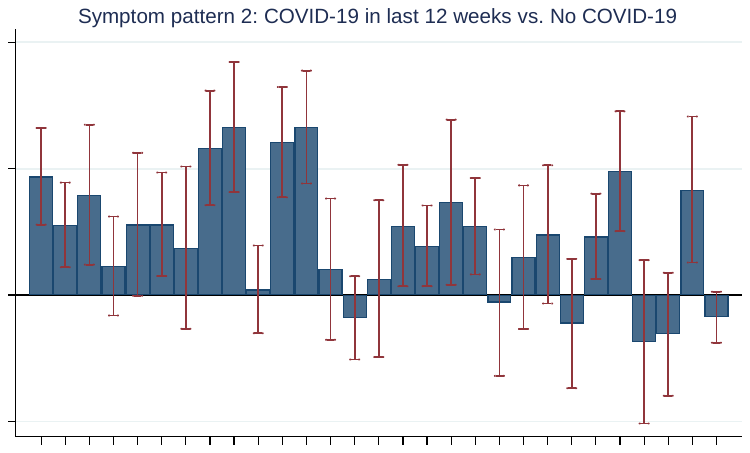

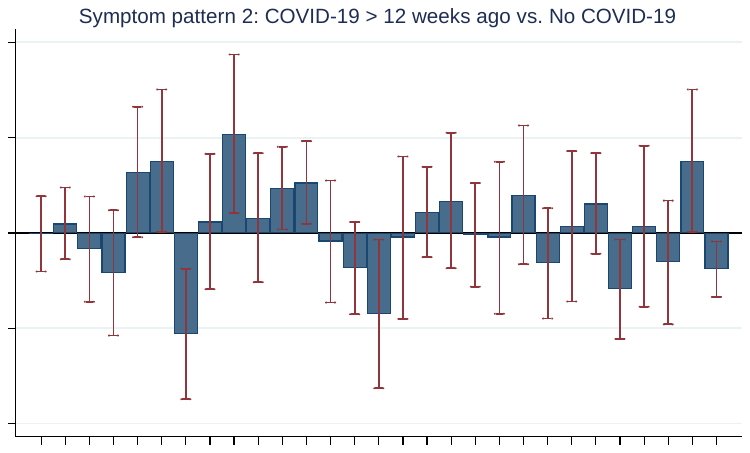

#### **Fig. S4B**. TwinsUK
